# Salient cue reactivity and eating behaviours in ex-smokers, abstinent alcohol use disorder and obesity

**DOI:** 10.64898/2026.03.13.26348339

**Authors:** Katherine E Herlinger, Yong Yong Ling, Liam J Nestor, Nienke J Pannekoek, Moaz Al-Lababidi, Natalie Ertl, Frederica Vanelli, Shahd Alabdulkader, Pallavi Chhibbar, Eva Guerrero, Silvia Canizares, Sri Akavarapu, Marcus R Munafò, Anne R Lingford-Hughes, David J Nutt, Anthony P Goldstone

## Abstract

**Introduction:** Neural cue reactivity is increasingly being investigated as a biomarker of treatment response and relapse prediction in addiction disorders. Whilst aberrant brain responses to salient cues (e.g. drugs) have been widely reported in addiction, it is unclear whether these brain responses persist during longer-term abstinence, how they compare between substance use disorder and obesity, and relate to potential differences in eating behaviours. As part of the Gut Hormones in ADDiction (GHADD) neuroimaging study, we investigated how salient cue reactivity to drugs or food, craving and eating behaviours compare in three clinical populations where alterations have been previously observed: abstinent nicotine use disorder (NUD) and alcohol use disorder (AUD), and obesity.

**Methods:** This study compared group differences in salient cue reactivity and eating behaviours between ex-smokers (n=25, ExS), adults with alcohol dependence who are abstinent (n=26, AAD), adults with obesity who were actively dieting (n=26, OB). Participants completed a high-energy food, preferred alcohol and cigarette functional magnetic resonance imaging (fMRI) cue reactivity task, along with eating behaviour questionnaires, appetite visual analogues scales and an *ad libitum* test meal.

**Results:** ExS exhibited greater blood oxygen level dependent (BOLD) signal to high-energy food pictures in several reward processing regions in both whole brain and region of interest (ROI) analyses, compared with the OB and AAD groups, with no difference in their appeal rating. Compared with the OB group, ExS exhibited greater BOLD signal to cigarette pictures in the frontal gyrus, orbitofrontal cortex, frontal pole and insula, with no difference in their appeal rating. There were no group differences in preferred alcohol cue reactivity. The AAD group rated sweet taste as more pleasant, and consumed more calories from sweet dishes in the *ad libitum* meal than the OB and ExS groups.

**Conclusions:** The presence of heightened cue reactivity to high-energy foods in ex-smokers could contribute to post-quitting weight gain after smoking cessation. Neuroimaging findings were consistent with persistence of some salient drug cue reactivity, despite absence of craving, after medium term abstinence in ExS, but not in AAD. This study also adds to the body of evidence supporting a sweet taste preference endophenotype predisposing individuals to AUD. These changes in eating behaviour in NUD and AUD may provide targets for treatments to reduce substance misuse and facilitate abstinence.

## INTRODUCTION

Tobacco use, harmful alcohol use and obesity are all listed by the World Health Organisation as leading risk factors for developing non-communicable diseases (1). Estimates suggest that tobacco use accounts for 14%, harmful alcohol use 5.3%, and obesity 4.9% of all deaths globally which, taken together, accounts for one quarter of all deaths globally per annum (2–4).

Addiction is a brain disorder, resulting from repeated consumption and exposure to the salient substance in the context of complex genetic and environmental factors (5). Clinically the chronic relapsing nature of addiction is a core problem, and both nicotine and alcohol dependence are particularly challenging to treat. Despite our advances in psychosocial and pharmacological interventions to assist with relapse prevention, over 50% of abstinent smokers will relapse at 12 months, whilst 75% of individuals with abstinent alcohol dependence will relapse at 12 months (6,7).

Obesity, defined as a body max index (BMI) >= 30 kg/m^2^, is similarly complex and challenging to treat. The prevalence of obesity has increased worldwide to pandemic proportions, with no country in the world having a major decrease in obesity prevalence since the 1980s (8,9). As with addiction, “relapse” with weight regain is a common feature after treatment. In one study, 70% of lost weight was regained over 2 years following dietary interventions (10), and ∼65% of weight regain occurred over 1 year after cessation of the GLP-1 and GLP-1/GIP analogue medications, semaglutide and tirzepatide (11,12).

Neuroimaging paradigms have been paramount in characterising the underlying neurobiology of these conditions, and dysregulation of reward processing is reported in both addiction disorders and obesity. Salient cue reactivity is a widely adopted neuroimaging paradigm to assess reward sensitivity and incentive salience to drug and natural rewards such as food. Functional magnetic resonance imaging (fMRI) studies measure blood-oxygen level dependent (BOLD) signal in response to salient cues contrasted with neutral cues or less salient cues (e.g. high-energy (HE) vs. low-energy dense foods) as an indirect measure of neural activity through assessment of the brain’s haemodynamic response. Presentation of a natural reward, drug or drug related stimulus can activate a classically conditioned response eliciting craving and urge to consume. This is often accompanied by physiological arousal and anticipation. Previous meta-analyses of fMRI studies looking at food and drug cue reactivity report activation of overlapping neural networks to both types of salient cue in areas associated with reward, emotion and habit formation (13,14).

Alterations in salient cue reactivity are seen in both substance dependence and obesity. Compared with controls, a recent meta-analysis reported that smokers exhibited increased BOLD signal to smoking cues in the striatum and putamen (15). Similarly, compared with controls, individuals with alcohol dependence display greater BOLD signal to alcohol cues in the medial prefrontal cortex and anterior and middle cingulate cortex (16). Individuals with obesity may also display heightened BOLD signal in response to HE food cues in the striatum compared with healthy weight individuals, providing support for the “food addiction” theory (17,18). However, recent meta-analysis studies have reported mixed evidence for different neural responses to food stimuli in obesity vs. normal weight, highlighting a need for further characterisation (19–22).

Dysregulation towards salient cues in these conditions appears to be associated with clinical outcomes. In substance dependence, the degree of dysregulation towards salient cues has been associated with poorer clinical outcomes such as increased drug use and risk of relapse (23,24). In both nicotine and alcohol dependence, this neural dysregulation may extend into early abstinence, potentially perpetuating relapse (24). Whether full recovery of these neural networks is possible with prolonged abstinence is unknown. Similarly, food cue reactivity and craving at baseline predicts eating and weight gain, while heightened food cue reactivity at baseline has been associated with poorer outcomes following dietary interventions (25–28).

Alterations in eating behaviours and food cue reactivity are seen in nicotine and alcohol dependence. Smoking cessation is associated with a mean weight gain of 4-5kg after 12 months abstinence (29), and also increases in body fat (30), visceral fat accumulation and metabolic risk profile (31). Whilst this weight gain does not counteract the long-term cardiovascular and mortality benefits of stopping smoking (32), concerns regarding weight gain during smoking cessation may undermine smoking cessation efforts and promote relapse (33–36). There is limited evidence for effective long-term dietary and pharmacological strategies to prevent weight gain after smoking cessation (37,38).

The mechanisms behind smoking cessation weight gain are unclear but may be related to loss of anorexigenic actions of nicotine increasing appetite, including through actions on hypothalamic feeding neuropeptides, as well as behavioural substitution from cigarettes to food, including in response to stress (37). One previous fMRI study of current smokers reported blunted BOLD responses in the caudate, putamen, insula and thalamus to personalised favourite food auditory cues compared with non-smokers, suggesting attenuated food cue reactivity in brain reward systems of current smokers (39). Smokers also had increased hypothalamic BOLD signal to ingestion of milkshake than non-smokers, a region whose response positively correlated to weight increase over 1 year in non-smokers (40). Whether these alterations in food cue and taste reactivity are reversed following smoking cessation and contribute to post-smoking cessation weight gain is currently unknown.

Similarly, depressed appetite and reduced food intake are typically seen in long-term dependent alcohol drinkers, often resulting in malnutrition (41). One recent fMRI study looking at the effect of “problem drinking” reported a negative correlation between Alcohol Use Disorders Identification Test (AUDIT, (42) scores and BOLD signal to palatable food cues, grey matter volume and functional connectivity strength in the lateral orbitofrontal cortex (OFC) (43). Activation of the lateral OFC has been consistently implicated in visual food cue exposure, food appeal, pleasant taste and taste anticipation in healthy controls (44,45). Therefore, attenuated food cue reactivity in alcohol dependence may contribute to decreased food intake, but again it is unknown if this reverses during abstinence from alcohol.

Abnormalities in taste for sweet or fat have not been consistently seen in obesity nor to normalise after bariatric surgery induced-weight loss (46–48). However, alcohol consumption may have links to individual differences in sweet taste preference, with a sweet-liking endophenotype identified in some, but not all, studies of alcohol dependence, that may persist at least into early abstinence, and which may confer vulnerability to relapse (49–53). It remains uncertain if there are abnormalities in sweet liking with longer-term abstinent alcohol dependence, and if this is accompanied by other alterations in eating behaviour.

This current study hypothesised that in brain reward circuitry, (i) ex-smokers would display heightened cigarette cue reactivity compared with never-smokers, (ii) ex-smokers would display heightened HE food cue reactivity compared with individuals with alcohol dependence who are abstinent (AAD) and adults with obesity, while (iii) those with AAD would display heightened alcohol cue reactivity compared with those without AUD. Furthermore, it was hypothesised that: (iv) those with AAD would display heightened sweet food preference.

This cross-sectional, multi-modal phenotyping study therefore aimed to compare salient cue reactivity using a picture evaluation fMRI task (HE food, cigarette, preferred alcohol), eating behaviour questionnaires, visual analogue scales ratings of appetite, taste ratings and *ad libitum* food intake of savoury/sweet and low/high fat foods in adults with obesity, ex-smokers and AAD.

## METHODS

This was a secondary analysis of our MRC funded Gut Hormones in ADDiction (GHADD) neuroimaging study that used a randomised, within-participant, cross-over design to investigate how acute administration of the appetitive hormones exendin-4 (a glucagon-like peptide-1 (GLP-1) analogue) or desacyl ghrelin (vs. saline, administered at three separate study visits) altered addictive and eating behaviours in obesity, ex-smokers and AAD, by using the study visit outcomes from the saline visit only to compare between groups (https://clinicaltrials.gov/study/NCT02690987).

### Participants

Inclusion criteria for all participants were as follows: (i) aged 18-60 years, (ii) females or males. Inclusion criteria for healthy controls (HC) included: (i) BMI between 18.0-28.0 kg/m^2^, (ii) <100 lifetime cigarettes. Inclusion criteria for adults with obesity (OB): (i) BMI between 28.0-50.0 kg/m^2^, (ii) self-reported active dieting, (iii) <100 lifetime cigarettes. Inclusion criteria for ex-smokers (ExS) included: (i) previous nicotine dependence as determined by (a) DSM-V criteria for NUD (54), who (b) when smoking, had their first cigarette within 60 mins of awakening, and (c) previously smoking at least five cigarettes daily, (ii) in stable abstinence for at least 6 weeks, (iii) off nicotine replacement therapy for at least 2 weeks. Inclusion criteria for abstinent alcohol dependent participants (AAD) included: (i) previous alcohol dependence as determined by DSM-5 criteria for moderate-severe AUD (54), i.e. moderate 4-5/11, severe ≥6/11 criteria, (ii) in stable abstinence for at least 6 weeks, (iii) any history of cigarette use, including never smoked, current smokers or ex-smokers, regardless of NUD severity. No serious medical or mental illnesses were allowed for any group. Previous problem gambling, previous substance dependence and current depressive disorder or antidepressant medication was only allowed in the AAD group. For full inclusion and exclusion criteria see Supplementary Methods and Supplementary Table S1.

Participants were recruited using public advertisements. The study was approved by the local research ethics committee (15/LO/1041) and performed in accordance with the principles of the Declaration of Helsinki. All participants provided written informed consent. Recruitment for this study ran from 15^th^ July 2015 until 15^th^ August 2019.

### Study protocol

During experimental sessions, participants were monitored at a total of 10 time points over 7.5 hours for heart rate, blood pressure, glucose measurements and venous sampling. During these time points they rated their appetite and food, alcohol and cigarette craving using visual analogue scales. Throughout the day participants also completed a variety of computer-based questionnaires and neurocognitive tasks.

In the scanner participants performed several fMRI tasks including HE food, cigarette and preferred alcohol picture evaluation task. While viewing the pictures, participants rated simultaneously their appeal. Following the scan, participants ate a further snack and then an *ad libitum* lunch (see Supplementary Table S2 for details). Results of questionnaires, neurocognitive tasks and fMRI paradigms that have not been presented here will be described in future publications.

Additional details about the study protocol are given in Supplementary Methods, and Supplementary Figure S1.

### MRI scanning protocol

Each participant underwent an 80-min MRI session starting at T=+115 min (3T Siemens Verio MRI scanner, Invicro, Hammersmith Hospital, London, UK). Each scanning session included a resting state fMRI scan, picture evaluation fMRI paradigm (55,56), monetary incentive delay task (57) and a negative emotional reactivity task (58,59). Structural MR brain scans including high-resolution T1-weighted scans were also collected. Additional details about scanning parameters are given in Supplementary Methods.

### Picture evaluation fMRI paradigm

Four types of colour photographs were presented in a block design split across two 10 mins run with only one picture category shown per block: (1) 60 HE food images (e.g. chocolate, cake, pizza), (2) 60 cigarette images (e.g. held cigarettes, smoked cigarettes), (3) 60 alcohol images, personalised to their two types of individualised preferred alcoholic drinks (e.g. beer, wine, spirits), and (iv) 60 neutral images (e.g. household objects, hands, faces). Food images were selected to represent familiar foods that are typical of a Western diet. Pictures were obtained from freely available websites and the International Affective Picture System (60).

While each image was on display, the participants were asked to simultaneously rate how appealing they found each picture by using a 5-finger button box (1 = not at all, 2 = not really, 3 = neutral, 4 = a little, and 5 = a lot), so the appeal rating was made and recorded simultaneously with the stimulus presentation, as used previously (45,55,56).

Given the major effect of nutritional status on food cue reactivity (45,55,61–63), healthy controls (HC) were excluded from analysis of the picture evaluation fMRI task as they had a different nutritional status, having had their usual breakfast at home. Participants with abstinent alcohol dependence (AAD) were excluded from analysis of the cigarette picture fMRI evaluation task as the group was a heterogenous mixture of never smoked, current smokers and ex-smokers.

Additional details for the fMRI paradigms are given in Supplementary Methods.

### Image pre-processing

Additional details about image pre-processing are given in Supplementary Methods. fMRI data processing was carried out with Functional Magnetic Resonance Imaging of the Brain (FMRIB) Software Library (FSL) v5.0.10 (https://fsl.fmrib.ox.ac.uk/fsl/fslwiki). One participant in the group with obesity was excluded from the fMRI analysis as there was excess head movement (average relative motion across both picture runs >0.5 mm/volume).

For each individual picture type, a second level analysis was carried out using a fixed-effect model to average across the 2 runs in FLAME (FMRIB’s Local Analysis of Mixed Effects) software. This was determined for the contrasts: HE food > neutral, cigarette > neutral and preferred alcohol > neutral.

### Functional region of interest analysis

Additional details about the region of interest analysis are given in Supplementary Methods, and Supplementary Figure S2.

Nine *a priori* functional regions of interest (fROIs) were chosen from previous fMRI studies using food-picture paradigms that have been shown to be modulated by nutritional status, top-down inhibitory control, gut hormone administration and bariatric surgery (45,55,56,64–66): nucleus accumbens (NAcc), caudate, putamen, anterior insula (Ins), anterior orbitofrontal cortex (OFC), amygdala, hippocampus, ventral anterior cingulate cortex (vACC) and dorsolateral prefrontal cortex (dlPFC).

The spatial extent of these fROIs was determined from the identical picture evaluation fMRI paradigm performed in a cohort of n=43 healthy, adults without obesity (including n=24 whose non-fMRI outcome measures are included in this study), who were scanned having had their usual breakfast at home and who did not receive any hormone or saline infusion (Supplementary Tables S3 and S4).

### Whole brain analysis

To examine the spatial extent of group differences in BOLD signal for each contrast, whole brain analysis was conducted using FEAT software. Inter-group differences were examined as a whole brain mixed effects between participants one-way ANOVA for HE food > neutral and alcohol > neutral contrasts (OB, ExS and AAD groups) and as unpaired t-tests for HE food > neutral contrast for OB vs. ExS, OB vs. AAD, ExS vs. AAD group comparisons) and for cigarettes > neutral contrast (OB vs. ExS groups only) with threshold cluster-wise Z>2.3, P<0.05 to control for multiple comparisons.

To further examine the nature of the inter-group differences in the regions identified from the whole brain ANOVA activation map, this functional map was masked with relevant anatomical ROIs defined from the cortical and subcortical structural Harvard FSL atlas thresholded at 10% probability. FSL Featquery was then used to extract the median % BOLD signal within each region, and a post-hoc repeated measures ANOVA to compare groups within each functional ROI performed using SPSS version 28.0 software (IBM Corp. Released 2021. IBM SPSS Statistics for Macintosh, Version 28.0. Armonk, NY: IBM Corp).

### Eating behaviour questionnaires

Eating behaviour was assessed on either the screening or one of the study visits through administration of several computer-based questionnaires: Yale Food Addiction Scale (YFAS) (67), Power of Food Scale (PFS) (68), Dutch Eating Behaviour Questionnaire (DEBQ) to measure dietary restraint, external eating and emotional eating (69), Three-factor Eating Questionnaire (TEFQ) to measure dietary restraint, disinhibited eating and hunger-related eating (70) and Binge Eating Scale (BES) (71).

### Visual analogue scales

Using a touchscreen iPad, participants rated hunger, fullness, volume able to eat, pleasantness to eat using a continuous 0-100 mm visual analogue scales (VAS) ratings at T= −35, −10, +45, +90, +210, +270, +295, +315, +370, and +400 mins from the start of the infusion (Figure 1) (55). A composite appetite VAS rating was calculated from individual questions using the equation: Appetite = [Hunger + Pleasantness to eat + Volume to eat + (100 – Fullness)]/4.

**Figure 1.**
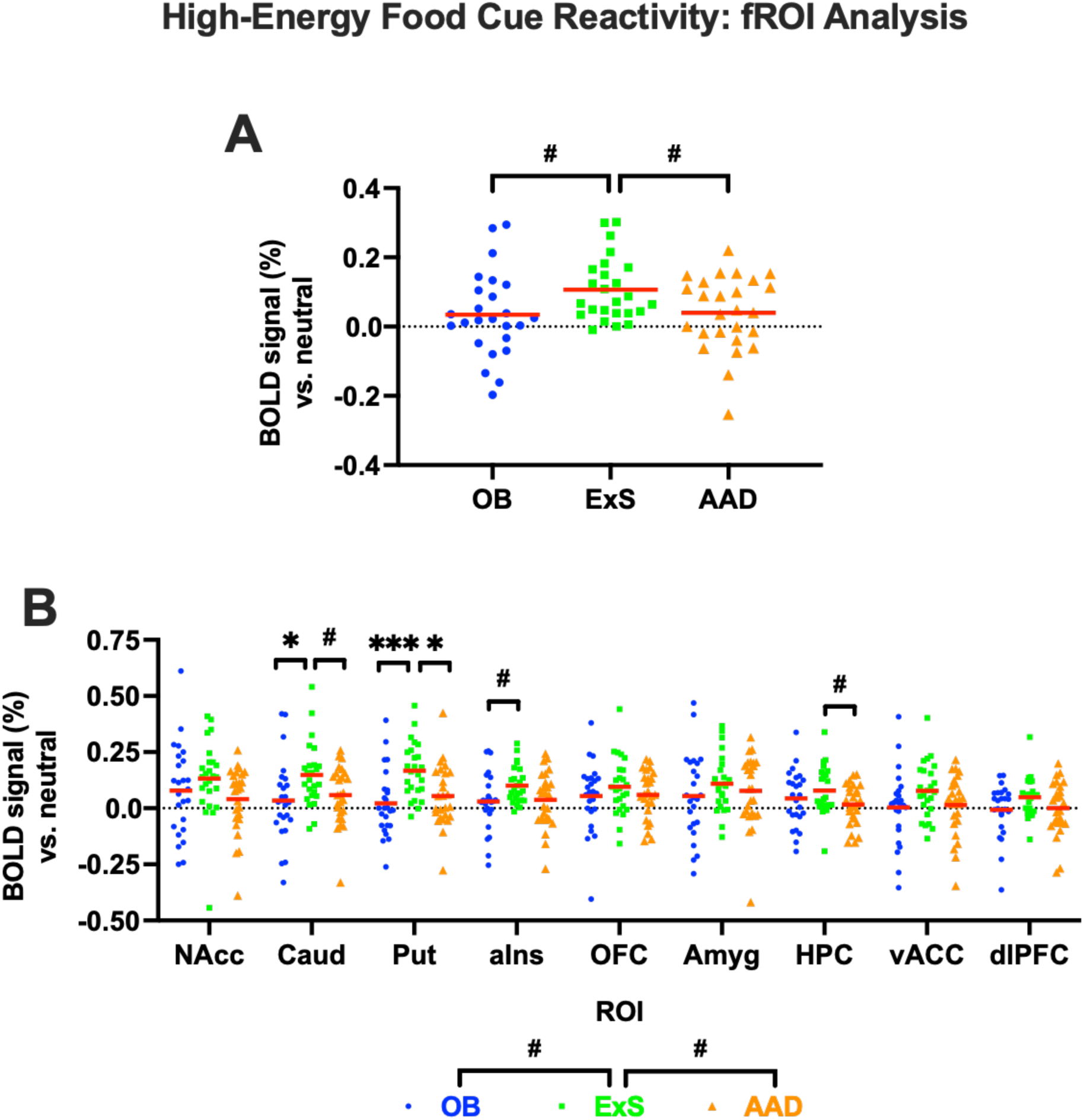
Group differences in high-energy food cue reactivity from functional region of interest analysis. Percentage BOLD signal to high-energy food pictures (vs. neutral) during picture evaluation fMRI task in adults with obesity (OB, blue circle, n=25), ex-smokers (ExS, green square, n-25) and abstinent alcohol dependence (AAD, orange triangle, n=26), when (A) averaged across all nine functional regions of interest (fROI), or (B) in individual fROI: bilateral nucleus accumbens (NAcc), caudate (Caud), putamen (Put), anterior insula (Ins), anterior orbitofrontal cortex (OFC), amygdala (Amyg), hippocampus (HPC), ventral anterior cingulate cortex (vACC), and dorsolateral prefrontal cortex (dlPFC). Data presented as individual data points with red line at mean. Compared using one-way repeated measures ANOVA with post-hoc LSD test: ^#^P<0.05, ^##^P<0.01, ^###^P<0.001, or post-hoc Sidak tests: *P<0.05, **P<0.01, ***P<0.001 (to correct for multiple comparisons). See Supplementary Figure S2B for anatomical location of fROIs, and Supplementary Table S4 for Montreal Neurological Institute (MNI) coordinates of fROIs.

At each of these time points, participants also rated alcohol craving using the Alcohol Urge Questionnaire (AUQ, score range 8-56) (72), cigarette craving using the Questionnaire of Smoking Urges, (QSU-brief, score range 10-70) (73), and food craving (using 2 questions “All I want to do is to eat right now” and “Nothing would be better than eating right now”, max score 14), using a 7-point Likert scale from strongly disagree to strongly agree (score range 2-14).

For each rating, absolute area under the curve (AUC_-10-315min_) was calculated using the trapezoid rule from timepoint 2 (t=-10 min) to timepoint 8 (t=+315 min), omitting the initial rating to avoid effects of stress on arrival and after cannulation, and ratings after the *ad libitum* meal given variability in amount eaten. The AUC_-10-315min_ value was then converted into an average AUC score to give a more meaningful number over this period by dividing by 325.

### Ad libitum lunch

At t=+325 min, an *ad libitum* meal of four dishes was presented consisting of tomato broth (low fat, savoury), cream of tomato soup (high fat, savoury), vanilla yoghurt (low fat, sweet) and ice cream (high fat, sweet) with each food type served in excess (see Supplementary Table S2). N=1 participant in the ExS group reported at the screening visit that they did not like tomato soup and so instead were served chicken broth and cream of chicken soups.

Taste was rated for each dish using the Sussex Ingestion Pattern Monitoring rating scales from 0-100mm (SPIM, University of Sussex) (74) for creaminess, sweetness, ‘just right’ for both creaminess or sweetness (using the intensity general Labelled Magnitude Scale, gLMS), and liking and pleasantness (using a linear scale), before participants were allowed to eat as much of each dish as they wished. Food intake for each dish was calculated as both absolute total energy intake (kcal) and energy intake as a percentage of estimated 24hr resting energy intake (kcal as % REE),

Additional methodology details about the *ad libitum* lunch are given in Supplementary Methods.

### Statistics

Results are presented as mean ± SEM for parametric data or medians (IQR) for non-parametric data. Comparisons between groups for demographic and behavioural data and ratings of appetite and craving were made using a one-way ANOVA with post-hoc Fisher LSD test, and Sidak test for correction for multiple comparisons, if normally distributed; while for non-parametric data a Kruskall-Wallis test was used with post-hoc Bonferroni correction for multiple comparisons.

For neuroimaging data from whole brain analyses, comparisons between groups were made using a one-factor repeated measures ANOVA with post-hoc Fisher LSD test, and Sidak test for correction for multiple comparisons.

The energy intake consumed at the *ad libitum* test meal was analysed as a Kruskall-Wallis test with post-hoc Bonferroni correction for multiple comparisons.

Energy intake and taste ratings of individual dishes and of different food categories was analysed using repeated measures ANOVA, including sweetness (savoury, sweet) and fat content (low fat, high fat) as within-participant, and group as between participant factors with post-hoc Fisher LSD and Sidak correction for multiple comparisons.

The significance level was set at P<0.05 and SPSS software version 28.0 (IBM Corp. Released 2021. IBM SPSS Statistics for Macintosh, Version 28.0. Armonk, NY: IBM Corp) was used for all statistical analyses.

## RESULTS

### Participants

The final number of participants recruited for the study were n=24 healthy controls without obesity (HC), n=26 adults with obesity (OB), n=25 ex-smokers (ExS) and n=26 individuals with AUD who are abstinent (AAD). One participant in the OB group was excluded from the neuroimaging dataset due to excess signal drop-out, leaving n=25 in the OB group for the fMRI analysis. Similarly, one other participant in the OB group declined to eat any of any of the *ad libitum* test meal as they disliked the type of food served, and so were excluded from analysis of the lunch energy intake and taste ratings, leaving n=25 in the OB group. An additional participant in the OB group did not complete all hormone infusion study visits, and so not all eating behaviour trait questionnaires were administered, and one participant in HC group did not complete all questionnaires at their single visit; therefore, for some questionnaires, data is only available for n=25 for OB and n=23 for HC.

Participant characteristics are listed in Table 1. The three clinical groups (OB, ExS, AAD) were well matched for age, sex ratio, ethnicity and years spent in education. HC were also well matched for sex, ethnicity and years in education with the clinical groups; however, they were significantly younger than OB, ExS and AAD groups. There was no significant difference in abstinence duration for cigarettes and alcohol between the ExS and AAD groups respectively, with median 20.7 and 32.6 weeks. As per protocol, all HC and OB participants were classified as ‘never smokers’ (lifetime smoked <100 cigarettes), whilst approximately half of the AAD were either ex-smokers (46.23%, median duration of abstinence 117 weeks, retrospective Fagerstrom score from time before quitting 3.58/10) or current smokers (46.2%, Fagerstrom score 3.72/10).

**Table 1.** Participant characteristics. Data displayed as mean ± SD (range) or median [interquartile range]. One-way ANOVA with post-hoc Sidak test for parametric data, Kruskal-Wallis test with Bonferroni corrected post-hoc test for non-parametric data and nominal data. Post-hoc tests: vs. HC *P<0.05, **P<0.01, ***P<0.001; vs. OB ^#^P<0.05, ^##^P<0.01, ^###^P<0.001; vs. ExS ^†^P<0.05, ^††^P<0.01, ^†††^P<0.001. ^a^ OB n=24. ^b^ Abstinence weeks from cigarettes. ^c^ Abstinence weeks from alcohol. n/a=not applicable.

|  | Healthy controls (HC) | Obesity (OB) | Ex-smokers (ExS) | Abstinent alcohol dependence (AAD) | Test statistic | P-value |
| --- | --- | --- | --- | --- | --- | --- |
| n | 24 | 26 | 25 | 26 |  |  |
| Age (years) | 24 [23, 30] | 46 [31, 53] *** | 36 [29, 44] *** | 44 [35, 50] *** | H(3)=34.5 | <0.001 |
| range | (20-51) | (24-60) | (25-60) | (21-59) |  |  |
| Females, n (%) | 12 (50.0%) | 18 (69.2%) | 13 (52.0%) | 13 (50.0%) | H(3)=2.7 | 0.44 |
| Caucasian, n (%) | 14 (58.3%) | 17 (65.4%) | 21 (84.0%) | 22 (84.6%) | H(3)=6.7 | 0.083 |
| Years of education | 16 [16, 18] | 16 [14, 18] | 17 [15, 18] | 14 [12, 19] | H(3)=4.8 | 0.19 |
| Index of Mutiple Deprivation (2015), max score 92.6 | 18.3 ± 8.7 (3.1-33.6) | 23.0 ± 12.0 (4.6-49.4) | 21.3 ± 11.2 (2.9-43.2) | 19.7 ± 9.5 (3.0-36.3) | F(3,97)=0.9 | 0.43 |
| Verbal intelligence WTAR, max score 50 | n/a | 43 [38, 46] | 45 [40, 48] | 45 [39, 48] | H(3)=1.8 | 0.41 |
| Current smokers, n (%) | 0 (0%) | 0 (0%) | 0 (0%) | 12 (46.2%) |  |  |
| Ex-smokers, n (%) | 0 (0%) | 0 (0%) | 25 (100%) | 11 (42.3%) |  |  |
| Fagerstrom (nicotine dependence), max score 10 | n/a | n/a | 4 [3, 6] | 0 [0, 4]<sup>†††</sup> | H(1)=15.4 | <0.001 |
| AUDIT, max score 40 | 7 [3, 9] | 3 [2, 5]<sup>a</sup> | 7 [4, 11]<sup>#</sup> | 30 [18, 34] *** ####<sup>†††</sup> | H(3)=61.3 | <0.001 |
| DSM 5 alcohol use disorder, max score 11 | 1 [0, 1] | 0 [0, 1] | 1 [0, 2] | 10 [9, 11] *** ####<sup>†††</sup> | H(3)=63.8 | <0.001 |
| Abstinence length (weeks) | n/a | n/a | 20.7 [12.7, 55.0]<sup>b</sup> | 32.6 [15.1, 103.7]<sup>c</sup> | H(3)=2.0 | 0.16 |
| Body mass index (BMI, kg/m<sup>2</sup>) | 22.2 ± 2.6 (17.5-28.5) | 37.0 ± 4.7 (29.6-46.3) *** | 25.5 ± 3.4 (18.9-32.1) * #### | 25.7 ± 4.8 (19.2-37.1) * #### | F(3,97)=66.9 | <0.001 |
| % body fat (BIA, adjusted for female sex) | 22.0 ± 6.3 | 39.2 ± 6.4 *** | 26.3 ± 6.3 #### | 26.5 ± 6.3 #### | F(4,96)=48.0 | <0.001 |
| Current antidepressant, n (%) | 0 (0%) | 0 (0%) | 0 (0%) | 9 (34.6%) |  |  |
| Previous depression, n (%) | 0 (0%) | 3 (11.5%) | 6 (24.0%) | 18 (69.2%) *** ####<sup>††</sup> | H(3)=35.5 | <0.001 |
| BDI-II, max score 63 | 2 [0, 4] | 3 [1, 8]<sup>a</sup> | 4 [0, 6] | 5 [1, 10] | H(3)=5.5 | 0.14 |
| Family history of obesity, n (%) | 3 (12.5%) | 16 (61.5%) ** | 6 (24.0%)<sup>#</sup> | 2 (7.7%) #### | H(3)=23.2 | <0.001 |
| Family history of smoking, n (%) | 8 (33.3%) | 13 (50.0%) | 21 (84.0%) ** | 14 (53.9%) | H(3)=13.2 | 0.004 |
| Family history of alcohol dependence, n (%) | 0 (0%) | 2 (7.7%) | 9 (36.0%) * | 10 (38.5%) **<sup>#</sup> | H(3)=17.8 | <0.001 |

The OB group had a significantly higher BMI compared with the other 3 groups, while HC had a lower BMI than ExS and AAD groups, who had similar BMI to each other (Table 1). As per protocol, the AAD group had a retrospective median DSM-5 alcohol score (from time when still drinking alcohol) of 10/11 with all individuals above 6/11, indicating severe AUD, whilst no individual in the other groups met DSM-5 criteria for severe AUD.

All the AAD group had severe AUD by DSM-V criteria with score range 7-11/11. Additionally, AAD participants were more likely to have a past medical history of depression compared with the other groups; however, there was no significant difference in the current BDI-II scores between the groups. Nine out of 26 (34.6%) of the AAD group (but none in the other groups) were currently taking anti-depressant medication.

### Picture evaluation fMRI task

#### High-energy food cue reactivity

##### Functional region of interest analysis

FROI analysis of group differences in BOLD signal to HE food pictures in the three clinical groups (OB, ExS, AAD) revealed no significant group x ROI interaction [F(10.3,374.4)=1.3, P=0.23, with Greenhouse-Geisser correction], but a significant overall effect of group, independent of ROI [F(2,73)=3.45, P=0.037]. This was driven by increased BOLD signal to HE food pictures (averaged across all nine fROIs) in ExS compared with both OB [effect size mean ± SEM 0.072 ± 0.030 (95% CI 0.011, 0.132), d=0.65, P=0.021 Fisher LSD, P=0.063 Sidak] and AAD [effect size mean ± SEM 0.067 ± 0.030 (95% CI 0.006, 0.127), d=0.61, P=0.031 Fisher LSD, P=0.089 Sidak corrected] groups (Figure 1A). However, BOLD signal to HE foods was not significantly different between the OB and AAD groups [effect size mean ± SEM 0.005 ± 0.030 (95% CI −0.055, 0.065), d=0.87, P=0.87 Fisher LSD, P=0.10 Sidak corrected].

Further exploratory post-hoc analysis indicated that the ExS group had significantly greater BOLD signal to HE food pictures than the OB group in the caudate, putamen and insula; but not the NAcc, aOFC, amygdala, hippocampus, vACC, or dlPFC regions (Figure 1B, Supplementary Table S5). ExS also displayed significantly greater BOLD signal to HE food pictures than the AAD group in the caudate, putamen and hippocampus; but not the NAcc, insula, aOFC, amygdala, vACC, or dlPFC regions (Figure 1B, Supplementary Table S5). There were no significant differences in BOLD signal to HE food pictures between the OB and AAD groups in any of the fROIs (range d=0.05-0.88, range P=0.37-0.88 Fisher LSD) (Figure 1B).

##### Whole brain analysis - between all groups

Whole brain activation maps for each group (OB, ExS, AAD) for BOLD signal to HE food (vs. neutral) pictures are given in Supplementary Table S6, Supplementary Figures S3-5.

Whole brain analysis of differences between the three clinical groups (OB, ExS, AAD) revealed one significant cluster showing group differences in BOLD signal to HE food pictures (Figure 2A, Supplementary Table S7 for coordinates). This cluster was primarily made up of 3 anatomical sub-regions: putamen, caudate and insula (Figure 2B).

**Figure 2.**
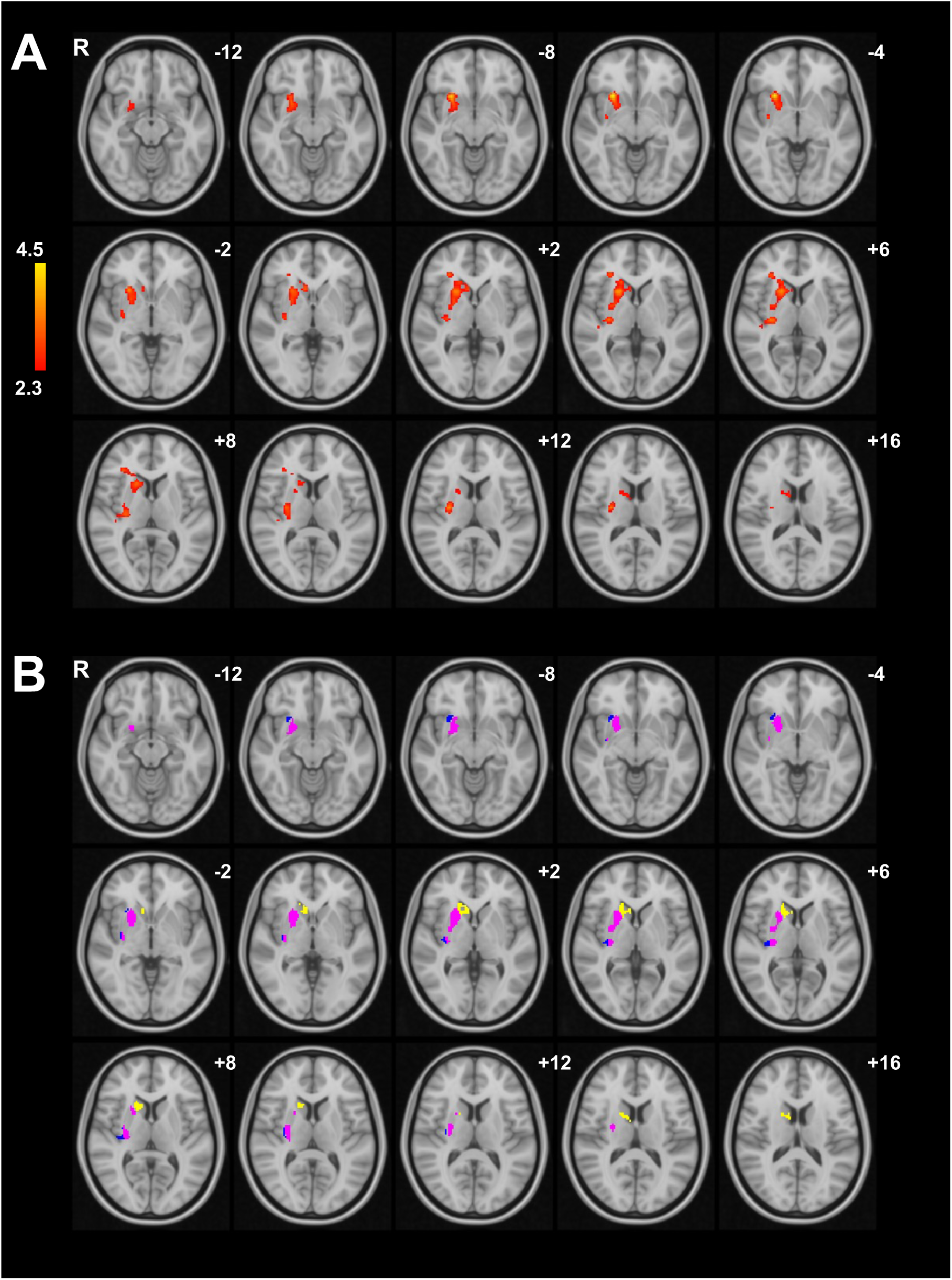
Group differences in high-energy food cue reactivity from whole brain analysis. Whole brain analysis showing cluster with between group differences in percentage BOLD signal to high-energy food pictures (vs. neutral) during picture evaluation fMRI task in adults with obesity (OB, n=25), ex-smokers (ExS, n=25) and abstinent alcohol dependence (AAD, n=26), showing (A) overall cluster, and (B) anatomical regions within this cluster in putamen (pink), caudate (yellow) and insula (blue). One-way ANOVA, cluster-wise threshold Z>2.3, family-wise error (FWE) P<0.05. z co-ordinates given in Montreal Neurological Institute (MNI) space. Abbreviations: R, right. See Supplementary Table S7 for MNI co-ordinates.

For these 3 sub-regions within the cluster, post-hoc testing revealed no significant group x sub-region interaction in BOLD signal to HE food pictures [F(3.72,135.8)=2.07, P=0.094, with Greenhouse-Geisser correction] but a significant overall effect of group, independent of sub-region [F(2,73)=9.91, P<0.001]. This was driven by the ExS group having significantly greater BOLD signal to HE food pictures averaged across the 3 sub-regions compared both the OB and AAD groups (Figure 3A, Supplementary Table S8), but BOLD signal to HE foods was similar in the OB and AAD groups.

**Figure 3.**
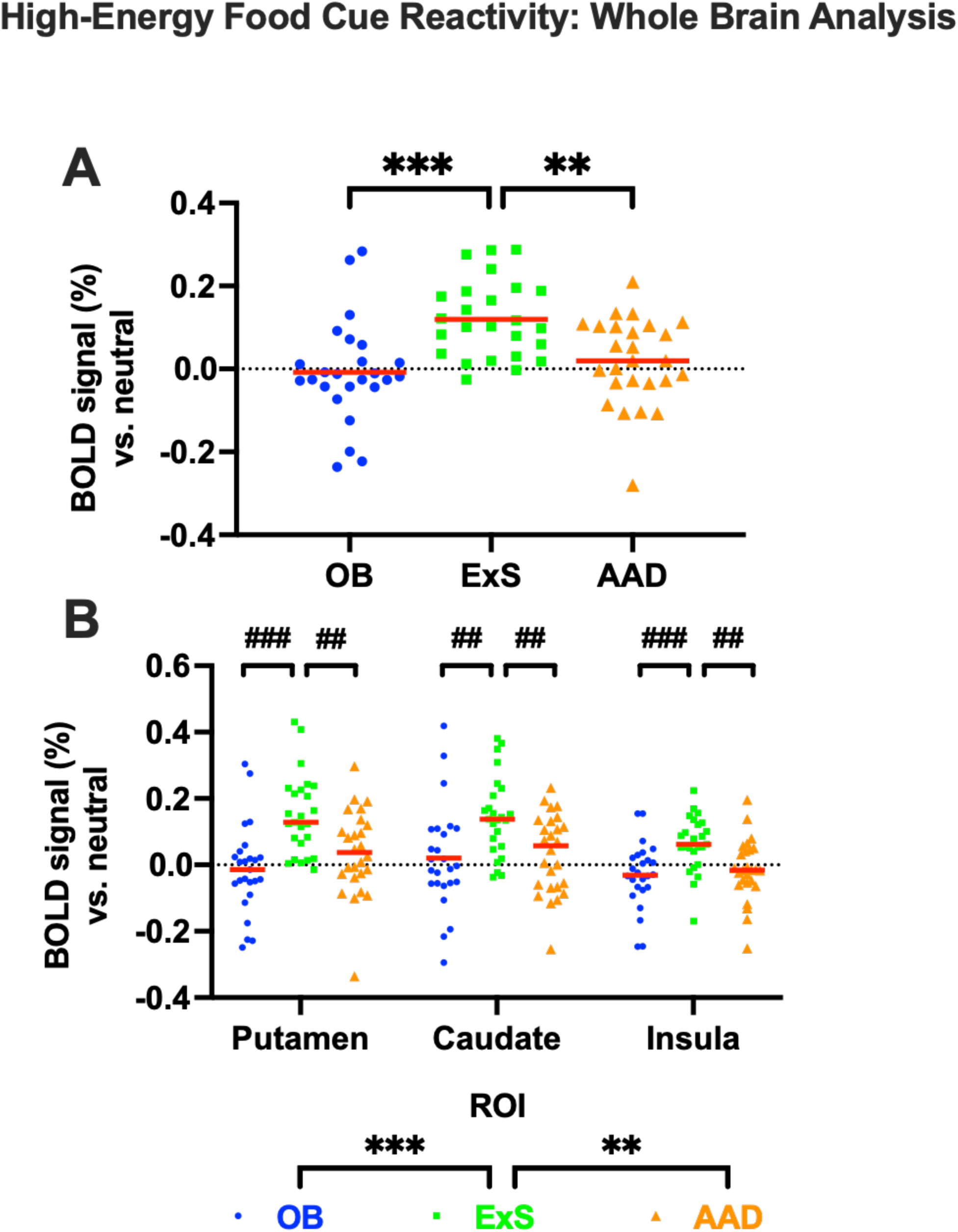
Greater high-energy food cue reactivity in putamen, caudate and insula in ex-smokers compared to obesity and abstinent alcohol dependence from whole brain analysis of picture evaluation fMRI task. Between group differences in percentage BOLD signal to high-energy food pictures (vs. neutral) in obesity (OB, blue circle, n=25), ex-smokers (ExS, green square, n=25) and abstinent alcohol dependence (AAD, orange triangle, n=26), in picture evaluation fMRI task (A) averaged across significant cluster from whole brain analysis, (B) in individual anatomical sub-regions within this cluster: bilateral putamen, caudate and insula. Data presented as individual data points with red line at mean. Compared using one-way ANOVA with post-hoc LSD test: ^#^P<0.05, ^##^P<0.01, ^###^P<0.001; Sidak tests (to correct for multiple comparisons): *P<0.05, **P<0.01, ***P<0.001. See Figure 2 for brain activation map, Supplementary Table S7 for Montreal Neurological Institute (MNI) co-ordinates, Tables S8 and S9 for statistical comparisons.

In further exploratory post-hoc analysis of individual sub-regions, the ExS group had significantly greater BOLD signal to HE foods than the OB group in the caudate, putamen, and insula (Figure 3B, Supplementary Table S9). The ExS group also had significantly greater BOLD signal to HE foods than the AAD group in the caudate, putamen and insula (Figure 3B, Supplementary Table S9). BOLD signal to HE foods was similar between OB and AAD groups in the caudate, putamen and insula (Figure 3B, Supplementary Table S9).

### Whole brain analysis - pairwise comparisons

#### Ex-smokers vs. obesity

Compared to the OB group, the ExS group had greater BOLD signal to HE foods in one cluster which included regions of the putamen and insula cortex (Supplementary Table S10, Supplementary Figure S6). There were no regions in which BOLD signal to HE foods was greater in the group with OB than ExS group.

#### Ex-smokers vs. abstinent alcohol dependence

Compared with the AAD group, the ExS group had increased BOLD signal to HE foods in four clusters, that included respectively (i) frontal pole and OFC; (ii) pallidum, putamen and hippocampus; (iii) SFG, frontal pole, paracingulate gyrus, anterior dorsal pre-motor cortex and SMA; (iv) insula cortex, OFC, cingulate gyrus, caudate, Nacc and inferior frontal gyrus (Supplementary Table S10, Supplementary Figure S6). There were no regions in which BOLD signal to HE foods was greater in the AAD than ExS group.

#### Obesity vs. abstinent alcohol dependence

There were no significant group differences between the group with OB and AAD group.

#### High-energy food picture appeal ratings

There were no significant difference in mean appeal ratings for the HE food (vs. neutral) pictures between the three clinical groups OB, ExS and AAD [F(2,73)=1.49, P=0.23] (Figure 5).

#### Influence of BMI on HE food cue reactivity in obesity

There were no significant correlations of BMI with BOLD signal to HE foods in the group with obesity from either fROI or whole brain analyses, nor with HE food (vs. neutral) appeal (see Supplementary Results).

### Alcohol cue reactivity

#### Functional region of interest analysis

In fROI analysis of BOLD signal to preferred alcohol pictures, there was no significant group x ROI interaction [F(9.96,363.4)=0.58, P=0.83 Greenhouse-Geisser correction], nor overall effect of group, independent of ROI [F(2,73)=0.49, P=0.62], between the OB, ExS and AAD groups (Supplementary Figure S7).

#### Whole brain analysis - between all groups

Separate whole brain activation maps for each group (OB, ExS, AAD) for BOLD signal for preferred alcohol (vs. neutral) pictures are given in Supplementary Table S11, Supplementary Figures S8-S10. In none of the groups was there greater BOLD signal to alcohol vs. neutral pictures in brain regions involved in reward processing, as this was only seen in areas such as the cuneus and precuneus which includes part of the default mode network.

In between group whole brain analysis, there were no significant differences in BOLD signal when viewing alcohol (vs. neutral) pictures between the OB, ExS and AAD groups.

#### Whole brain analysis - pairwise comparisons

There were no significant differences in BOLD signal to preferred alcohol pictures for pairwise comparisons between any of the OB, ExS or AAD groups.

#### Alcohol picture appeal rating

There were no significant differences in appeal ratings for preferred alcohol (vs. neutral) pictures between the OB, ExS and AAD groups [F(2,73)=0.30, P=0.74] (Figure 5).

### Cigarette cue reactivity

#### Functional region of interest analysis

In fROI analysis of BOLD signal to cigarette pictures between groups with OB and ExS, there was no significant group x ROI interaction [F(4.3,205.8)=0.85, P=0.50; Greenhouse-Geisser (GG) correction], nor any overall effect of group, independent of ROI [F(1,480)=0.4, P=0.53] (Supplementary Figure S11).

#### Whole brain analysis - pairwise comparisons

Separate whole brain activation maps for each group (OB, ExS) for BOLD signal to cigarettes (vs. neutral) pictures are given in Supplementary Table S12, Supplementary Figures S12 and S13.

In whole brain analysis, the group with ExS had greater BOLD signal to cigarettes compared with the group with OB group in two clusters including respectively regions of the middle frontal gyrus, inferior frontal gyrus pars opercularis, frontal operculum cortex, OFC, insula and frontal pole; and the frontal operculum cortex, OFC, insula, inferior frontal gyrus pars triangularis and frontal pole (Figure 4, Supplementary Table S13). There were no significant clusters in which the OB group had higher BOLD signal to cigarettes then the ExS group.

**Figure 4.**
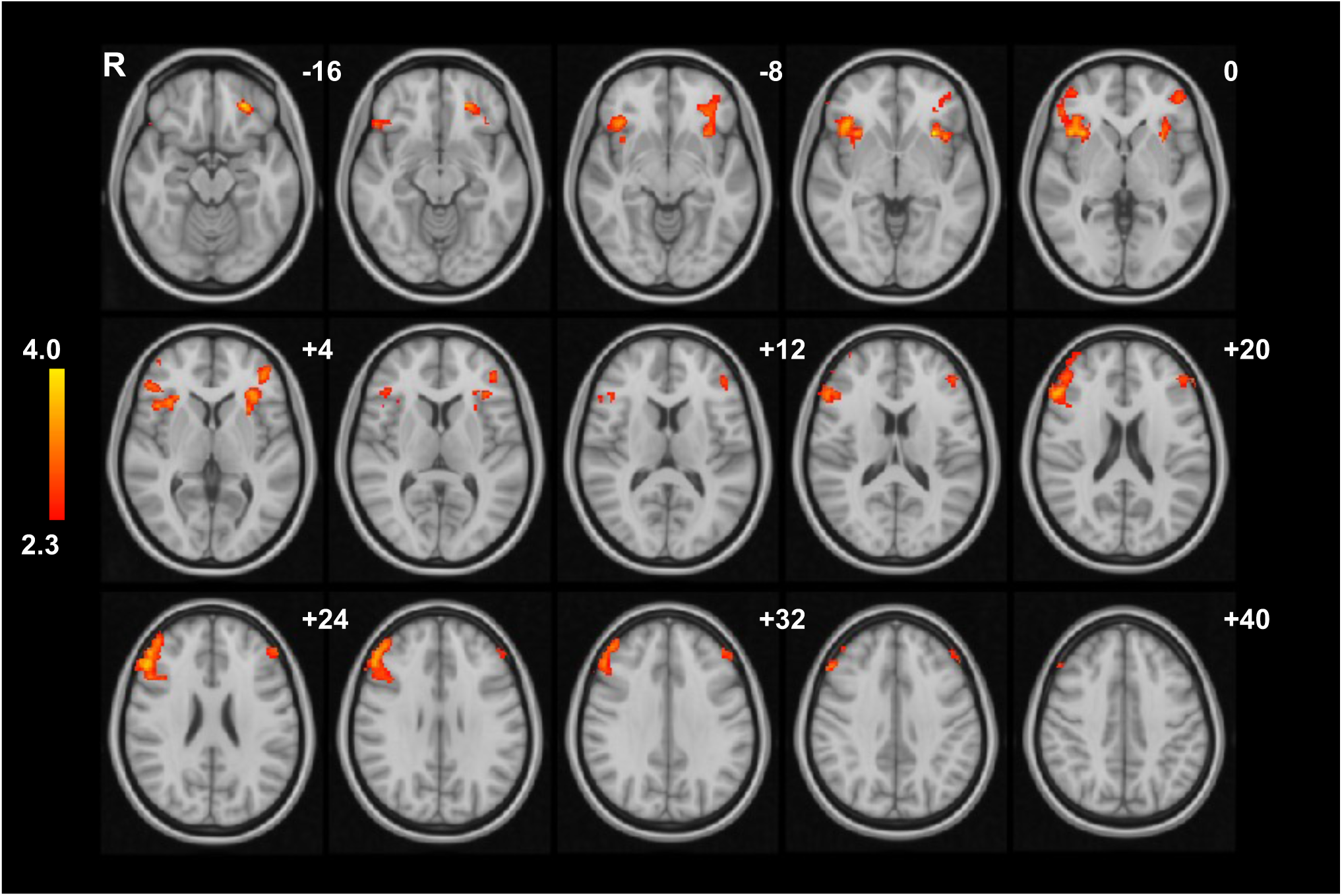
Greater cigarette cue reactivity in ex-smokers compared to obesity from whole brain analysis of picture evaluation fMRI task. Whole brain analysis showing brain regiosn with higher percentage BOLD signal to cigarette pictures (vs. neutral) in ex-smokers (ExS, n=25) than obesity (OB, n= 25) group. Unpaired t-test, cluster-wise threshold Z>2.3, family-wise error (FWE) P<0.05. z co-ordinates given in Montreal Neurological Institute (MNI) space. Abbreviations: R, right. See Supplementary Table S13 for MNI co-ordinates.

#### Cigarette picture appeal rating

The ExS group rated cigarette pictures (vs. neutral) as more appealing than the OB group [U=502.0, P<0.001] (Figure 5).

**Figure 5.**
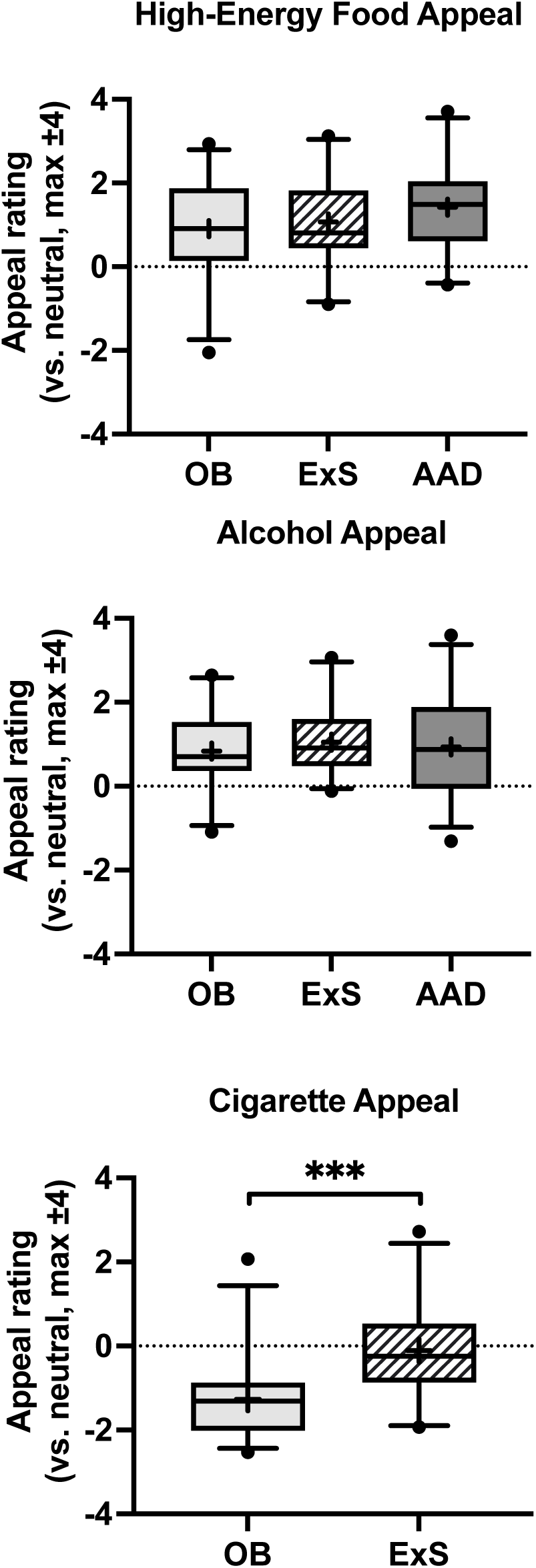
Appeal ratings to high-energy food, preferred alcohol and cigarette pictures. Group comparison of (top) high-energy food, (middle) preferred alcohol and (bottom) and cigarette picture appeal rating (vs. neutral) for adults with obesity (OB, n=25), ex-smokers (ExS, n=25) and abstinent alcohol dependence (AAD, n=26). Data presented as box plot with median (line), mean (+), interquartile range (box) and 5-95^th^ percentiles (error bars), with outliers as a dot. Compared using one-way ANOVA (high-energy food, alcohol), or Mann-U Whitney test (cigarettes) ***P<0.001.

#### Eating behaviour questionnaires

To investigate whether differences in HE food cue reactivity between groups were accompanied by differences in eating behaviour, between group comparisons were made for eating behaviour trait questionnaires. The OB group scored higher than HC but not ExS or AAD groups for DEBQ-restraint and PFS, but not for TFEQ-restraint. The OB group scored higher on DEBQ-emotional eating and BES than HC and ExS groups, but not AAD group. The OB group scored higher than HC, ExS and AAD groups for TFEQ-disinhibition. The OB group scored higher for YFAS than ExS and HC, but not AAD groups, while AAD group scored higher than ExS group. There were no group differences for TFEQ hunger-related eating, or DEBQ-external eating (Supplementary Table S14).

#### Ratings of hunger, appetite, food and drug craving

For hunger VAS ratings, there was a significant difference between groups (P=0.042), with AAD group scoring significantly higher than ExS (P=0.036), but not OB (P=0.51) groups (Table 2).

**Table 2.** Ratings of hunger, appetite and food or drug craving. Data displayed as mean ± SD or median [interquartile range]. Analysed by one-way ANOVA with post-hoc Sidak test for parametric data, or Kruskal-Wallis test with Bonferroni corrected post-hoc test or Mann-Whitney test for non-parametric data. Values are calculated as average across −10 to +315 min using trapezoid rule for area under curve formula. Post-hoc tests: vs. OB ^#^P<0.05, ^##^P<0.01, ^###^P<0.001; vs. ExS ^†^P<0.05, ^††^P<0.01, ^†††^P<0.001.

|  | Obesity (OB) | Ex-smokers (ExS) | Abstinent alcohol dependence (AAD) | Test statistic | P-value |
| --- | --- | --- | --- | --- | --- |
| **n** | 26 | 25 | 25 |  |  |
| **Hunger VAS (score 0-100)** | 50.0 [34.6, 63.9] | 46.4 [16.5, 57.1] | 59.8 [35.3, 76.0] <sup>†</sup> | H(2)=6.35 | **0.042** |
| **Appetite VAS (score 0-100)** | 58.4 [44.0, 66.0] | 58.1 [36.0, 64.3] | 67.2 [53.6, 76.7] <sup>†</sup> | H(2)=7.41 | **0.025** |
| **Food craving (score 2-14)** | 6.71 ± 2.78 | 5.73 ± 2.36 | 7.87 ± 3.32 <sup>†</sup> | F(2,74)=3.59 | **0.033** |
| **Cigarette craving (QSU-brief, score 10-70)** | 10.0 [10.0, 10.1] | 10.0 [10.0, 12.7] | n/a | U=215 | **0.015** |
| **Alcohol craving (AUQ, score 8-56)** | 9.62 [8.32, 14.0] | 16.92 [11.96, 20.00] | 14.81 [10.13, 20.04] | H(2)=3.39 | 0.18 |

Appetite VAS ratings also differed significantly between the groups (P=0.025), with AAD group scoring significantly higher than ExS (P=0.025), but not OB (P=0.17) groups (Table 2).

For food craving, there was a significant difference between groups (P=0.033), with AAD scoring significantly higher than ExS (effect size mean ± SEM 2.14 ± 0.80 (95% CI 0.18, 4.09), P=0.027], but not OB (effect size mean ± SEM 1.13 ± 0.79 (95% CI 0.42, 2.74), P=0.38) groups (Table 2).

For cigarette craving using QSU-brief, ExS scored significantly higher than OB (P=0.015], but average scores were still very low (Table 2).

For alcohol craving using AUQ, there were no significant differences between AAD, ExS and OB groups (P=0.18), with average scores in the lower quartile of possible range for all groups (Table 2).

#### *Ad libitum* lunch taste ratings

For rating of taste liking, there was no significant group*sweet*fat interaction [F(2,73)=0.01, P=0.99], nor fat*group interaction [F(2,73)=0.99, P=0.38]. However, there was a significant sweet*group interaction [F(2,73)=4.43, P=0.015], driven by the AAD group liking the taste of sweet dishes more than the ExS group, but not the OB group (Figure 6, Supplementary Table S15).

**Figure 6.**
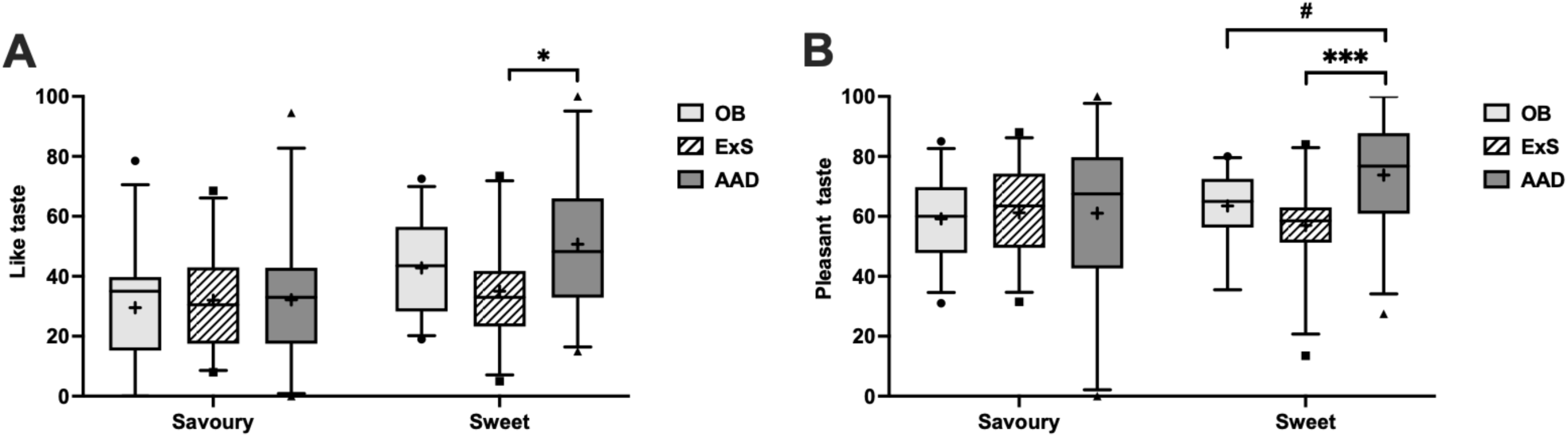
*Ad libitum* lunch taste ratings for liking and pleasantness of savoury and sweet dishes. Group comparison of visual analogue scale ratings of taste (A) liking and (B) pleasantness in adults with obesity (OB, n=25), ex-smokers (ExS, n=25) and abstinent alcohol dependence (AAD, n=26), for savoury and sweet dishes (independent of low/high fat content). Data presented as box plot with median (line), mean (+), interquartile range (box) and 5-95^th^ percentiles (error bars), with outliers as a dot. One-way repeated measures ANOVA with post-hoc LSD test: ^#^ P<0.05, or post-hoc Sidak test: * P<0.05, *** P<0.001 (to control for multiple comparisons). See Supplementary Table S15 for statistical comparisons.

For rating of taste pleasantness, there was no significant group*sweet*fat interaction [F(2,73)=0.45, P=0.64], nor fat*group interaction [F(2,73)=0.28, P=0.75]. However, there was a significant sweet*group interaction [F(2,73)=4.17, P=0.019], driven by the AAD rating sweet dishes as more pleasant than both the ExS and OB groups (Figure 6, Supplementary Table S15).

For creamy taste, there was no significant group*sweet*fat interaction [F(2,72)=0.36, P=0.68]; however there were significant sweet*group [F(2,73)=4.26, P=0.018] and fat*group [F(2,73)=3.93, P=0.024]. interactions. For sweet dishes, the ExS group rated the taste as less creamy than both OB and AAD groups (Supplementary Figure S14, Supplementary Table S15). For high-fat dishes, the ExS group also rated the taste as less creamy than both OB and AAD groups (Supplementary Figure S14, Supplementary Table S15).

For ideal creaminess, there was no significant group*sweet*fat interaction [F(2,73)=0.51, P=0.6], nor fat*group interaction [F(2,73)=2.30, P=0.11], but there was a significant sweet*group interaction [F(2,73)=3.53, P=0.034]. The ExS rated savoury dishes as more ideally creamy than AAD but not OB (Supplementary Figure S14 and Supplementary Table S15).

For sweet dishes only, there was no significant fat*group interaction [F(2,73)=0.43, P=0.65] or effect of group [F(2,73)=2.12, P=0.13]. For ideal sweetness of the sweet dishes only, there was also no significant fat*group interaction [F(2,73)=2.57, P=0.083], nor effect of group [F(2,73)=0.91, P=0.41].

#### Ad libitum lunch

The total kcal consumed during the *ad libitum* meal was significantly different between the groups for both absolute total kcal [H(2)=6.45, P=0.040] and total kcal as % REE [H(2)=6.83, P=0.033]. Post-hoc tests indicated that this was driven by the AAD group (median [IQR] absolute kcal 765.5 [501.1, 1194.6]; kcal %REE 43.3 [36.0, 64.0]) consuming significantly more than the ExS group (median [IQR] absolute kcal 499.0 [413.5, 675.0]; kcal %REE 31.7 [23.3, 38.0]) for both absolute kcal (P=0.033 Bonferroni corrected) and kcal as %REE (P=0.033 Bonferroni corrected).

When examining differences between total kcal consumed as % REE by the types of dish, there was no significant group*sweet*fat interaction [F(2,73)=2.78, P=0.068], nor fat*group interaction [F(2,73)=2.82, P=0.066]. There was however a significant sweet*group interaction [F(2,73)=7.59, P=0.001], driven by the AAD group consuming significantly more kcal as % REE from sweet dishes than bot the ExS [effect size mean ± SEM 10.81 ± 2.67 (95% CI 5.509, 16.11), d=1.15, P<0.001] and OB [effect size mean ± SEM 6.950 ± 2.660 (95% CI 1.648, 12.252), d=0.74, P<0.001] groups.

## DISCUSSION

This experimental medicine fMRI study compared food and salient drug cue reactivity (in a picture evaluation task) and eating behaviours in adults with obesity, ex-smokers and AAD (with the latter two groups having similar average BMI that was lower than the group with obesity), with in summary the following findings.

Recently abstinent ex-smokers exhibited: (i) increased BOLD signal to HE food averaged across the *a priori* fROIs (NAcc, caudate, putamen, anterior insula, OFC, amygdala, hippocampus, vACC and dlPFC) in fROI analysis, and (ii) increased BOLD signal to HE food in the caudate, putamen and anterior insula in whole brain analysis, compared to both the groups with obesity and AAD; however, (iii) this was not accompanied by higher HE food picture appeal rating, hunger and appetite VAS and food craving ratings, or total calorie intake (as % REE) in an *ad libitum* meal in the ex-smokers than the other two groups.

When examining calories consumed from individual dishes, the group with AAD (i) consumed more calories (as % REE) from the sweet dishes compared with both the ex-smokers and adults with obesity. Furthermore, during taste testing, (ii) the group with AAD and obesity both liked sweet dishes more than the ex-smokers, while (iii) the group with AAD rated sweet dishes as more pleasant than the ex-smokers and group with obesity (independent of low/high fat category).

For preferred alcohol cues, there was no significant difference in BOLD signal between the groups in whole brain or fROI analyses during the picture evaluation fMRI task, but unfortunately the task did not elicit reward system activation in any group (i.e. no greater BOLD signal than neutral pictures). There were no group differences in craving for alcohol (AUQ) or preferred alcohol picture appeal ratings.

For cigarette cues, although on average there was no reward system activation during the picture evaluation fMRI task in the ex-smokers, they did exhibit: (i) greater BOLD signal in the MFG, IFG pars opercularis and pars triangularis, frontal operculum cortex, OFC, anterior insula and frontal pole, compared with the group with obesity (never smoked) in whole brain analysis, accompanied by (ii) higher cigarette picture appeal rating in the ex-smokers than the group with obesity. The ex-smokers had higher cigarette craving scores (QSU) and cigarette picture appeal ratings compared with the group with obesity. The group with AAD were a mixture of never, ex-smokers and current smokers and so the lack of differences with other groups was not unexpected.

### Food cue reactivity in ex-smokers

This study hypothesised that ex-smokers would display heightened HE food cue reactivity. We did indeed observe heightened BOLD signal to HE food pictures in recently abstinent ex-smokers on both fROI and whole brain analysis across the following regions: nucleus accumbens, caudate, putamen, anterior insula, OFC, amygdala, hippocampus, ventral anterior cingulate cortex and dorsolateral prefrontal cortex, than group with obesity (all never smokers) and group with AAD. A caveat is that in the AAD group, 46% were current smokers and 42% ex-smokers. However, the individuals with AAD who were ex-smokers had a significantly longer time since quitting than the ex-smokers (without AAD; median 117 and 20.7 weeks respectively). Furthermore, the ex-smokers (without AAD) still displayed greater BOLD signal to HE food than the AAD who were ex-smokers in the anterior insula, OFC, caudate and putamen (see Supplementary Table 11).

Interestingly, this novel finding of heightened food cue reactivity and its distribution in ex-smokers is in keeping with previous fMRI studies of current smokers who demonstrated attenuated food cue reactivity in brain reward systems. Currently smoking adults (average 9 cigarettes per day) displayed lower BOLD signal in the caudate, putamen, insula and thalamus to personalised favourite food spoken words compared with non-smokers (whole brain analysis, n=23 per group) (39). Even low cigarette exposure appears to have a suppressive effect on food cue reactivity, with adolescent smokers (average 1-5 cigarettes per day) exhibiting lower BOLD signal to pleasurable food images in the anterior insula, inferior frontal region and rolandic operculum compared with non-smoking adolescents (whole brain analysis, though only n=12 per group and using uncorrected statistics) (75).

The results of the current paper would be consistent with reversal of this blunting of HE food cue reactivity following smoking cessation, though to our knowledge there are no within participant longitudinal fMRI studies of food cue reactivity. However, an increase in reward value of food was seen at 1 week after sustained smoking abstinence (measured by preference of effort willing to expend for individualised highly palatable snack foods compared to money using a progressive ratio task) in a longitudinal study of smokers (n=36 receiving placebo from a controlled-trial of bupropion for smoking cessation) in those carrying at least one copy of the addiction-risk A1 minor allele in the Taq1A/Ankyrin repeat and kinase domain containing 1 kinase (ANKK1) that influences dopamine receptor D2 (DRD2) function (but not those with two copies of the wild-type A2 allele) (76). Food reinforcement after quitting smoking also predicted increases in weight after 6 months follow-up independent of Taq1A genotype (76).

Difference in food taste reactivity have also been reported in ex-smokers. Current smokers (n=11) exhibited greater BOLD signal when tasting milkshake than healthy controls in the hypothalamus, though not in the midbrain, thalamus and ventral striatum, which were the brain regions in healthy controls where greater BOLD signal positively correlated with greater BMI increase at one year follow-up (40).

The benefits of smoking cessation are numerous and well established, therefore in recent years attention has turned to identifying barriers preventing individuals from stopping smoking. Of these barriers, smoking cessation weight gain is an important issue, with 25-50% of smokers reporting that this is a concern (34,77,78). On average individuals will consume an extra 227 kcal per day following smoking cessation, which is thought to account for 67% of the weight gained in the first 3 months (79) (37). Body fat also increases following smoking cessation (80). However, there is limited evidence for effective long-term dietary and pharmacological strategies to prevent weight gain after smoking cessation (37,81).

In current smokers, there are several proposed mechanisms contributing to nicotine mediated weight loss. Nicotine has anorexigenic effects through stimulation of hypothalamic anorexigenic neuropeptide pathways including pro-opiomelanocortin neurons and also via actions on monoamine systems including serotonin, dopamine and noradrenaline. These act to reduce hunger, increase satiety, alter eating behaviour and increase energy expenditure, while smoking cigarettes also provide a behavioural alternative to eating, all leading to weight loss (37,82,83).

On cessation of smoking these anorexigenic and metabolic effects of nicotine appear to reverse, promoting weight gain (37). The heightened HE food cue reactivity particularly in the caudate, putamen and anterior insula that we observed in ex-smokers (greater than both groups with obesity and AAD), and consistent with previous literature where current smokers had lower favourite food auditory cue reactivity in caudate and putamen than non-smokers (39), may provide a neurobiological basis for smoking cessation increases in food reward and weight gain. Unfortunately, retrospective data on post-quitting weight gain in the ex-smokers was not available in our study. Food cue reactivity in the dorsal striatum may therefore be a useful biomarker to aid the development of novel therapeutic interventions to avoid smoking cessation weight gain.

The caudate, putamen and anterior insula have previously been related to food reward processing. The insula is a primary gustatory centre that plays a role in processing food cues, taste memory and food craving (18,84). Increased BOLD signal in the caudate has previously been reported in response to tasting an energy-dense milk shake compared to a tasteless solution in healthy controls (40), whilst BOLD signal change in the caudate and putamen has been reported in healthy controls during magnitude estimation of taste pleasantness (versus intensity) of a sucrose solution (versus water) (85). Healthy controls with high appetite ratings have greater BOLD signal in the putamen compared with those reporting low appetite (86).

Whilst heightened BOLD signal to HE food pictures was observed in the ex-smokers, this was not accompanied by greater appeal rating of HE food pictures or intake of all or high fat/sweet foods at the *ad libitum* meal (compared to groups with obesity or AAD), nor unhealthier eating behaviours from questionnaires (compared to healthy volunteers, groups with obesity nor AAD). There are several factors that may have contributed to this. With regards to the appeal rating of food pictures, the regions in which we saw heightened BOLD signal to HE food in the ex-smokers may not directly relate to the “appeal” of the HE foods viewed. The OFC has been implicated in food craving, palatability, salience attribution, desire to consume food and hunger (87–89). Using a similar food picture evaluation fMRI task the increase in BOLD signal in the OFC to HE vs. low-energy (LE) foods when fasted (vs. fed) positively correlated with the fasting-induced increase in appeal rating for HE vs. LE food (45). However, the OFC was not a discrete brain region in which enhanced BOLD signal to HE food was seen in the current study of ex-smokers (although it was one of the regions in out ROI analysis), nor in which attenuated food cue reactivity was seen in the previous fMRI studies of current smokers (39,75).

With regards to the *ad libitum* lunch intake, only four dishes were presented of which only half were HE foods which may have not been preferred by all participants (unlike the food cue reactivity fMRI task which consisted of 60 different HE food pictures), as indicated by the high variability in liking and pleasantness taste ratings and intake of the dishes. Only the ice cream scored on average above 40/100 in taste liking. In addition, the participants were in a hungry state having only had 148 kcal snacks at 350 mins and 105 mins before the meal, which may have led to ceiling effects, limiting the ability to detect greater food intake in the ex-smokers. Furthermore, the dishes were served a meal in a laboratory rather than ecological setting, impairing our ability to detect real world differences in energy intake.

### Sweet taste endophenotype in alcohol use disorder

The finding that the group with AAD both consumed more calories from the sweet dishes (independent of fat content) in the *ad libitum* meal, and rated taste liking and pleasantness of sweet dishes as higher, than the ex-smokers, and/or group with obesity is consistent with increased sweet taste hedonics being an underlying endophenotype predisposing to AUD (49,50). In partial agreement with our findings, preference for sweet taste in AUD has been seen in early abstinence (<1 month), though not at 6 months, while those with sweet-liking AUD were less likely to remain abstinent at 6 months (51). Furthermore, the proportion of sweet-likers in AUD was increased if there was a paternal history of AUD (77.3 vs. 47.8%) (53), and brain responses to oral sucrose differed with a family history of AUD (90). However, in other studies there were no differences in sweet taste pleasantness or intensity ratings or proportion of sweet likers in AUD (abstinence length mean 40 days, range 6–105) (52), nor sweet taste pleasantness or intensity ratings in sons of men with AUD (91). However, unlike the current study, none of these previous studies examined preference for actual intake of sweet foods, which we found to be increased in AAD. Overall, these findings are supportive of increased sweet taste hedonics being a potential underlying endophenotype predisposing to AUD. Indeed, wild chimpanzees who eat large amounts of (sweet) fruits (∼4.5kg daily) have the equivalent alcohol ingestion of 14g ethanol per day, due to yeast fermentation of carbohydrates, providing a potential evolutionary link between intake of sweet foods and alcohol (92).

Further support for this conclusion and a potential mechanistic link between sweet and alcohol taste/intake, comes from the findings that the liver-derived hormone fibroblast growth factor 21 (FGF21), that acts on the FGF receptor and its obligate co-receptor β-Klotho (*KLB* gene), is an inhibitor of both alcohol and sugar intake in mice and non-human primates (93–97), and single nucleotide polymorphisms in the *FGF21* and *KLB* genes are associated with increased consumption of alcohol, sweets and carbohydrate (98–101). Plasma FGF21 increases after acute ingestion of alcohol and sugar including in AUD, and sustained binge drinking (98,102–104). However, to date there are no reports in humans of effects of FGF21 or analogues on sweet or alcohol taste or intake in AUD.

Given the caloric content of alcohol, energy intake will decrease after abstaining from alcohol in AUD, at least in the short term (105). It is currently unclear if the underlying sweet preferring phenotype in AUD, might predispose to longer-term greater intake of sweet foods in compensation, and hence predisposition to weight maintenance and even weight gain despite abstinence from alcohol.

### Alcohol cue reactivity and craving

The lack of difference in preferred alcohol cue-reactivity and alcohol craving in AAD compared to the other clinical groups may be related to several factors. Firstly, the comparison groups were obesity and ex-smokers without AUD rather than a BMI-matched and smoking-matched group (though 42.3% of group AAD were ex-smokers). Secondly, the current study examined ‘successful’ abstainers who had been in stable abstinence for on average eight months, and with longer-term abstinence, there may be recovery of neural networks in response to alcohol cues. Previous studies demonstrating greater alcohol cue reactivity in AAD have been in earlier abstinence. For example, in a meta-analysis of 28 studies (n=679) heavy drinkers and individuals with AUD at less than one month abstinence had greater alcohol cue reactivity in precuneus, PCC and superior temporal gyrus than healthy controls (106). A meta-analysis of 21 studies (n=795) reported greater alcohol cue reactivity in ACC, middle cingulate gyrus and mPFC in AUD at less than one month abstinence than healthy controls (16). In AUD abstinent for average seven weeks (n=10), there was greater alcohol cue reactivity in putamen, ACC and mPFC, than in healthy controls (107). It is estimated that at six months, approximately 60% of abstinent AUD individuals will have already relapsed, highlighting that the individuals recruited in the current study are likely to maintain their abstinence compared with the wider AUD population (108). Consistent with this interpretation, the group with AAD had similarly low alcohol craving (AUQ) and preferred alcohol appeal ratings than the ex-smokers and adults with obesity, indicating low desire for alcohol in the abstinent AAD group.

Alcohol cue reactivity in the current study was observed only in the precuneus and PCC in the AAD, ex-smokers and adults with obesity, regions that are part of the default mode network. This network is more active during rest and deactivated during attention, and thus this may represent the individuals being disinterested in the alcohol pictures across the three groups.

### Cigarette cue reactivity and craving

In the current study, ex-smokers at average eight months abstinence had greater cigarette cue reactivity than adults with obesity (never-smokers) in multiple areas of the anterior insula and prefrontal cortex (MFG, IFG including right dlPFC (Brodmann area 9/46V), frontal opercular cortex, OFC and frontal pole), though not the striatum in whole brain analysis, and ex-smokers rated the cigarette pictures as more appealing and craved cigarettes (QSU) more than adults with obesity.

Several of these brain regions overlap with those found in previous studies of group differences and factors modulating neural reactivity to smoking cues (109). In a meta-analysis of current smokers (816 participants from 28 studies), there was heightened cigarette cue reactivity in the MFG and OFC compared with healthy controls (217 participants from 13 studies) (15). Fagerstrom Test of Nicotine Dependence has been positively correlated with smoking cue-induced reactivity in OFC and anterior cingulate in current smokers (n=30) (110). Fifty-two days after smoking cessation (n=12), cigarette cue reactivity was still seen in several areas including the MFG and the anterior and posterior insula, though only the former was seen before quitting in dependent smokers (111). In female smokers (n=21), eight weeks abstinence reduced cigarette cue reactivity in the prefrontal cortex including IFG and dlPFC, while individuals who “slipped” (smoked greater than one cigarette during an eight week abstinence period, but no full relapse) had heightened cigarette cue reactivity in IFG, MFG, anterior and posterior insula compared with those who managed to stay abstinent (111).

Heightened cigarette cue reactivity in the PFC in current smokers has also been linked to later relapse after subsequent quit attempts. In current smokers, heightened BOLD responses for cigarette (vs. erotic/romantic) pictures in the mPFC and dlPFC was observed in those who relapsed at six months compared with those who remained abstinent (112).

Of particular interest, the insula is thought to integrate interoceptive states into conscious feelings and is therefore thought to play an important role in the “urge to use” drugs (113). Smokers with insula damage from cerebrovascular accidents are more likely to quit smoking immediately, without the persisting urge to smoke and without relapse, compared with smokers who do not have insula damage, highlighting the potentially critical role of the insula in maintaining nicotine use disorder (113).

Ongoing reactivity to cigarette cues of the PFC and anterior insula in even longer-term ex-smokers as in the current study might confer an increased relapse vulnerability. The GHADD study had planned to collect clinical outcome data to assess relapse in the ex-smokers one year later, however this was not possible due to interruption to data collection from the COVID-19 pandemic and lockdown.

In the current study, exposure to smoking pictures did not increase cigarette craving (QSU) in ex-smokers, however despite >20 weeks abstinence, ex-smokers had greater cigarette craving and rated the cigarette pictures as more appealing than never-smokers (adults with obesity). This is consistent with previous studies indicating that cigarette craving can persist in long-term in ex-smokers. For example, 59% of ex-smokers with an abstinence range of 1-5 years, reported a desire to smoke in the past year, with 11% reporting “clinically significant” craving (114). However, the importance of persistent cigarette craving after quitting to increase relapse vulnerability remains uncertain.

#### Strengths and Limitations

This was a comprehensive phenotyping and functional neuroimaging study that provides novel comparison of the underlying neurobiology, including assessments of eating and addictive behaviours, between obesity, ex-smokers and abstinent AUD, and all participants in the AAD group had severe AUD by DSM-V criteria.

There are some limitations to consider. The sample size was only moderate for fMRI and behavioural studies at n=24-26 per group, and so there is a risk of both type 1 and 2 errors, including for the null findings for between group differences in alcohol and smoking cue reactivity. Determining statistical power can be difficult for fMRI analyses, especially as the alcohol and smoking cue reactivity fMRI task did not particularly engage brain reward systems in the relevant groups. The findings should be taken as preliminary and in need of confirmation with larger sample sizes.

For interpretation of the HE food cue reactivity task, a major caveat is that due to the difference in nutritional state between the three clinical groups and the healthy controls without obesity or substance dependence, the latter were not a suitable group for direct comparison. Recent meta-analyses have concluded that adults with obesity or higher BMI generally have no differences in HE food cue reactivity than those without obesity or lower BMI, though sometimes heightened reward system responses to HE food pictures are seen (19–22). One would therefore expect that if a control group without obesity had been included in this study, they would display either similar or perhaps lower BOLD responses to HE food cues compared with the adults with obesity. Neither result would affect the interpretation of the finding of heightened HE food cue reactivity in ex-smokers. BMI was also quite variable within the group with obesity which may have influenced the food cue reactivity outcomes, but exploratory analysis did not reveal any significant influence of BMI on BOLD signal to HE food or picture appeal in that group.

The group with obesity reported self-directed active dieting, none of whom were currently participating in a structured weight management programme. Therefore, there will have been differing degrees of dietary restraint and caloric restriction between individuals which may have increased variability in outcome measures. Indeed, participation in a structured commercial or clinical weight management programme with a supervised low-calorie diet or meal replacement programme, with different levels of psychological input, may have produced greater weight loss or changes in food selection, and produced different changes in HE food cue reactivity (115–119). Furthermore, documented retrospective recent weight change data was unavailable and so it could not be determined with any accuracy how much weight had been lost in the group with obesity through their dieting.

Although the group with obesity displayed significantly higher dietary restraint, Power of Food Scale, emotional eating, disinhibited eating and BES scores than the healthy controls, only disinhibited eating, YFAS and BES scores were significantly higher than the ex-smokers, and there was heterogeneity within the groups. Given that HE food cue reactivity can also be influenced by underlying eating behaviour traits (118,120–123), this may also have contributed to the lack of heightened food cue reactivity in obesity compared to the ex-smokers and AAD groups. The healthy control group were younger than the three clinical groups, which may affect interpretation of the eating behaviour questionnaire results.

Similarly, there had been no measurement in the ex-smokers as to how much weight had been gained after stopping smoking, nor in the AAD group after abstaining from alcohol. Future studies would benefit from performing prospective assessments of food cue reactivity using fMRI, as well as food hedonics and intake, prior to and shortly after quitting smoking in AUD, and abstaining from alcohol in AUD, to see if they predict longitudinal weight gain, which will help inform weight management strategies especially in smoking cessation programmes.

The preferred alcohol picture evaluation fMRI paradigm did not elicit reward system activation in the AAD group, and the QSU and AUQ craving scores were on average low in the ex-smokers and AAD groups respectively. This likely represents groups who had successfully maintained abstinence for on average 4.8 and 7.5 months respectively after quitting smoking and alcohol. Future studies should examine these groups at early time points after quitting. Furthermore, the majority of the AAD group were a mixture of current and ex-smokers, albeit for the latter having been successfully abstinent for average 2.3 years, which may complicate direct comparison with the ex-smokers who had been abstinent for on average 4.8 months. However, finding adults with alcohol dependence who have never smoked (as well as having no other history of substance dependence) for such experimental medicine research studies is practically difficult.

Similarly, the lack of cue task elicited craving reported for the cigarette and preferred alcohol cue reactivity paradigms should also be interpreted with caution. Craving was assessed before and after the MRI scan which lasted 1.5 hours, with the cue task close to the start and lasting only 20 minutes. This was followed by several sequences and fMRI tasks, including a final negative emotional reactivity fMRI task, which may have interfered with craving. A more reliable measurement of the effect of the tasks on craving would be to obtain craving scores directly before and after the cue tasks whilst in the scanner. However, appeal ratings for the cigarette and alcohol pictures in the ex-smokers and abstinent AUD groups were generally low.

The *ad libitum* meal consisted of a limited selection of two soups, yoghurt and ice cream, which had a similar semi-liquid consistency. This is a selection that may not adequately capture typical eating behaviour in obesity, where there is often consumption of a wider variety of solid, hyper-palatable foods. However, we have found appropriate decreases in consumption of these foods at an *ad libitum* meal in adults with obesity after Roux-en-Y gastric bypass surgery (unpublished observations). Nevertheless, the meal design may not have been appropriate for detecting meaningful group differences in energy intake. This might explain why the ex-smokers had heightened HE food cue reactivity, but did not display greater sweet, high-fat food intake at the *ad libitum* meal (although the AAD group did display both heightened taste hedonics and intake of sweet foods). Future studies should consider using a greater variety of dishes at *ad libitum* meals, or consider using a virtual computer-based portion size creation task that can easily present a number of different foods (124,125).

## Conclusion

The important conclusions from this study were that: (i) ex-smokers display heightened cigarette cue reactivity compared with never-smokers with obesity, mirroring brain responses observed in current smokers, that may represent potential pharmacological/behavioural targets for treatment in NUD; (ii) ex-smokers display heightened HE food cue reactivity in dorsal striatum compared with adults with obesity who had never smoked, and AAD who were mainly either current smokers or been longer-term abstinent from cigarettes, indicating a potential mechanism for smoking cessation weight gain, and a neuroimaging biomarker for pharmacological/behavioural interventions that target post-quitting weight management, which may also help with smoking cessation, such as the use of GLP-1 analogues (126–128); (iii) individuals with severe AUD who are abstinent display heightened sweet taste hedonics and intake compared with ex-smokers and adults with obesity, providing additional evidence of sweet taste preference being an underlying endophenotype predisposing to AUD, which may open up novel avenues for treatment; (iv) unlike with cigarettes cue reactivity in ex-smokers, heightened preferred alcohol cue reactivity was not seen in AAD after medium-term abstinence, perhaps due to normalisation over time.

## Data Availability

All data produced in the present study are available upon reasonable request to the authors

## Abbreviations

AAD: abstinent alcohol use disorder
AUD: alcohol use disorder
BOLD: blood oxygen level dependent signal
dlPFC: dorsolateral prefrontal cortex
ExS: ex-smokers
fMRI: functional magnetic resonance imaging
fROI: functional region of interest
FWE: family-wise error
HC: healthy controls
Ins: insula
NAcc: nucleus accumbens
NUD: nicotine use disorder
OB: obesity
OFC: orbitofrontal cortex
ROI: region of interest
vACC: ventral anterior cingulate cortex.

## ACKNOWLEDGMENTS

The authors thank the UK Research and Innovation (UKRI) for funding the study via a Medical Research Council (MRC) Experimental Medicine Challenge grant (MR/M007022/1). We acknowledge assistance from Invicro staff (Matt Wall, Mari Lambrechts, James Davies, Mark Tanner, Jonathan Howard, Yvonne Shearley) for MRI scanning; research nurses (Barbara Kobson, Felicity Aiano, Silvia Moreira, Rebecca Mswelli, Emmima Manoharan, Jake Dagen); National Institute for Health Research (NIHR) Imperial Clinical Research Facility staff for support; Central London Community Healthcare NHS Trust for recruitment. Infrastructure support was provided by the NIHR Imperial Biomedical Research Centre and the NIHR Imperial Clinical Research Facility. The views expressed are those of the author(s) and not necessarily those of the National Health Service (NHS), NIHR or Department of Health and Social Care (DHSC).

For the purpose of open access, the authors have applied a Creative Commons Attribution (CC-BY) licence to any author accepted manuscript version arising.

