## Supplementary Material for "Salient cue reactivity and eating behaviours in ex-smokers, abstinent alcohol use disorder and obesity"

Table S13. Spatial coordinates from whole brain analysis for cigarette cue reactivity during picture evaluation fMRI task between ex-smokers and obesity. 30
Table S14. Eating behaviour questionnaires. 31
Table S15. *Ad libitum* lunch taste ratings. 32
Table S16. Spatial coordinates from exploratory whole brain analysis for cigarette cue reactivity during picture evaluation fMRI task between ex-smokers without and with alcohol dependence. 34
**SUPPLEMENTARY FIGURES 35**
Figure S1. GHADD study day protocol. 35
Figure S2. Functional regions of interest for picture evaluation fMRI task. 36
Figure S3. Whole brain analysis of high-energy food cue reactivity from picture evaluation fMRI task in obesity. 37
Figure S4. Whole brain analysis of high-energy food cue reactivity from picture evaluation fMRI task in ex-smokers. 38
Figure S5. Whole brain analysis of high-energy food cue reactivity from picture evaluation fMRI task in abstinent alcohol dependence. 39
Figure S6. Whole brain analysis of high-energy food cue reactivity from picture evaluation fMRI task for comparison between ex-smokers and other groups. 40
Figure S7. Group differences in alcohol cue reactivity from functional region of interest analysis. 41
Figure S8. Whole brain analysis of alcohol cue reactivity from picture evaluation fMRI task in obesity. 42
Figure S9. Whole brain analysis of alcohol cue reactivity from picture evaluation fMRI task in ex-smokers. 43
Figure S10. Whole brain analysis of alcohol cue reactivity from picture evaluation fMRI task in abstinent alcohol dependence. 44
Figure S11. Group differences in cigarette cue reactivity from functional region of interest analysis. 45
Figure S12. Whole brain analysis of cigarette cue reactivity from picture evaluation fMRI task in obesity. 46
Figure S13. Whole brain analysis of cigarette cue reactivity from picture evaluation fMRI task in ex-smokers. 47
Figure S14. *Ad libitum* lunch taste ratings for creamy taste and ideal creaminess. 48
**SUPPLEMENTARY REFERENCES 49**

**SUPPLEMENTARY METHODS**

**Participant inclusion criteria**

1. Male or female volunteers between the ages of 18 and 60 years.
2. Healthy as determined by a responsible physician, based on a medical evaluation including medical history, physical examination, laboratory tests, cardiac monitoring and a psychiatric evaluation. Any volunteer with a clinical abnormality or laboratory parameters outside the reference range for the population being studied may be included, only if the investigators concur that the finding is unlikely to jeopardize either volunteer safety or study integrity. With regards to liver function, see exclusion criterion 16.
3. Participant is capable of giving written informed consent, which includes compliance with the requirements and restrictions listed in the consent form.
4. Participant is able to read, comprehend and record information written in English.
5. A signed and dated written informed consent is obtained from the participant.
6. For groups without history of substance dependence:
   (i) Overweight/obesity with BMI 28.0-50.0 kg/$m^2$ who self-reported as actively dieting.
   (ii) Healthy volunteers for pilot testing of main protocol with BMI 18.0-35.0 kg/$m^2$.
7. For groups with history of substance-dependence:
   (i) Abstinent alcohol dependent individuals who meet DSM-V criteria for previous alcohol dependence (moderate-severe alcohol use disorder), but who are in stable abstinence. Minor lapses will be allowed but not relapses into dependence.
   (ii) Abstinent tobacco dependent individuals who meeting DSM-V criteria for previous nicotine dependence, but who are in stable abstinence. Minor lapses will be allowed but not relapses into dependence. When smoking subjects must have had their first cigarette within 60 minutes of waking and have smoked at least 5 cigarettes per day, as measured retrospectively using the Fagerström Test for Nicotine Dependence (FTND) (Heatherton et al., 1991), and who have been in stable tobacco abstinence for at least 6 weeks.

**Participant exclusion criteria**

Potential volunteers will NOT be eligible for inclusion in this study if any of the following criteria apply:

1. Previous history of recreational use or abuse of other substances of addiction will be permissible, but there should be no use of any illegal drugs (except cannabis) in the month prior to the Screening Visit or during the course of the study, except where specified for individual groups below.

2. For individual groups:
   (i) Group with overweight/obesity and healthy volunteers: history of or current alcohol abuse or dependence; nicotine use other than "never smoked", i.e. >100 cigarettes lifetime use; history of

dependence, abuse or heavy recreational use of cocaine, cannabis, opiates or other substance of abuse; history of problem gambling. Any previous or current psychiatric diagnosis listed in DSM-V Axis I, which in the opinion of the clinical team will compromise conduct and interpretability of the study. Participants currently suffering from DSM-V depressive disorder or on anti-depressant medication will be excluded, though a previous history of depression will be allowed. Screening for Axis I psychiatric diagnoses will be performed using a summarized version of the Mini International Neuropsychiatric Interview for Mini International Neuropsychiatric Interview for DSM-V (MINI). This interview will be performed by appropriately trained study personnel. The MINI is a short, structured interview requiring "yes" or "no" answers only.

(ii) Group with abstinent tobacco dependence: history of or current alcohol abuse or dependence; current dependence for cocaine, cannabis, opiates or other substance of abuse, or problem gambling (previous history will be allowed); taking varenicline, bupropion or other prescription medications for smoking cessation. Any previous or current psychiatric diagnosis listed in DSM-V Axis I, which in the opinion of the clinical team will compromise conduct and interpretability of the study. Screening for Axis I psychiatric diagnoses will be performed using the MINI Interview for DSM-V. Appropriately trained study personnel will perform this interview.

(iii) Group with abstinent alcohol dependence: current dependence for cocaine, cannabis, opiates or other substance of abuse, or problem gambling (previous history will be allowed); taking prescription medication for alcohol or smoking cessation or withdrawal; smoking is allowed past or present including dependence; current nicotine replacement therapy is allowed. Any previous or current psychiatric diagnosis listed in DSM-V Axis I, which in the opinion of the clinical team will compromise conduct and interpretability of the study. Screening for Axis I psychiatric diagnoses will be performed using the MINI Interview for DSM-V. Appropriately trained study personnel will perform this interview.

3. A current or past history of enduring severe mental illness (e.g., schizophrenia, bipolar affective disorder) will not be allowed. Screening for Axis I psychiatric diagnoses will be performed using the MINI Interview for DSM-V. This interview will be performed by appropriately trained study personnel.

For all groups:

4. Cannabis use up to five times in the month prior to the Screening visit will be allowed, but no use within one week of experimental assessments; no use of any other illegal drugs in the month prior to the Screening Visit or during the course of the study.
5. Intoxication at any of the visits, as manifested by difficulty in walking, slurring of speech, difficulty concentrating or drowsiness (or by the subject volunteering this information directly to the research team). This exclusion criterion would exclude a volunteer from that study day only and not the whole study, at the discretion of the research team.

6. Positive drug/alcohol screens on testing at the screening visit, other than that explicable by other causes (e.g. recent use of opiate containing analgesic, consumption of poppy seeds for positive opiate screen), at the discretion of the research team. A minimum list of drugs that will be screened for include amphetamines, barbiturates, cocaine, opiates, cannabinoids and benzodiazepines. Positive results for cannabinoids will be allowed given the long half-life of cannabinoid metabolites. If positive at a study visit this exclusion criterion would exclude a subject from that study day only and not the whole study, at the discretion of the research team.
7. The British National Institute for Clinical Excellence (NICE) stipulates that a non-smoker is identified by a breath carbon monoxide reading of less than 10ppm (NHS Stop Smoking Services; Service and Monitoring Guidance 2010/11). Therefore, carbon monoxide levels of =/>10ppm in the healthy volunteer, overweight/obesity and abstinent smoker groups at the screening visit will result in exclusion. If positive at a study visit this would exclude a subject from that study day only and not the whole study, at the discretion of the research team.
8. Use of current regular prescriptions (including smoking or alcohol cessation medicines such as Disulfiram, Acamprosate, Naltrexone, Bupropion; weight loss medication including Orlistat, Metformin, GLP-1 agonists, Bupropion, Naltrexone), or over-the-counter medications that in the opinion of the Investigators may affect subject safety or outcome measures.
9. Pulse rate <40 or >100 beats per minute OR systolic blood pressure >160 and <100 and a diastolic blood pressure >95 and <50 in the semi-supine position.
10. Claustrophobia or feels that they will be unable to lay still on their back in the MRI scanner for a period of ~80 minutes.
11. Presence of a cardiac pacemaker or other electronic device or ferromagnetic metal foreign bodies as assessed by a standard pre-MRI questionnaire and radiographer.
12. History or presence of a neurological diagnosis (not limited to but including, for example, stroke, epilepsy, space occupying lesions, multiple sclerosis, Parkinson's disease, vascular dementia, transient ischemic attack, that may influence the outcome or analysis of the scan results). A history of alcohol related or alcohol-withdrawal seizures will be allowed for volunteers in the abstinent alcoholic group.
13. Significant current or past medical or psychiatric history that, in the opinion of the investigators, contraindicates their participation. Screening for Axis I psychiatric diagnoses will be performed using the MINI Interview for DSM-V.
14. Clinically significant head injury (e.g. requiring hospitalisation or surgical intervention) that in the opinion of the investigators may affect subject safety or outcome measures.
15. Unwillingness or inability to follow the procedures outlined in the protocol.
16. Any of the following liver function tests (LFT) abnormalities at screening: alkaline phosphatase, AST, ALT or gammaGT > 4 x upper limit of normal (ULN), INR > 1.5, Albumin <25 g/L, raised bilirubin (other than just isolated i.e. without other liver function tests abnormalities).

17. History of decompensated alcoholic liver disease, i.e. history of variceal bleeding, ascites, jaundice, encephalopathy.
18. History of pancreatitis from any cause.
19. History of type 1 or type 2 diabetes mellitus.
20. ECG abnormality, which in the opinion of the study physician, is clinically significant and represents a safety risk.
21. The volunteer has participated in a clinical trial and has received an investigational product within the following time period prior to the first experimental visit in the current study: 90 days, 5 half-lives or twice the duration of the biological effect of the investigational product (whichever is longer).
22. Exposure to more than 3 new investigational medicinal products within 12 months prior to the scan.
23. History of sensitivity to any of the peptides, or components thereof, or a history of drug or other allergy that, in the opinion of the investigators, contraindicates their participation.
24. Diagnosis of endocrine disorder, including uncontrolled hypothyroidism (stable treated hypothyroidism with currently normal thyroid function tests is allowed), history of hyperthyroidism or Cushing's syndrome, which, in the opinion of the investigators, may affect subject safety or outcome measures.
25. History of ischaemic heart disease, heart failure, cardiac arrhythmia or peripheral vascular or cerebrovascular disease.
26. History or presence of significant respiratory, gastrointestinal, hepatic, oncological or renal disease or other condition that in the opinion of the Investigators may affect subject safety or outcome measures.
27. Previous bariatric surgery for obesity including Roux-en-Y gastric bypass, gastric banding, sleeve gastrectomy.
28. Current pregnancy or breast-feeding in female volunteers (the Investigators would advise on using contraception for the duration of the visits).
29. Vegetarian, vegan, gluten or lactose-intolerant (as food pictures and test meals in the paradigms include meat, dairy and wheat products).
30. Volunteers who have donated, or intend to donate, blood within three months before the screening visit or following study visit completion.

**Study protocol**

Participants attended one screening visit and three experimental study visits. In addition to confirming eligibility on screening, they were familiarised with the experimental procedures (including food, cigarette and preferred alcohol picture viewing to minimise any habituation effect) and completed several computer-based questionnaires and neurocognitive tasks. For the purpose of this paper results from the Wechsler Test of Adult Reading (WTAR) (Wechsler 2001), Beck's Depression Inventory (BDI-II) (Beck et al. 1996) and Alcohol Use Disorders Identification Test (AUDIT) (Saunders et al. 1993) are described.

Following screening participants attended three study visits separated by at least five days in which they received intravenous infusions of either placebo (using diluent for peptide hormones, 0.5% human albumin solution in 0.9% normal saline), des-acyl ghrelin (4.0 mcg/kg/hr, Clinalfa, Bachem) or Exenatide (Exendin-4, 0.015 mcg/kg/hr, Byetta, AstraZeneca) in a double blinded, randomised cross-over design. For this analysis only results from the placebo saline infusion visit are reported. The HC group did not receive any infusions.

Prior to each experimental visit, participants in the clinical groups (OB, ExS, AAD) were asked to fast from 20:00 hours the night before and to avoid exercise and alcohol intake the day before and morning of the study visit. They arrived fasted to the In Vicro Clinical Imaging Facility, Hammersmith Hospital, London, United Kingdom, and received a 148-kcal snack (T=-25 min pre-infusion), 130 min before the MRI scanning session. The HC group had their usual breakfast at home on the morning of the study visit.

All participants were breathalysed for alcohol and carbon monoxide (to determine abstinence from smoking for HC, OB and ExS groups), urine tested for illicit substances and had measurements of height and weight taken. Pregnancy was excluded using a urinary human chorionic gonadotropin test and antecubital vein cannulas were inserted for the infusion (started at T=0 min) and serial blood sampling. At the end of each visit, participants had body fat measured by bio-electrical impedance analysis (Tanita Europe BV, Amsterdam, The Netherlands).

**Picture evaluation fMRI paradigm**

At T= 115min (~12noon), fMRI scans were performed including a practice run of picture evaluation task of neutral and animal pictures, followed by the actual task using an MRI-compatible button box. This fMRI picture evaluation task was based on protocols of previous studies (Goldstone et al. 2014, Scholtz et al. 2014, Byrne et al. 2016, Goldstone et al. 2016). In the picture evaluation task, four types of colour photographs were presented in a block design split across two 10 minute runs: (i) 60 high-energy (HE) food (e.g. pizza, cake, chocolate), (ii) 60 cigarette (e.g. lit cigarettes, people holding a cigarette), (iii) 60 personalised alcohol to two preferred types of alcoholic drinks (e.g. beer, wine, spirits) and (iv) 60 matched control neutral images (e.g. containing hands, faces, objects). Participants simultaneously rated the appeal of individual images with the 5-button box (1 = not at all, 2 = not really, 3 = neutral, 4 = a little, 5 = a lot).

These images were presented in blocks of 18 seconds each and separated by rest periods of 12 seconds (software: E-prime, Psychology Software Tool Inc, Pittsburgh, PA, USA). Images were viewed via a mirror mounted above an 8 channel radiofrequency head coil which displayed images from a projector using the E-Prime 2 Professional software. Each participant viewed a total of 60 images in each of the four categories across ten blocks by using a pseudorandomised block order (randomised for each subject across study visits) with a randomised picture order within each block. Every image was displayed for 2500ms and followed by an interstimulus fixation cross interval of 500ms. The images were of similar luminosity and resolution.

**MRI scanning parameters**

Whole brain fMRI data was acquired on a 3 Tesla Siemens Verio MRI scanner at the In Vicro Clinical Imaging Facility, Hammersmith Hospital, London, UK using a 32 channel radiofrequency head coil with T2* weighted gradient-echo echoplanar imaging (EPI) with an automated higher-order shim procedure: 54 ascending contiguous 3.0mm thick slices, 3.0 x 3.0 mm voxels, multiband 2, GRAPPA acceleration factor 2 repetition time (TR) 1500ms, echo time (TE) 30ms, 80o flip angle; field of view (FOV) 192mm, slice acquisition angle parallel to anterior-posterior commissure line. 414 volumes were collected over 10.35 mins for each of the 2 runs.

High-resolution T1-weighted MPRAGE structural scans were also collected for image registration (TE 2.98 ms, TR 2300 ms, flip angle 9o, FOV 256 mm, voxel dimensions 1.0 x 1.0 x 1.0 mm). Field maps were used to correct for geometric distortions caused by inhomogeneities in the magnetic field as follows (taken from FMRI Expert Analysis Tool (FEAT) analysis software output). Structural T2-weighted scans were also acquired to identify any structural abnormalities.

**Image pre-processing**

The following pre-processing steps were applied: motion correction by using MCFLIRT software, field-map-based EPI unwarping by using PRELUDE and FUGUE software, non-brain removal by using BET software, spatial smoothing by using a Gaussian kernel of full-width half-maximum (FWHM) 6.0 mm, temporal smoothing by high-pass temporal filtering (Gaussian-weighted least-squares straight line fitting, with sigma 45.0s).

Time-series statistical analysis was carried out by using FILM software with local autocorrelation correction including picture onsets convolved with the gamma hemodynamic response function as explanatory variables within the context general linear model (GLM) on a voxel-by-voxel basis, including motion parameters as a covariate of no interest. Registration to high-resolution T1 structural images was carried out by using FLIRT software using boundary-based registration (BBR). Registration from high-resolution structural to standard spaces was then further refined by using FNIRT non-linear registration software.

**Functional region of interest analysis**

In a separate cohort of healthy volunteers without obesity, a whole brain mixed effects analysis using FEAT software v.6.0 was performed for the contrast of HE food or alcohol > neutral pictures. Correction for multiple comparisons was made using a cluster threshold $Z>3.1$, family-wise error (FWE) $P<0.05$.

The fROIs were then created by masking the clusters identified from the group activation map with the *a priori* anatomical regions listed above using fslmaths software within FSL. The fROIs were defined by the relevant bilateral ROIs from the cortical and subcortical structural Harvard FSL atlas (https://fsl.fmrib.ox.ac.uk/fsl/fslwiki/Atlases) except the dorsolateral prefrontal cortex (dlPFC) was defined using the Sallet atlas (Brodmann areas BA9 and BA48). The nucleus accumbens (NAcc) and caudate were thresholded at 50% probability to avoid voxel overlap, while all other areas were thresholded at 10% probability. The OFC fROI included regions in the anterior OFC and frontal pole with y > 24 because the functional activation in this region showed distinct bilateral clusters that overlapped these two anatomical Harvard atlas regions. The insula (Ins) fROI was subdivided into the anterior insula (y > -2) and the anterior cingulate cortex (ACC) subdivided into the ventral ACC (vACC) (y > 30). Coordinates of the fROIs are given in Supplementary Table S4 and visualised in Supplementary Figure S2.

**Exploratory analysis of BMI on food cue reactivity in obesity**

To examine the influence of BMI on HE food cue reactivity in the group with obesity from fROI analysis, Spearman correlation coefficients were calculated for BOLD signal for HE food (vs. objects) in each individual fROI, and averaged across all six fROIs, and HE food picture appeal rating (vs. objects), as dependent variable, were correlated with BMI. Similarly, in whole brain analysis, within the group with obesity, demeaned BMI was included as a co-variate to identify brain regions where BOLD to signal to HE food (vs. objects) correlated with BMI, using threshold cluster-wise Z>2.3, P<0.05 to control for multiple comparisons.

**Snacks**

Nutritional information relating to the administered snack is given in Supplementary Table S2.

***Ad libitum* test meal**

Initially each dish was tasted in the order of savoury then sweet with randomisation for the order of low fat and high fat between each visit and participant (Supplementary Table S2). Taste was rated for each dish using the Sussex Ingestion Pattern Monitoring rating scales from 0-100mm (SPIM, University of Sussex) (Kissileff et al. 1980) for creaminess, sweetness, 'just right' for both creaminess or sweetness (using the intensity general Labelled Magnitude Scale, gLMS), and liking and pleasantness (using a linear scale), before participants were allowed to eat as much of each dish as they wished. They consumed their test meal alone in a quiet room with minimal distractions, and they were not told that their food intake was measured. The remaining food for each dish was weighed and consumption recorded both as the absolute total energy intake (kcal) and energy intake as a percentage of estimated 24hr resting energy intake (kcal as % REE), calculated using the Cunningham equation (Cunningham 1982), equating lean body mass with fate-free mass determined from bio-electrical impedance analysis at relevant visit.

### SUPPLEMENTARY RESULTS

#### Influence of BMI on food cue reactivity in obesity

In the group with obesity (n=25), there was no significant correlation (Spearman) of BOLD signal during evaluation of HE food pictures (vs. objects) with BMI averaged across all six fROIs (r=0.07, P=0.75), nor in individual fROIs: nucleus accumbens (r=-0.21, P=0.31), caudate (r=0.06, P=0.79), putamen (r=0.92, P=0.93), amygdala (r=0.07, P=0.73), anterior OFC (r=0.36, P=0.08), anterior insula (r=0.11, P=0.62), nor with appeal of HE food (vs. object) pictures (r=0.32, P=0.13). In whole brain analysis, there were no brain regions where BOLD signal to HE food pictures (vs. objects) correlated positively or negatively with BMI (cluster-wise FWE Z>2.3, P<0.05).

**SUPPLEMENTARY TABLES**

**Table S1. Group specific inclusion and exclusion criteria.**

| | **Healthy controls** | **Obesity** | **Ex-smokers** | **Abstinent alcohol dependence** |
|---|---|---|---|---|
| Never drunk alcohol | X | X | X | X |
| >100 cigarettes lifetime use | X | X | ✓ | ✓ |
| Current smoker | X | X | X | ✓ |
| Alcohol use disorder (>4/11 DSM V criteria) | X | X | X | ✓ |
| Previous substance abuse | X | X | X | ✓ |
| Current substance abuse | X | X | X | X |
| Anti-depressant medication | X | X | X | ✓ |

X = excluded from study, ✓ = included in study.

**Table S2. Nutritional information for food provided during study visits.**

| | Snack | | *Ad libitum* lunch | | | |
|---|---|---|---|---|---|---|
| **per 100g** | Plain digestive biscuits | No added sugar jelly | Tomato and Basil soup | Cream of Tomato soup | Vanilla yoghurt | Vanilla ice cream |
| **Category** | n/a | n/a | Low fat, savoury | High fat, savoury | Low fat, sweet | High fat, sweet |
| **Energy (kcal)** | 483.0 | 5.0 | 35.6 | 110.1 | 87.0 | 251.0 |
| **Fat (g)** | 21.30 | 0.00 | 0.98 | 7.87 | 0.00 | 17.00 |
| **of which saturates (g)** | 10.10 | 0.00 | 0.18 | 4.31 | 0.00 | 10.40 |
| **Carbohydrates (g)** | 63.60 | 1.00 | 5.07 | 6.45 | 15.50 | 20.20 |
| **of which sugars (g)** | 15.10 | 1.00 | 4.71 | 5.12 | 15.50 | 14.30 |
| **Fibre (g)** | 3.70 | 0.00 | 0.62 | 0.36 | 0.00 | 0.00 |
| **Protein (g)** | 7.00 | 0.00 | 1.24 | 1.18 | 5.90 | 4.20 |
| **Salt (g)** | 1.30 | 0.15 | 0.38 | 0.63 | 0.23 | 0.15 |
| **Fat (kcal % total)** | 39.7% | 0% | 24.8% | 64.4% | 0% | 61.0% |
| **Carbohydrate (kcal % total)** | 52.7% | 100% | 57.0% | 23.4% | 71.3% | 32.2% |
| **Sugars (kcal % total)** | 12.5% | 100% | 53.0% | 18.6% | 71.3% | 22.8% |
| **Protein (kcal % total)** | 5.8% | 0% | 13.9% | 4.3% | 27.1% | 6.7% |
| **Amount served (g)** | 29.4 (2 biscuits) | 115 | 800 | 800 | 450 | 500 |
| **Amount served (kcal)** | 142.0 | 5.8 | 284.4 | 880.4 | 391.5 | 1255.0 |

Abbreviations: g, grams; kcal, kilocalories; n/a, not applicable

**Table S3. Characteristics of healthy controls without obesity used for generation of functional regions of interest in picture evaluation fMRI task.**

| **N** | 43 | **range** |
|---|---|---|
| **Age (years)** | 25.0 [23.0, 30.0] | 20-51 |
| **Females, n (%)** | 22 (51.2%) | |
| **Caucasian, n (%)** | 23 (56.1%) [a] | |
| **Current smokers, n (%)** | 4 (9.3%) | |
| **Ex-smokers, n (%)** | 4 (9.3%) | |
| **AUDIT, max score 40** | 5.6 ± 3.3 [b] | 0-14 |
| **Alcohol intake (units per week)** | 6.0 [3.0, 10.0] [b] | 0-24 |
| **Body mass index (kg/m$^2$)** | 21.8 [20.3, 24.9] | 17.5-33.5 |
| **% body fat (bio-impedance analysis)** | 22.4 [17.6, 27.1] [a] | 11.3-49.0 |

Data displayed as n (%), mean ± SD or median [interquartile range] with (range).

Data only available for [a] n=41, [b] n=40.

**Table S4. Coordinates of functional regions of interest for picture evaluation fMRI task.**

| Laterality | fROI | Voxels | Z | x | y | z |
|---|---|---|---|---|---|---|
| R | Nucleus accumbens | 42 | 4.62 | 8 | 18 | -4 |
| L | Nucleus accumbens | 44 | 5.86 | -6 | 8 | -4 |
| R | Caudate | 257 | 5.25 | 10 | 2 | 10 |
| L | Caudate | 198 | 5.32 | -6 | 10 | 0 |
| R | Putamen | 422 | 6.02 | 16 | 4 | -12 |
| a | Putamen | 58 | 4.29 | -18 | 20 | -4 |
| L | Putamen | 24 | 4.05 | -14 | 6 | -14 |
| R | Anterior insula | 831 | 8.41 | 38 | 8 | -10 |
| L | Anterior insula | 617 | 8.41 | -38 | 8 | -10 |
| R | Anterior orbitofrontal cortex | 325 | 7.15 | 24 | 38 | -14 |
| L | Anterior orbitofrontal cortex | 348 | 7.16 | -22 | 38 | -14 |
| R | Amygdala | 318 | 5.88 | 18 | 0 | -12 |
| L | Amygdala | 205 | 5.19 | -14 | -4 | -14 |
| R | Hippocampus | 272 | 6.67 | 22 | -34 | 2 |
| L | Hippocampus | 187 | 6.69 | -22 | -28 | -6 |
| R | Dorsolateral prefrontal cortex | 1712 | 7.36 | 46 | 42 | 20 |
| L | Dorsolateral prefrontal cortex | 500 | 5.85 | -34 | 40 | 16 |
| L | Dorsolateral prefrontal cortex | 81 | 4.64 | -22 | 58 | 20 |
| B | Ventral anterior cingulate cortex | 1569 | 8.29 | 0 | 36 | 20 |

Co-ordinates of functional regions of interest (fROI) for picture evaluation functional MRI task determined from group mixed effects analysis of separate cohort of n=43 healthy controls without obesity for contrast of high-energy food or alcohol pictures (vs. neutral). Cluster-wise threshold Z>3.1, FWE P<0.05. x ,y, z co-ordinates given in Montreal Neurological Institute (MNI) space. Abbreviations: R, right; L, left; B, bilateral. See Supplementary Figure S2 for group activation map.

**Table S5. Exploratory between group comparison of high-energy food cue reactivity in functional region of interest analysis for picture evaluation fMRI task.**

| Group contrast | fROI | Mean difference | SEM | 95% Confidence Interval | | Cohen's d | Post-hoc P values | |
|---|---|---|---|---|---|---|---|---|
| | | | | Lower bound | Upper bound | | LSD | Sidak |
| ExS > AAD | NAcc | 0.091 | 0.049 | -0.006 | 0.189 | 0.52 | 0.065 | 0.18 |
| | Caudate | 0.090 | 0.043 | 0.005 | 0.175 | 0.59 | **0.038** | **0.110** |
| | Putamen | 0.113 | 0.038 | 0.036 | 0.189 | 0.82 | **0.004** | **0.013** |
| | Ins | 0.063 | 0.032 | -0.001 | 0.126 | 0.55 | 0.052 | 0.15 |
| | OFC | 0.036 | 0.036 | -0.036 | 0.109 | 0.28 | 0.320 | 0.69 |
| | Amyg | 0.032 | 0.045 | -0.057 | 0.121 | 0.20 | 0.476 | 0.86 |
| | HPC | 0.062 | 0.030 | 0.002 | 0.123 | 0.57 | **0.043** | 0.125 |
| | vACC | 0.063 | 0.040 | -0.016 | 0.142 | 0.44 | 0.118 | 0.31 |
| | dlPFC | 0.049 | 0.031 | -0.012 | 0.109 | 0.45 | 0.116 | 0.31 |
| ExS > OB | NAcc | 0.054 | 0.049 | -0.044 | 0.152 | 0.31 | 0.28 | 0.63 |
| | Caudate | 0.114 | 0.043 | 0.028 | 0.199 | 0.77 | **0.010** | **0.030** |
| | Putamen | 0.146 | 0.030 | 0.069 | 0.223 | 1.08 | **<0.001** | **<0.001** |
| | Ins | 0.071 | 0.032 | 0.006 | 0.135 | 0.62 | **0.031** | **0.092** |
| | OFC | 0.042 | 0.037 | -0.031 | 0.115 | 0.32 | 0.255 | 0.59 |
| | Amyg | 0.055 | 0.045 | -0.035 | 0.145 | 0.35 | 0.225 | 0.53 |
| | HPC | 0.035 | 0.031 | -0.026 | 0.096 | 0.32 | 0.26 | 0.59 |
| | vACC | 0.073 | 0.040 | -0.007 | 0.153 | 0.51 | 0.071 | 0.20 |
| | dlPFC | 0.056 | 0.031 | -0.006 | 0.117 | 0.51 | 0.075 | 0.21 |
| AAD > OB | NAcc | -0.038 | 0.049 | -0.135 | 0.059 | 0.21 | 0.44 | 0.83 |
| | Caudate | 0.024 | 0.043 | -0.061 | 0.109 | 0.16 | 0.58 | 0.92 |
| | Putamen | 0.033 | 0.038 | -0.043 | 0.110 | 0.25 | 0.39 | 0.77 |
| | Ins | 0.008 | 0.032 | -0.056 | 0.071 | 0.07 | 0.81 | 0.99 |
| | OFC | 0.006 | 0.036 | -0.067 | 0.078 | 0.05 | 0.88 | 0.99 |
| | Amyg | 0.023 | 0.045 | -0.066 | 0.112 | 0.14 | 0.61 | 0.93 |
| | HPC | -0.027 | 0.030 | -0.088 | 0.033 | 0.25 | 0.37 | 0.75 |
| | vACC | 0.011 | 0.040 | -0.068 | 0.089 | 0.08 | 0.79 | 0.99 |
| | dlPFC | 0.007 | 0.031 | -0.054 | 0.068 | 0.06 | 0.82 | 0.99 |

Post-hoc between group comparisons for BOLD signal for high-energy food vs. neutral picture contrast in picture evaluation fMRI task between adults with obesity (OB, n=25), ex-smokers (ExS, n-25) and abstinent alcohol dependence (AAD, n=26) in functional regions of interest (fROI) using repeated measures ANOVA with post-hoc Fisher LSD (uncorrected) or Sidak (corrected for multiple comparisons) tests. Abbreviations: Amyg, amygdala; dlPFC, dorsolateral prefrontal cortex; HPC, hippocampus; Ins, anterior insula; LSD, least significant difference; NAcc, nucleus accumbens; OFC, anterior orbitofrontal cortex; SEM, standard error of the mean; vACC, ventral anterior cingulate cortex.

**Table S6. Spatial coordinates from whole brain analysis for high-energy food cue reactivity during picture evaluation fMRI task within individual groups.**

| Group | Cluster | Voxels | Z max | x | y | z | Laterality | Location |
|---|---|---|---|---|---|---|---|---|
| AAD | 1 | 11,353 | 8.16 | 6 | -80 | -2 | R | Lingual gyrus (48%), Intracalcarine cortex (24%) |
| | | | 8.06 | 14 | -88 | -6 | R | Lingual gyrus (26%), Occipital pole (16%), Occipital fusiform gyrus (14%) |
| | | | 8.03 | -8 | -86 | -8 | L | Lingual gyrus (47%), Occipital fusiform gyrus (7%) |
| | | | 8.00 | -4 | -82 | -6 | L | Lingual gyrus (76%), Intracalcarine cortex (8%) |
| | | | 6.75 | 26 | -74 | -12 | R | Occipital fusiform gyrus (64%) |
| | | | 6.54 | 22 | -80 | -12 | R | Occipital fusiform gyrus (49%), Lingual gyrus (8%) |
| | 2 | 2,515 | 4.73 | -38 | 6 | -10 | L | Insula (95%) |
| | | | 4.64 | -20 | 4 | -18 | L | Anterior parahippocampal gyrus (6%) |
| | | | 4.28 | 20 | -24 | 10 | R | Thalamus (77%), Cerebral white matter (23%) |
| | | | 3.92 | -38 | -4 | 6 | L | Insula (84%) |
| | | | 3.66 | 12 | -18 | 10 | R | Thalamus (100%) |
| | | | 3.63 | -2 | -14 | 2 | L | Thalamus (100%) |
| | 3 | 1,115 | 4.88 | 24 | -70 | 52 | R | Lateral occipital cortex superior division (60%) |
| | | | 3.32 | 32 | -54 | 66 | R | Superior parietal lobule (48%), Lateral occipital cortex superior division (14%) |
| | | | 3.3 | 28 | -54 | 68 | R | Superior parietal lobule (43%), Lateral occipital cortex superior division (23%) |
| | | | 3.02 | 34 | -52 | 54 | R | Superior parietal lobule (51%), Angular gyrus (13%),<br>Lateral occipital cortex superior division (5%) |
| | | | 2.91 | 30 | -60 | 64 | R | Lateral occipital cortex superior division (55%), Superior parietal lobule (13%) |
| | | | 2.91 | 42 | -48 | 64 | R | Superior parietal lobule (41%) |
| | 4 | 952 | 4.38 | 4 | -32 | 34 | R | Posterior cingulate gyrus (91%) |
| | | | 4.14 | -4 | -32 | 32 | L | Posterior cingulate gyrus (63%) |
| | | | 3.52 | -2 | -6 | 28 | L | Anterior cingulate gyrus (36%) |
| | | | 3.19 | 0 | 12 | 22 | B | Anterior cingulate gyrus (21%) |

| | | | | | | | | |
|---|---|---|---|---|---|---|---|---|
| | | | 3.1 | -2 | 18 | 32 | L | Anterior cingulate gyrus (76%) |
| | | | 3.09 | -2 | 28 | 26 | L | Anterior cingulate gyrus (74%), Paracingulate gyrus (20%) |
| | 5 | 662 | 4.58 | -22 | -64 | 44 | L | Lateral occipital cortex superior division (45%) |
| | | | 4.55 | -24 | -66 | 52 | L | Lateral occipital cortex superior division (62%) |
| | | | 3.17 | -24 | -68 | 62 | L | Lateral occipital cortex superior division (52%) |
| | | | 2.58 | -20 | -60 | 66 | L | Lateral occipital cortex superior division (49%), Superior parietal lobule (29%) |
| ExS | 1 | 56,990 | 9.03 | 4 | -84 | -6 | R | Lingual gyrus (54%), Intracalcarine cortex (14%) |
| | | | 8.73 | 6 | -76 | 2 | R | Lingual gyrus (44%), Intracalcarine cortex (35%) |
| | | | 8.71 | -10 | -84 | -12 | L | Lingual gyrus (40%), Occipital fusiform gyrus (25%) |
| | | | 8.64 | -4 | -80 | -6 | L | Lingual gyrus (71%), Intracalcarine cortex (12%) |
| | | | 8.25 | -10 | -88 | -4 | L | Lingual gyrus (25%), Intracalcarine cortex (17%), Occipital pole (8%) |
| | | | 8.21 | 24 | -80 | -14 | R | Occipital fusiform gyrus (59%), Lingual gyrus (8%) |
| | 2 | 982 | 5.11 | -38 | 40 | 14 | L | Frontal pole (49%), Middle frontal gyrus (8%), Inferior frontal gyrus pars triangularis (7%) |
| | | | 3.41 | -30 | 32 | 16 | L | Cerebral white matter (90%), Cerebral cortex (10%) |
| | | | 3.08 | -46 | 40 | 28 | L | Frontal pole (19%), Middle frontal gyrus (14%) |
| OB | 1 | 21,112 | 9.33 | 6 | -82 | -4 | R | Lingual gyrus (54%), Intracalcarine cortex (19%) |
| | | | 9.27 | 12 | -88 | -6 | R | Lingual gyrus (24%), Occipital pole (16%), Occipital fusiform gyrus (13%) |
| | | | 9.09 | -4 | -80 | -6 | L | Lingual gyrus (80%) |
| | | | 9.03 | -10 | -90 | -2 | L | Occipital pole (23%), Intracalcarine cortex (20%), Lingual gyrus (17%) |
| | | | 9.01 | -12 | -94 | 0 | L | Occipital pole (52%), Intracalcarine cortex (6%) |
| | | | 8.27 | -8 | -84 | -12 | L | Lingual gyrus (39%), Occipital fusiform gyrus (21%) |

Spatial coordinates of significant clusters from whole brain mixed effects analysis of high-energy food picture (vs. neutral) contrast in picture evaluation fMRI task for individual groups: abstinent alcohol dependence (AAD, n=26), ex-smokers (ExS, n=25) or adults with obesity (OB, n=25). Cluster-wise threshold Z>2.3, family-wise error (FWE) P<0.05. x ,y, z co-ordinates given in Montreal Neurological Institute (MNI) space. Location and percentage likelihood given using the Harvard-Oxford cortical and subcortical structural atlases. Abbreviations: B, bilateral; R, right; L, left. See Supplementary Figures S3, S4 and S5 for group activation maps.

**Table S7. Whole brain analysis of between group differences in high-energy food cue reactivity in picture evaluation fMRI task.**

| Contrast | Cluster | Voxels | Z max | x | y | z | Laterality | Location |
|---|---|---|---|---|---|---|---|---|
| High-energy food > neutral picture | 1 | 940 | 4.48 | 28 | 18 | -6 | R | Anterior insula (21%), Orbitofrontal cortex (7%) |
| | | | 3.56 | 20 | 16 | 4 | R | Putamen (9%), Caudate (7%) |
| | | | 3.48 | 30 | -10 | 12 | R | Putamen (29%) |
| | | | 3.26 | 32 | -16 | 6 | R | Putamen (32%) |
| | | | 3.12 | 12 | 0 | 20 | R | Caudate (67%) |
| | | | 3.07 | 10 | 18 | 0 | R | Caudate (81%), Nucleus accumbens (11%) |

Spatial coordinates of significant clusters from whole brain mixed effects analysis for high-energy food pictures (vs. neutral) contrast in picture evaluation fMRI task showing between group differences for obesity (OB, n=25), ex-smokers (ExS, n=25) and abstinent alcohol dependence (AAD, n=26) groups using ANOVA. Cluster-wise threshold Z>2.3, family-wise error (FWE) P<0.05. x ,y, z co-ordinates given in Montreal Neurological Institute (MNI) space. Location and percentage likelihood given using the Harvard-Oxford cortical and subcortical structural atlases. Abbreviations: R, right. See main paper Figure 2 for group difference activation map.

**Table S8. Greater high-energy food cue reactivity across putamen, caudate and insula from whole brain analysis in ex-smokers in picture evaluation fMRI task.**

| Group contrast | Mean difference | SEM | 95% Confidence Interval | | Cohen's d | Post-hoc P values | |
|---|---|---|---|---|---|---|---|
| | | | Lower bound | Upper bound | | LSD | Sidak |
| ExS > OB | 0.128 | 0.030 | 0.067 | 0.188 | 1.22 | **<0.001** | **<0.001** |
| ExS > AAD | 0.100 | 0.030 | 0.041 | 0.160 | 0.94 | **0.001** | **0.004** |
| AAD > OB | 0.027 | 0.030 | -0.032 | 0.087 | 0.26 | 0.36 | 0.74 |

Post-hoc comparisons of extracted median BOLD signal for high-energy food vs. neutral picture contrast in picture evaluation fMRI task adults between groups with obesity (OB, n=25), ex-smokers (ExS, n-25) and abstinent alcohol dependence (AAD, n=26), averaged across significant cluster from between group whole brain analysis (see main paper Figure 2 for group difference activation map, Supplementary Table S7 for cluster co-ordinates), using repeated measures ANOVA with post-hoc Fisher LSD (uncorrected) or Sidak (corrected for multiple comparisons) tests. Abbreviations: LSD, least significant difference; SEM, standard error of the mean.

**Table S9. Greater high-energy food cue reactivity in putamen, caudate and insula from exploratory post-hoc whole brain analysis in ex-smokers in picture evaluation fMRI task.**

| Group contrast | fROI | Mean difference | SEM | 95% Confidence Interval | | Cohen's d | Post-hoc P values | |
|---|---|---|---|---|---|---|---|---|
| | | | | Lower bound | Upper bound | | LSD | Sidak |
| ExS > AAD | Putamen | 0.112 | 0.036 | 0.040 | 0.183 | 0.87 | **0.003** | **0.008** |
| | Caudate | 0.111 | 0.037 | 0.037 | 0.186 | 0.83 | **0.004** | **0.012** |
| | Insula | 0.078 | 0.026 | 0.027 | 0.129 | 0.86 | **0.003** | **0.009** |
| ExS > OB | Putamen | 0.164 | 0.036 | 0.092 | 0.235 | 1.26 | **<0.001** | **<0.001** |
| | Caudate | 0.125 | 0.038 | 0.049 | 0.200 | 0.92 | **0.001** | **0.004** |
| | Insula | 0.095 | 0.026 | 0.044 | 0.147 | 1.06 | **<0.001** | **0.001** |
| AAD > OB | Putamen | 0.052 | 0.036 | -0.019 | 0.123 | 0.41 | 0.15 | 0.39 |
| | Caudate | 0.014 | 0.037 | -0.061 | 0.088 | 0.10 | 0.72 | 0.98 |
| | Insula | 0.017 | 0.026 | -0.034 | 0.068 | 0.19 | 0.51 | 0.88 |

Exploratory post-hoc comparisons of extracted median BOLD signal for high-energy food vs. neutral picture contrast in picture evaluation fMRI task adults between groups with obesity (OB, n=25), ex-smokers (ExS, n-25) and abstinent alcohol dependence who are abstinent (AAD, n=26), within individual functional regions of interest (fROI) from the significant cluster determined from between group whole brain analysis (see main paper Figure 2 for group difference activation map, Supplementary Table S7 for cluster co-ordinates), using repeated measures ANOVA with post-hoc Fisher LSD (uncorrected) or Sidak (corrected for multiple comparisons) tests. Abbreviations: LSD, least significant difference; SEM, standard error of the mean.

**Table S10. Spatial coordinates from whole brain analysis for high-energy food cue reactivity during picture evaluation fMRI task between individual groups.**

| Group Contrast | Cluster | Voxels | Z max | x | y | z | Laterality | Location |
|---|---|---|---|---|---|---|---|---|
| ExS > AAD | 1 | 1857 | 4.78 | 28 | 18 | -6 | R | Insula (21%), Orbitofrontal cortex (7%) |
| | | | 3.73 | 8 | -12 | 28 | R | Posterior cingulate gyrus (5%) |
| | | | 3.66 | 12 | -2 | 20 | R | Caudate (63%) |
| | | | 3.60 | 32 | 26 | 18 | R | Cerebral white matter (94%) |
| | | | 3.58 | 10 | 18 | 0 | R | Caudate (81%), Nucleus accumbens (11%) |
| | | | 3.55 | 32 | 34 | 6 | R | Inferior frontal gyrus (6%) |
| | 2 | 1369 | 4.09 | 16 | 36 | 58 | R | Superior frontal gyrus (6%), Frontal pole (8%) |
| | | | 3.50 | 8 | 58 | 40 | R | Frontal pole (45%) |
| | | | 3.35 | 10 | 28 | 42 | R | Paracingulate gyrus (18%), Superior frontal gyrus (16%) |
| | | | 3.23 | 8 | 20 | 56 | R | Superior frontal gyrus (36%) |
| | | | 3.14 | -18 | 14 | 40 | L | White matter (98%) |
| | | | 3.10 | 14 | 8 | 56 | R | Anterior dorsal pre-motor area (31%), Supplementary motor area (37%) [a] |
| | 3 | 991 | 4.19 | -22 | -2 | -8 | L | Pallidum (12%), Putamen (8%) |
| | | | 3.86 | -26 | -28 | 6 | L | Cerebral white matter (98%) |
| | | | 3.59 | -28 | 6 | 2 | L | Putamen (97%) |
| | | | 3.50 | -26 | -34 | 0 | L | Hippocampus (7%) |
| | | | 3.25 | -32 | -4 | 2 | L | Putamen (43%) |
| | | | 3.21 | -30 | -4 | 14 | L | White matter (97%) |

| | | | | | | | | |
|---|---|---|---|---|---|---|---|---|
| | 4 | 731 | 3.50 | -38 | 38 | -10 | L | Frontal pole (33%), Orbitofrontal cortex (11%) |
| | | | 3.22 | -28 | 46 | -2 | L | Frontal pole (6%) |
| | | | 3.18 | -28 | 52 | -4 | L | Frontal pole (24%) |
| | | | 3.15 | -32 | 46 | 12 | L | Frontal pole (56%) |
| | | | 3.05 | -30 | 52 | 6 | L | Frontal pole (72%) |
| | | | 2.85 | -28 | 54 | 12 | L | Frontal pole (82%) |
| AAD > ExS | Nil | | | | | | | |
| ExS > OB | 1 | 6135 | 4.10 | 22 | 16 | 4 | R | Putamen (43%) |
| | | | 4.05 | 30 | -10 | 12 | R | Putamen (29%) |
| | | | 4.03 | -26 | -26 | 8 | L | Putamen (7%) |
| | | | 4.03 | -24 | 2 | -6 | L | Putamen (93%) |
| | | | 3.97 | 28 | 16 | -6 | R | Insula (9%), Putamen (6%) |
| | | | 3.87 | -26 | 4 | -2 | L | Putamen (100%) |
| OB > ExS | Nil | | | | | | | |
| AAD > OB | Nil | | | | | | | |
| OB > AAD | Nil | | | | | | | |

Spatial coordinates of significant clusters from whole brain mixed effects analysis of high-energy food picture (vs. neutral) contrast in picture evaluation fMRI task betweenl groups as unpaired t-test: abstinent alcohol dependence (AAD, n=26), ex-smokers (ExS, n=25) or adults with obesity (OB, n=25). Cluster-wise threshold Z>2.3, family-wise error (FWE) P<0.05. x ,y, z co-ordinates given in Montreal Neurological Institute (MNI) space. Location and percentage likelihood given using the Harvard-Oxford cortical and subcortical structural atlases, except [a] Sallet dorsal frontal connectivity-based parcellation atlas. Abbreviations: B, bilateral; R, right; L, left. See Supplementary Figure S6 for group activation maps.

**Table S11. Spatial coordinates from whole brain analysis for alcohol cue reactivity during picture evaluation fMRI task within individual groups.**

| Group | Cluster | Voxels | Z max | x | y | z | Laterality | Location |
|---|---|---|---|---|---|---|---|---|
| AAD | 1 | 3454 | 5.24 | 8 | -66 | 42 | R | Precuneus (49%) |
| | | | 4.69 | 8 | -64 | 32 | R | Precuneus (46%) |
| | | | 4.64 | -4 | -22 | 30 | L | Posterior cingulate gyrus (46%), Anterior cingulate gyrus (6%) |
| | | | 4.54 | 6 | -22 | 30 | R | Posterior cingulate gyrus (39%), Anterior cingulate gyrus (5%) |
| | | | 4.35 | 8 | -52 | 30 | R | Posterior cingulate gyrus (38%), Precuneus cortex (36%) |
| | | | 4.09 | 6 | -44 | 24 | R | Posterior cingulate gyrus (46%) |
| | 2 | 2265 | 5.35 | 6 | -80 | 0 | R | Intracalcarine cortex (38%), Lingual gyrus (38%) |
| | | | 4.74 | -6 | -96 | 6 | L | Occipital pole (63%) |
| | | | 4.61 | 12 | -96 | 18 | R | Occipital pole (68%) |
| | | | 4.38 | 16 | -98 | 12 | R | Occipital pole (64%) |
| | | | 4.34 | -4 | -80 | -2 | L | Lingual gyrus (68%), Intracalcarine cortex (15%) |
| | | | 4.34 | -4 | -86 | -6 | L | Intracalcarine cortex (62%), Lingual gyrus (17%) |
| ExS | 1 | 4384 | 5.45 | 4 | -42 | 30 | R | Posterior cingulate gyrus (72%) |
| | | | 5.29 | -8 | -66 | 40 | L | Precuneus cortex (57%) |
| | | | 5.17 | 6 | -44 | 24 | R | Posterior cingulate gyrus (46%) |
| | | | 5.16 | 2 | -62 | 42 | R | Precuneus cortex (88%) |
| | | | 5.02 | 6 | -54 | 32 | R | Precuneus cortex (52%), Posterior cingulate gyrus (34%) |
| | | | 5.00 | 4 | -58 | 38 | R | Precuneus cortex (89%) |
| | 2 | 3312 | 4.24 | 24 | 42 | 44 | R | Frontal pole (68%), Superior frontal gyrus (10%) |
| | | | 4.23 | 22 | 38 | 40 | R | Frontal pole (47%), Superior frontal gyrus (15%), Middle frontal gyrus (8%) |
| | | | 4.14 | 38 | 22 | 50 | R | Middle frontal gyrus (64%) |

| | | | | | | | | |
|---|---|---|---|---|---|---|---|---|
| | | | 4.01 | 44 | 20 | 58 | R | nil |
| | | | 3.98 | 40 | 30 | 54 | R | nil |
| | | | 3.87 | 8 | 44 | 12 | R | Paracingulate gyrus (46%), Anterior cingulate gyrus (40%) |
| | 3 | 2292 | 5.28 | -4 | -78 | 0 | L | Lingual gyrus (65%), Intracalcarine cortex (18%) |
| | | | 4.56 | -4 | -96 | 6 | L | Occipital pole (72%) |
| | | | 4.40 | -6 | -96 | 14 | L | Occipital pole (59%) |
| | | | 4.29 | 8 | -96 | -6 | R | Occipital pole (50%) |
| | | | 4.13 | 2 | -90 | 8 | R | Occipital pole (36%), Supracalcarine cortex (25%), Cuneal cortex (9%), Intracalcarine cortex (8%), Lingual gyrus (5%) |
| | | | 3.93 | -6 | -94 | 18 | L | Occipital pole (59%), Cuneal cortex (5%) |
| | 4 | 1762 | 4.6 | 50 | -66 | 48 | R | Lateral occipital cortex superior division (24%) |
| | | | 4.43 | 40 | -64 | 38 | R | Lateral occipital cortex superior division (45%) |
| | | | 4.27 | 50 | -52 | 56 | R | Angular gyrus (34%), Lateral occipital cortex superior division (5%), Posterior supramarginal gyrus (5%) |
| | | | 4.12 | 54 | -56 | 42 | R | Angular gyrus (57%), Lateral occipital cortex superior division (26%) |
| | | | 3.90 | 56 | -54 | 48 | R | Angular gyrus (44%), Lateral occipital cortex superior division (10%) |
| | | | 3.85 | 54 | -46 | 54 | R | Posterior supramarginal gyrus (31%), Angular gyrus (30%) |
| | 5 | 1281 | 4.61 | 28 | 58 | 2 | R | Frontal pole (78%) |
| | | | 4.32 | 38 | 54 | -4 | R | Frontal pole (72%) |
| | | | 3.58 | 20 | 52 | -4 | R | Frontal pole (16%) |
| | | | 3.47 | 22 | 50 | -10 | R | Frontal pole (30%) |
| | | | 3.43 | 26 | 66 | 10 | R | Frontal pole (72%) |
| | | | 3.40 | 12 | 60 | 0 | R | Frontal pole (13%) |

| | | | | | | | | |
|---|---|---|---|---|---|---|---|---|
| OB | 1 | 3454 | 5.46 | -4 | -78 | -2 | L | Lingual gyrus (75%), Intracalcarine cortex (7%) |
| | | | 5.24 | 4 | -80 | -2 | R | Lingual gyrus (56%), Intracalcarine cortex (19%) |
| | | | 5.05 | -8 | -96 | 10 | L | Occipital pole (61%) |
| | | | 5.04 | 8 | -96 | 12 | R | Occipital pole (71%) |
| | | | 4.40 | -12 | -94 | 0 | L | Occipital pole (52%), Intracalcarine cortex (6%) |
| | | | 3.85 | 14 | -92 | 0 | R | Occipital pole (52%), Intracalcarine cortex (7%), Lingual gyrus (5%) |
| | 2 | 2704 | 4.68 | 6 | -70 | 44 | R | Precuneus cortex (63%) |
| | | | 4.68 | 10 | -68 | 40 | R | Precuneus cortex (48%), Cuneal cortex (5%) |
| | | | 4.21 | 2 | -24 | 36 | R | Posterior cingulate gyrus (95%), Anterior cingulate gyrus (5%) |
| | | | 4.16 | 4 | -42 | 26 | R | Posterior cingulate gyrus (69%), Precuneus cortex (27%) |
| | | | 3.89 | -2 | -34 | 30 | L | Posterior cingulate gyrus (98%) |
| | | | 3.87 | 2 | -32 | 30 | R | Posterior cingulate gyrus (98%) |
| | 3 | 1252 | 4.26 | 38 | -64 | 52 | R | Lateral occipital cortex superior division (67%) |
| | | | 4.03 | 36 | -62 | 46 | R | Lateral occipital cortex superior division (59%), Angular gyrus (7%) |
| | | | 3.56 | 42 | -48 | 38 | R | Angular gyrus (30%), Posterior supramarginal gyrus (12%) |
| | | | 3.47 | 42 | -48 | 48 | R | Angular gyrus (29%), Superior parietal lobule (19%), Posterior supramarginal gyrus (18%) |
| | | | 3.36 | 50 | -42 | 52 | R | Posterior supramarginal gyrus (57%), Angular gyrus (13%) |
| | | | 3.20 | 40 | -44 | 32 | R | Posterior supramarginal gyrus (6%) |

Spatial coordinates of significant clusters from whole brain mixed effects analysis of preferred alcohol picture (vs. neutral) contrast in picture evaluation fMRI task within individual groups: abstinent alcohol dependence (AAD, n=26), ex-smokers (ExS, n=25) or adults with obesity (OB, n=25). Cluster-wise threshold Z>2.3, family-wise error (FWE) P<0.05. x ,y, z co-ordinates given in Montreal Neurological Institute (MNI) space. Location and percentage likelihood given using the Harvard-Oxford cortical and subcortical structural atlases. Abbreviations: B, bilateral; R, right; L, left. See Supplementary Figures S8, S9 and S10 for group activation maps.

**Table S12. Spatial coordinates from whole brain analysis for cigarette cue reactivity during picture evaluation fMRI task within individual groups.**

| Group | Cluster | Voxels | Z max | x | y | z | Laterality | Location (%) |
|---|---|---|---|---|---|---|---|---|
| ExS | 1 | 4370 | 4.22 | -4 | 16 | 6 | L | Lateral ventricle (94%), White matter (5%) |
| | | | 4.07 | -14 | 46 | 0 | L | Paracingulate gyrus (21%), Anterior cingulate gyrus (5%), Medial frontal cortex (5%) |
| | | | 4.02 | 22 | 50 | 0 | R | Frontal pole (7%) |
| | | | 3.98 | 8 | 44 | 10 | R | Anterior cingulate gyrus (49%), Paracingulate gyrus (41%) |
| | | | 3.96 | -20 | 54 | -8 | L | Frontal pole (23%) |
| | | | 3.92 | 2 | 8 | 6 | R | Lateral ventricle (83%) |
| | 2 | 3394 | 4.70 | 4 | -16 | 30 | R | Anterior cingulate gyrus (48%), Posterior cingulate gyrus (24%) |
| | | | 4.60 | 4 | -24 | 30 | R | Posterior cingulate gyrus (85%) |
| | | | 4.28 | -4 | -16 | 30 | L | Anterior cingulate gyrus (28%), Posterior cingulate gyrus (14%) |
| | | | 4.15 | 2 | -42 | 26 | R | Posterior cingulate gyrus (97%) |
| | | | 4.13 | -6 | -32 | 28 | L | Posterior cingulate gyrus (37%) |
| | | | 3.95 | 8 | -70 | 40 | R | Precuneus (55%), Cuneal cortex (10%) |
| | 3 | 2017 | 4.77 | 56 | -54 | 38 | R | Angular gyrus (73%), Lateral occipital cortex superior division (7%) |
| | | | 4.48 | 58 | -48 | 38 | R | Angular gyrus (47%), Posterior supramarginal gyrus (30%) |
| | | | 4.44 | 52 | -48 | 34 | R | Angular gyrus (55%), Posterior supramarginal gyrus (15%) |
| | | | 4.28 | 50 | -68 | 48 | R | Lateral occipital cortex superior division (7%) |
| | | | 4.07 | 44 | -72 | 46 | R | Lateral occipital cortex superior division (48%) |
| | | | 3.99 | 54 | -42 | 44 | R | Posterior supramarginal gyrus (57%), Angular gyrus (16%) |
| | 4 | 864 | 4.96 | -52 | -48 | 44 | L | Posterior supramarginal gyrus (51%), Angular gyrus (19%) |
| | | | 4.69 | -54 | -50 | 48 | L | Posterior supramarginal gyrus (51%), Angular gyrus (28%) |

| | | | | | | | | |
|---|---|---|---|---|---|---|---|---|
| | | | 3.31 | -46 | -44 | 38 | L | Posterior supramarginal gyrus (19%), Anterior supramarginal gyrus (7%) |
| | | | 3.30 | -60 | -42 | 44 | L | Posterior supramarginal gyrus (44%), Anterior supramarginal gyrus (32%) |
| | | | 2.98 | -54 | -52 | 26 | L | Angular gyrus (29%), Posterior supramarginal gyrus (26%), |
| | | | 2.92 | -38 | -66 | 50 | L | Lateral occipital cortex superior division (68%) |
| OB | 1 | 5122 | 5.51 | 10 | -54 | 42 | R | Precuneus (80%) |
| | | | 5.23 | 8 | -56 | 36 | R | Precuneus (62%), Posterior cingulate gyrus (9%) |
| | | | 5.22 | 0 | -42 | 26 | B/L | Posterior cingulate gyrus (91%) |
| | | | 5.21 | 4 | -42 | 24 | R | Posterior cingulate gyrus (81%) |
| | | | 5.01 | 2 | -22 | 36 | R | Posterior cingulate gyrus (91%), Anterior cingulate gyrus (9%) |
| | | | 4.84 | 16 | -50 | 36 | R | Precuneus (14%), Posterior cingulate gyrus (7%) |
| | 2 | 4073 | 4.86 | 20 | 42 | 26 | R | Frontal pole (21%) |
| | | | 4.85 | -12 | 46 | 2 | L | Paracingulate gyrus (57%), Anterior cingulate gyrus (13%) |
| | | | 4.41 | -32 | 30 | 44 | L | Middle frontal gyrus (74%) |
| | | | 4.37 | 8 | 42 | 6 | R | Anterior cingulate gyrus (52%), Paracingulate gyrus (27%) |
| | | | 4.33 | 6 | 44 | 10 | R | Anterior cingulate gyrus (62%), Paracingulate gyrus (34%) |
| | | | 4.19 | -18 | 58 | 6 | L | Frontal pole (18%) |
| | 3 | 2705 | 5.86 | -52 | -48 | 44 | L | Supramarginal gyrus posterior division (51%), Angular gyrus (19%) |
| | | | 5.09 | -62 | -34 | 36 | L | Anterior supramarginal gyrus (80%), Posterior supramarginal gyrus (5%) |
| | | | 3.88 | -36 | -50 | 28 | L | White matter (95%) |
| | | | 3.68 | -36 | -66 | 14 | L | Lateral occipital cortex inferior division (18%), Lateral occipital cortex superior division (11%), Middle temporal gyrus (5%) |
| | | | 3.6 | -18 | -44 | 18 | L | White matter (62%), Lateral ventricle (38%) |
| | | | 3.44 | -36 | -68 | 18 | L | Lateral occipital cortex superior division (34%), Lateral occipital cortex inferior division |

| | | | | | | | | |
|---|---|---|---|---|---|---|---|---|
| | | | | | | | | (7%) |
| | 4 | 2411 | 5.14 | 48 | -48 | 40 | R | Angular gyrus (32%), Posterior supramarginal gyrus (17%) |
| | | | 4.71 | 60 | -46 | 38 | R | Posterior supramarginal gyrus (53%), Angular gyrus (29%) |
| | | | 4.35 | 52 | -60 | 42 | R | Lateral occipital cortex superior division (60%), Angular gyrus (25%) |
| | | | 4.26 | 64 | -36 | 42 | R | Posterior supramarginal gyrus (44%), Anterior supramarginal gyrus (12%) |
| | | | 3.82 | 42 | -42 | 30 | R | White matter (96%) |
| | | | 3.68 | 48 | -62 | 52 | R | Lateral occipital cortex superior division (36%), Angular gyrus (8%) |

Spatial coordinates of significant clusters from whole brain mixed effects analysis of cigarette picture (vs. neutral) contrast in picture evaluation fMRI task within individual groups: ex-smokers (ExS, n=25) or adults with obesity (OB, n=25). Cluster-wise threshold Z>2.3, family-wise error (FWE) P<0.05. x ,y, z co-ordinates given in Montreal Neurological Institute (MNI) space. Location and percentage likelihood given using the Harvard-Oxford cortical and subcortical structural atlases. Abbreviations: B, bilateral; R, right; L, left. See Supplementary Figures S12 and S13 for group activation maps.

**Table S13. Spatial coordinates from whole brain analysis for cigarette cue reactivity during picture evaluation fMRI task between ex-smokers and obesity.**

| Group Contrast | Cluster | Voxels | Z max | x | y | z | Laterality | Location |
|---|---|---|---|---|---|---|---|---|
| ExS > OB | 1 | 1587 | 3.66 | 52 | 30 | 26 | R | Middle frontal gyrus (54%), Inferior frontal gyrus pars opercularis (9%), Cluster 6 area 9/46V (29%) [a] |
| | | | 3.54 | 44 | 20 | 0 | R | Frontal operculum cortex (59%), Frontal orbital cortex (12%) |
| | | | 3.53 | 40 | 26 | -6 | R | Frontal orbital cortex (63%), Frontal operculum cortex (11%) |
| | | | 3.51 | 36 | 20 | 2 | R | Insula (72%), Frontal operculum cortex (10%) |
| | | | 3.41 | 50 | 40 | 24 | R | Frontal pole (41%), Middle frontal gyrus (14%) |
| | | | 3.39 | 50 | 38 | 30 | R | Frontal pole (13%) |
| | 2 | 901 | 3.92 | -22 | 40 | -14 | L | Frontal operculum cortex (52%), Frontal orbital cortex (7%) |
| | | | 3.82 | -30 | 18 | -4 | L | Insula (45%) |
| | | | 3.43 | -30 | 16 | 0 | L | Insula (8%) |
| | | | 3.25 | -32 | 28 | 4 | L | Frontal orbital cortex (19%), Insula (19%), Inferior frontal gyrus pars triangularis (17%), Frontal operculum cortex (135) |
| | | | 3.23 | -40 | 42 | 4 | L | Frontal pole (56%) |
| | | | 3.2 | -38 | 16 | -4 | L | Insula (89%) |
| OB > ExS | Nil | | | | | | | |

Spatial coordinates of significant clusters from whole brain mixed effects analysis of cigarette picture (vs. neutral) contrast in picture evaluation fMRI task between groups as unpaired t-test: ex-smokers (ExS, n=25) or adults with obesity (OB, n=25). Cluster-wise threshold Z>2.3, family-wise error (FWE) P<0.05. x ,y, z co-ordinates given in Montreal Neurological Institute (MNI) space. Location and percentage likelihood given using the Harvard-Oxford cortical and subcortical structural atlases, except [a] Sallet dorsal frontal connectivity-based parcellation atlas. Abbreviations: B, bilateral; R, right; L, left. See main Figure 4 for group activation map.

**Table S14. Eating behaviour questionnaires.**

| | Healthy controls (HC) | Obesity (OB) | Ex-smokers (ExS) | Abstinent alcohol dependence (AAD) | Group comparisons | |
|---|---|---|---|---|---|---|
| | | | | | Test statistic | P-value |
| **N** | 23 | 26 | 25 | 25 | | |
| **DEBQ restraint (score 0-5)** | 2.30<br>[1.60, 2.60] | 3.10<br>[2.78, 3.50] ** | 2.40<br>[1.60, 3.40] | 2.70<br>[1.68, 3.30] | H(3)=12.1 | **0.007** |
| **TFEQ restraint (score 0-21)** | 7.70 ± 3.89<br>(0-14) | 8.73 ±5.02<br>(0-18) | 7.88 ± 4.60<br>(1-17) | 9.11 ± 6.05<br>(0-20) | F(3,95)=2.02 | 0.11 |
| **PFS (score 21-105)** | 43.0<br>[35.0, 50.0] | 61.0<br>[41.0, 75.5] * | 42.0<br>[35.0, 55.5] | 50.5<br>[34.8, 61.5] | (H3)=8.17 | **0.043** |
| **DEBQ emotional (score 0-5)** | 1.62<br>[1.31, 2.54] | 2.89<br>[2.19, 4.02] * | 1.69<br>[1.19, 2.38] * | 2.39<br>[1.77, 2.87] | H(3)=12.3 | **0.006** |
| **TFEQ disinhibition (score 0-16)** | 3.0<br>[2.0, 7.0] | 10.0<br>[6.5, 13.5] *** | 5.0<br>[3.0, 7.0] ## | 5.0<br>[2.0, 9.0] ## | H(3)=24.2 | **<0.001** |
| **YFAS (score 0-7)** | 1<br>[1, 1] | 2.5<br>[1.0, 4.3] ** | 0<br>[0, 0] * ### | 1<br>[1, 2.3] ††† | H(3)=52.8 | **<0.001** |
| **DEBQ external (score 0-5)** | 2.73 ± 0.65<br>(1.80-4.20) | 3.18 ± 0.84<br>(1.70-5.00) | 2.95 ± 0.65<br>(1.60-4.20) | 3.05 ± 0.45<br>(2.10-3.80) | F(3,96)=1.93 | 0.13 |
| **TFEQ hunger (score 0-14)** | 4.0<br>[2.0, 7.0] | 7.0<br>[3.5, 10.0] | 5.0<br>[3.0, 6.5] | 5.5<br>[2.0, 8.0] | H(3)=4.89 | 0.18 |
| **BES (score 0-46)** | 5.0<br>[4.0, 10.0] | 21.0<br>[8.0, 26.0] *** | 5.5<br>[3.0, 12.0] ## | 9.0<br>[3.0, 16.25] | H(3)=19.9 | **<0.001** |

Data displayed as mean ± SD (range) or median [interquartile range]. Analysed as one-way ANOVA with post-hoc Sidak test for parametric data, Kruskal-Wallis test with post-hoc Bonferroni correction, or Mann-Whitney U test for non-parametric data. Post hoc tests: vs. HC *P<0.05, **P<0.01, ***P<0.001; vs. OB #P<0.05, ##P<0.01, ###P<0.001; vs. ExS †P<0.05, ††P<0.01, †††P<0.001. Abbreviations: BES, Binge Eating Scale; DEBQ, Dutch Eating Behaviour Questionnaire; PFS, Power of Food Scale; TFEQ, Three Factor Eating Questionnaire; YFAS, Yale Food Addiction Scale.

**Table S15.** ***Ad libitum*** **lunch taste ratings.**

| Dish type | Group comparison | Mean difference | SEM | 95% Confidence Interval | | Cohen's d | Post-hoc P values | |
|---|---|---|---|---|---|---|---|---|
| | | | | Lower bound | Upper bound | | LSD | Sidak |
| **Creamy taste rating** | | | | | | | | |
| Savoury | OB > ExS | 4.22 | 3.95 | -3.66 | 12.10 | 0.30 | 0.289 | 0.64 |
| | OB > AAD | 2.08 | 3.92 | -5.72 | 9.99 | 0.15 | 0.60 | 0.94 |
| | AAD > ExS | 2.14 | 3.92 | -5.66 | 9.95 | 0.15 | 0.59 | 0.93 |
| Sweet | OB > ExS | 15.6 | 5.09 | 5.46 | 25.74 | 0.87 | **0.003** | **0.009** |
| | OB > AAD | 0.78 | 5.04 | -9.26 | 10.82 | 0.04 | 0.88 | 1.0 |
| | AAD > ExS | 14.8 | 5.04 | 4.78 | 24.86 | 0.82 | **0.004** | **0.013** |
| Low fat | OB > ExS | 3.02 | 4.42 | -5.79 | 11.83 | 0.19 | 0.50 | 0.87 |
| | AAD > OB | 0.33 | 4.38 | -8.39 | 9.06 | 0.02 | 0.94 | 1.0 |
| | AAD > ExS | 3.35 | 4.38 | -5.37 | 12.08 | 0.22 | 0.45 | 0.83 |
| High fat | OB > ExS | 16.8 | 4.85 | 7.13 | 26.45 | 0.98 | **<0.001** | **0.003** |
| | OB > AAD | 3.19 | 4.80 | -6.39 | 12.76 | 0.19 | 0.51 | 0.88 |
| | AAD > ExS | 13.6 | 4.80 | 4.04 | 23.19 | 0.79 | **0.006** | **0.018** |
| **Ideal creaminess rating** | | | | | | | | |
| Savoury | ExS > OB | 2.64 | 3.62 | -4.58 | 9.86 | 0.21 | 0.47 | 0.85 |
| | ExS > AAD | 7.49 | 3.59 | 0.337 | 14.64 | 0.58 | **0.04** | 0.12 |
| | OB > AAD | 4.85 | 3.59 | -2.30 | 12.00 | 0.38 | 0.18 | 0.45 |
| Sweet | OB > ExS | 6.04 | 3.08 | -0.10 | 12.18 | 0.55 | 0.54 | 0.15 |
| | OB > AAD | 2.46 | 3.05 | -3.63 | 8.54 | 0.23 | 0.42 | 0.81 |
| | AAD > ExS | 3.59 | 3.05 | -2.50 | 9.67 | 0.33 | 0.24 | 0.57 |
| Low fat | ExS > OB | 1.26 | 2.99 | -4.52 | 7.04 | 0.12 | 0.67 | 0.96 |
| | ExS > AAD | 6.57 | 2.87 | 0.85 | 12.29 | 0.64 | 0.25 | 0.07 |
| | OB > AAD | 5.31 | 2.87 | -0.41 | 11.03 | 0.52 | 0.68 | 0.19 |
| High fat | OB > ExS | 4.66 | 3.77 | -2.85 | 12.17 | 0.35 | 0.22 | 0.53 |
| | OB > AAD | 1.99 | 3.73 | -5.45 | 9.43 | 0.15 | 0.60 | 0.93 |
| | AAD > ExS | 2.67 | 3.73 | -4.77 | 10.11 | 0.20 | 0.48 | 0.86 |

| Dish type | Group comparison | Mean difference | SEM | 95% Confidence Interval | | Cohen's d | Post-hoc P values | |
|---|---|---|---|---|---|---|---|---|
| | | | | Lower bound | Upper bound | | LSD | Sidak |
| **Taste liking rating** | | | | | | | | |
| Savoury | ExS > OB | 2.48 | 5.19 | -7.87 | 12.83 | 0.14 | 0.63 | 0.95 |
| | AAD > OB | 2.62 | 5.14 | -7.63 | 12.86 | 0.14 | 0.61 | 0.94 |
| | AAD > ExS | 0.14 | 5.14 | -10.11 | 10.38 | 0.01 | 0.98 | 1.0 |
| Sweet | OB > ExS | 7.78 | 5.21 | 2.60 | 18.16 | 0.42 | 0.14 | 0.36 |
| | AAD > OB | 7.89 | 5.16 | -2.38 | 18.17 | 0.43 | 0.13 | 0.34 |
| | AAD > ExS | 15.67 | 5.16 | 5.40 | 25.95 | 0.85 | **0.003** | **0.010** |
| Low fat | ExS > OB | 1.38 | 5.22 | -9.02 | 11.78 | 0.08 | 0.79 | 0.991 |
| | AAD > OB | 8.45 | 5.17 | -1.85 | 18.75 | 0.46 | 0.11 | 0.29 |
| | AAD > ExS | 7.07 | 5.17 | -3.23 | 17.37 | 0.38 | 0.18 | 0.44 |
| High fat | OB > ExS | 6.68 | 5.56 | -4.40 | 17.76 | 0.34 | 0.23 | 0.55 |
| | AAD > OB | 2.06 | 5.51 | -8.91 | 13.03 | 0.11 | 0.71 | 0.98 |
| | AAD > ExS | 8.73 | 5.51 | -2.23 | 19.71 | 0.44 | 0.12 | 0.31 |
| **Taste pleasantness** | | | | | | | | |
| Savoury | ExS > OB | 2.04 | 5.30 | -8.52 | 12.60 | 0.11 | 0.70 | 0.97 |
| | ExS > AAD | 0.18 | 5.25 | -10.28 | 10.64 | 0.01 | 0.97 | 1.0 |
| | AAD > OB | 1.86 | 5.25 | -8.60 | 12.32 | 0.10 | 0.72 | 0.99 |
| Sweet | OB > ExS | 6.52 | 4.33 | -2.10 | 15.14 | 0.43 | 0.14 | 0.36 |
| | AAD > ExS | 16.8 | 4.28 | 8.27 | 25.35 | 1.10 | **<0.001** | **<0.001** |
| | AAD > OB | 10.29 | 4.28 | 1.75 | 18.83 | 0.67 | **0.019** | 0.055 |
| Low fat | OB > ExS | 1.36 | 4.74 | -8.09 | 10.81 | 0.08 | 0.78 | 0.99 |
| | AAD > OB | 8.51 | 4.69 | -0.85 | 17.87 | 0.50 | 0.07 | 0.21 |
| | AAD > ExS | 9.87 | 4.69 | 0.52 | 19.23 | 0.58 | **0.04** | 0.11 |
| High fat | OB > ExS | 3.12 | 5.35 | -7.55 | 13.79 | 0.17 | 0.56 | 0.92 |
| | AAD > OB | 3.64 | 5.30 | -6.93 | 14.20 | 0.19 | 0.50 | 0.87 |
| | AAD > ExS | 6.76 | 5.30 | -3.81 | 17.32 | 0.36 | 0.21 | 0.50 |

Post-hoc group comparisons for taste ratings at *ad libitum* lunch between adults with obesity (OB, n=25), ex-smokers (ExS, n-25) and abstinent alcohol dependence (AAD, n=26). Analysed using repeated measures ANOVA with dish sweetness (savourty, sweet) and fat content (low fat, high fat) as within participant factors, with post-hoc Fisher LSD (uncorrected) or Sidak (corrected for multiple comparisons) tests. Abbreviations: LSD, least significant difference; SEM, standard error of the mean.

**Table S16. Spatial coordinates from exploratory whole brain analysis for cigarette cue reactivity during picture evaluation fMRI task between ex-smokers without and with alcohol dependence.**

| Group Contrast | Cluster | Voxels | Z max | x | y | z | Laterality | Location |
|---|---|---|---|---|---|---|---|---|
| ExS > AAD-ExS | 1 | 4799 | 4.28 | 28 | 18 | -8 | R | Insula (32%), Orbitofrontal cortex (12%) |
| | | | 4.12 | 30 | 26 | 18 | R | White matter (100%) |
| | | | 3.91 | 14 | 20 | 10 | R | Caudate (66%), Lateral ventricle (33%) |
| | | | 3.85 | 24 | -12 | 18 | R | White matter (100%) |
| | | | 3.84 | -16 | 20 | 38 | L | White matter (100%) |
| | | | 3.81 | 28 | -20 | 12 | R | White matter (84%), Putamen (16%) |
| AAD-ExS > ExS | Nil | | | | | | | |

Spatial coordinates of significant clusters from whole brain mixed effects analysis of cigarette picture (vs. neutral) contrast in picture evaluation fMRI task between groups as unpaired t-test: ex-smokers without history of alcohol dependence (ExS, n=25) and abstinent alcohol dependence group who were ex-smokers (AAD-ExS, n=11). Cluster-wise threshold Z>2.3, family-wise error (FWE) P<0.05. x ,y, z co-ordinates given in Montreal Neurological Institute (MNI) space. Location and percentage likelihood given using the Harvard-Oxford cortical and subcortical structural atlases, except [a] Sallet dorsal frontal connectivity-based parcellation atlas. Abbreviations: B, bilateral; R, right; L, left.

### SUPPLEMENTARY FIGURES

**Figure S1. GHADD study day protocol.**

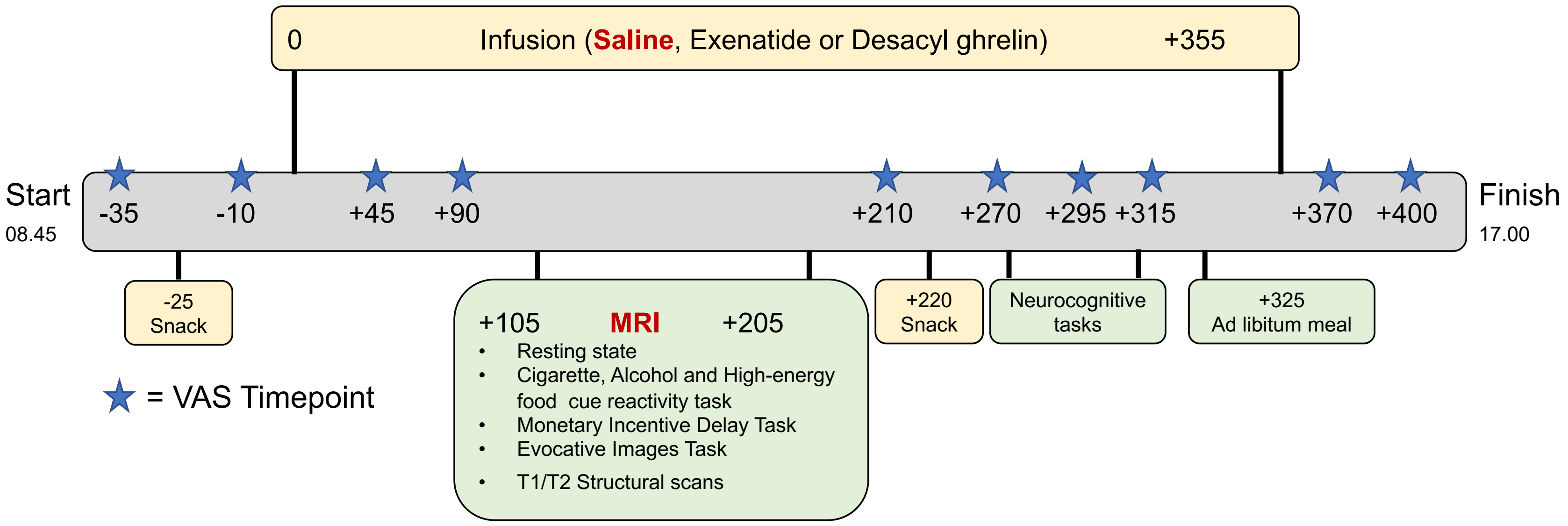

Timeline of study visit showing infusion, MRI scanning session, neurocognitive tasks, snacks, *ad libitum* test meal, and time points of visual analogue scale (VAS) and blood sampling (blue star).

**Figure S2. Functional regions of interest for picture evaluation fMRI task.**

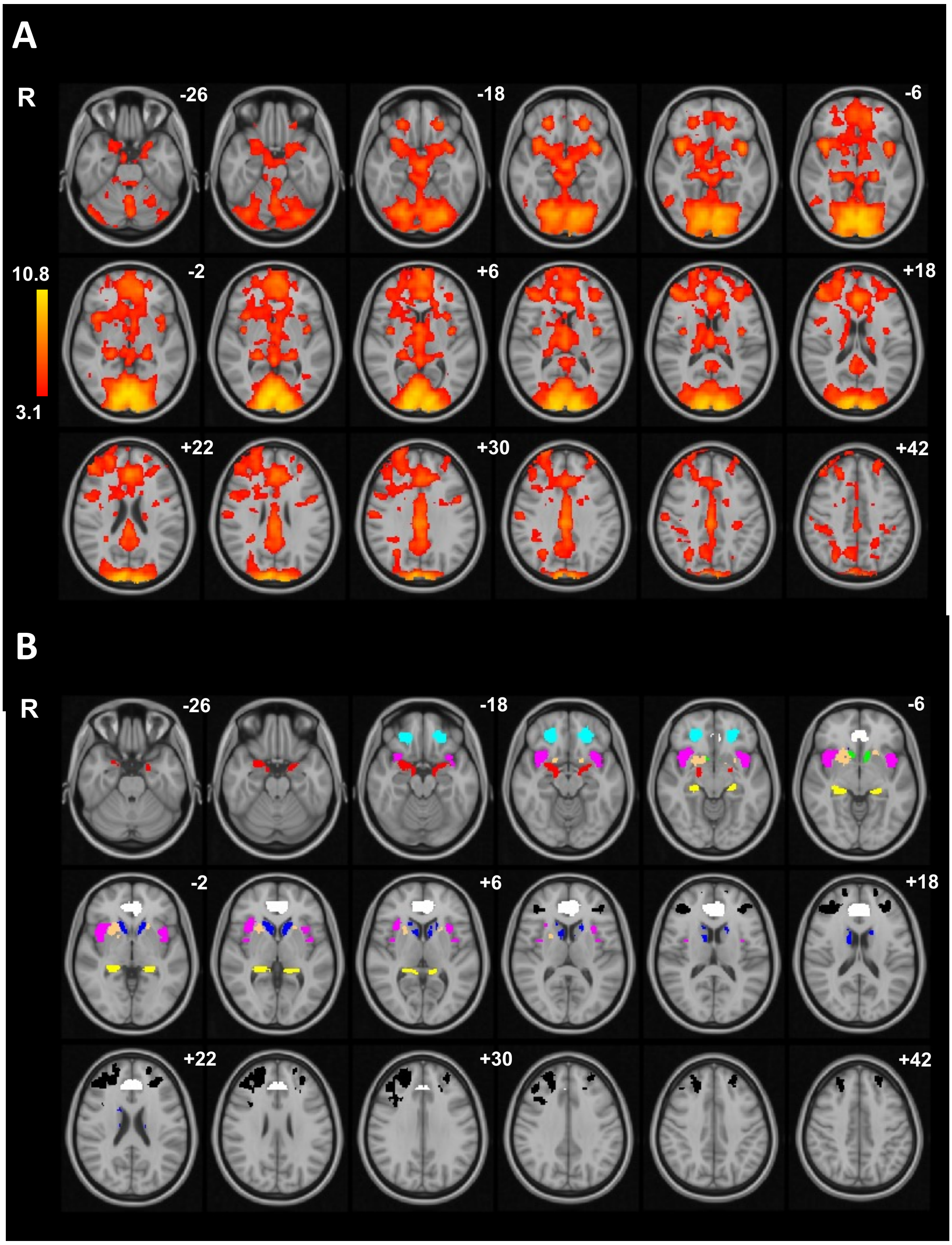

(A) Group activation maps for high energy food or alcohol (vs. neutral) pictures in separate cohort of healthy controls without obesity (n=43), cluster-wise threshold Z>3.1, family-wise error P<0.05 where colour bar indicates Z-score. (B) Individual functional regions of interest (fROIs): amygdala (Amyg), red; caudate (Caud), dark blue; dorsolateral prefrontal cortex (dlPFC), black; hippocampus (HPC), yellow; anterior insula (Ins), magenta; nucleus accumbens (NAcc), green; orbitofrontal cortex (OFC), light blue; putamen (Put), beige; ventral anterior cingulate cortex (vACC), white. z co-ordinates given in Montreal Neurological Institute (MNI) space. Abbreviations: R, right. See Supplementary Table S4 for coordinates.

**Figure S3. Whole brain analysis of high-energy food cue reactivity from picture evaluation fMRI task in obesity.**

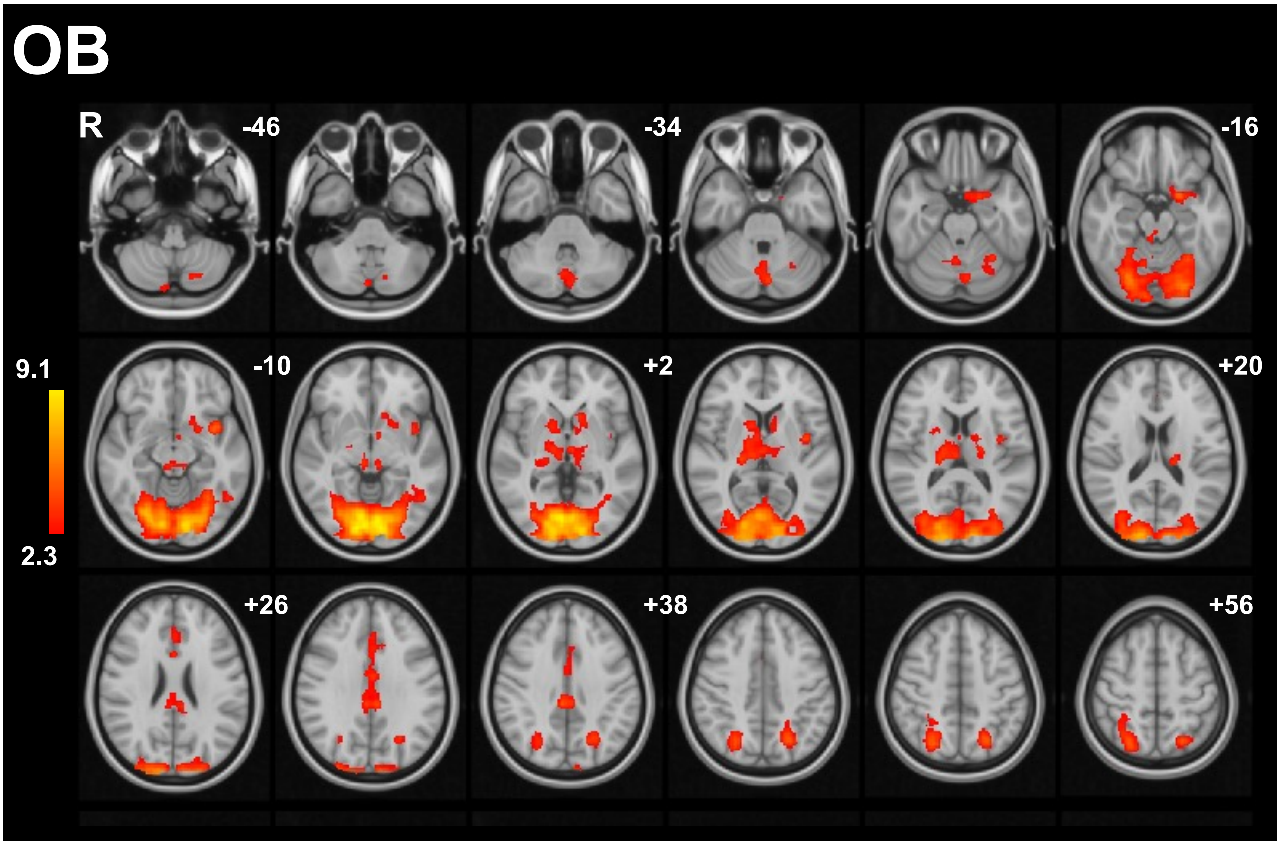

Group activation map for high-energy food pictures (vs. neutral) contrast in group with obesity (OB, n=25), cluster-wise threshold Z>2.3, family-wise error P<0.05. Colour bar indicates Z-score. z co-ordinates given in Montreal Neurological Institute (MNI) space. Abbreviations: R, right. See Supplementary Table S5 for coordinates.

**Figure S4. Whole brain analysis of high-energy food cue reactivity from picture evaluation fMRI task in ex-smokers.**

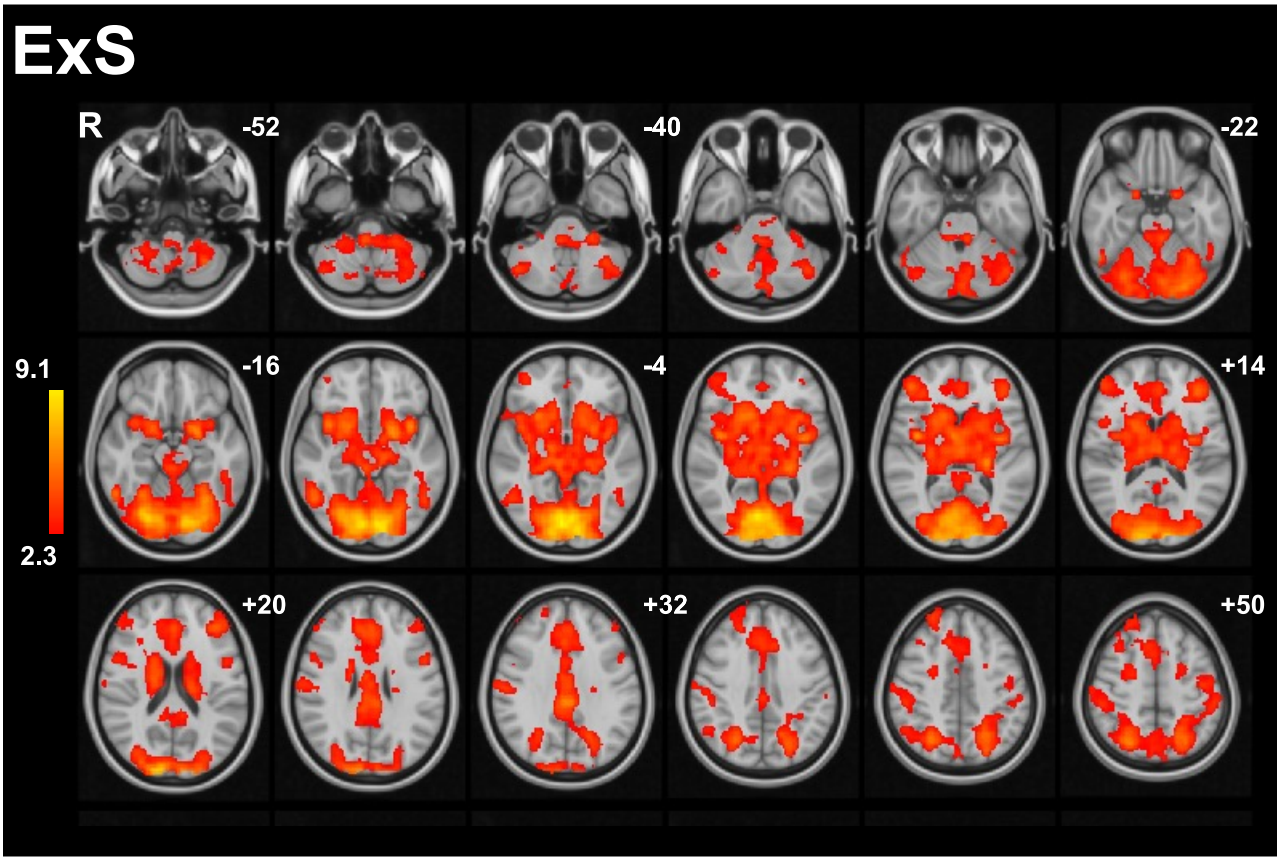

Group activation map for high-energy food pictures (vs. neutral) contrast in ex-smokers (ExS, n=25), cluster-wise threshold Z>2.3, family-wise error P<0.05. Colour bar indicates Z-score. z co-ordinates given in Montreal Neurological Institute (MNI) space. Abbreviations: R, right. See Supplementary Table S5 for coordinates.

**Figure S5. Whole brain analysis of high-energy food cue reactivity from picture evaluation fMRI task in abstinent alcohol dependence.**

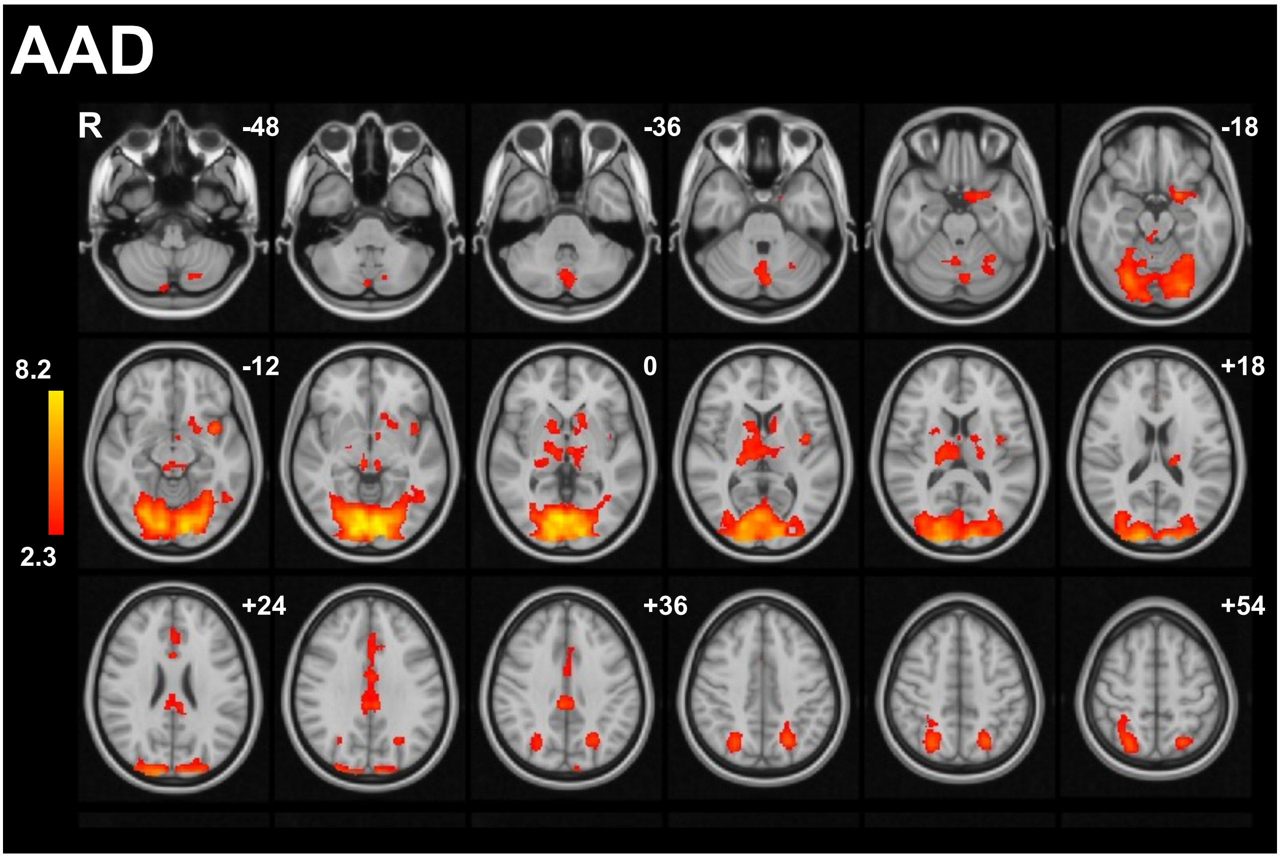

Group activation map for high-energy food pictures (vs. neutral) contrast in abstinent alcohol dependence (AAD, n=26), cluster-wise threshold Z>2.3, family-wise error P<0.05. Colour bar indicates Z-score. z co-ordinates given in Montreal Neurological Institute (MNI) space. Abbreviations: R, right. See Supplementary Table S5 for coordinates.

**Figure S6. Whole brain analysis of high-energy food cue reactivity from picture evaluation fMRI task for comparison between ex-smokers and other groups.**

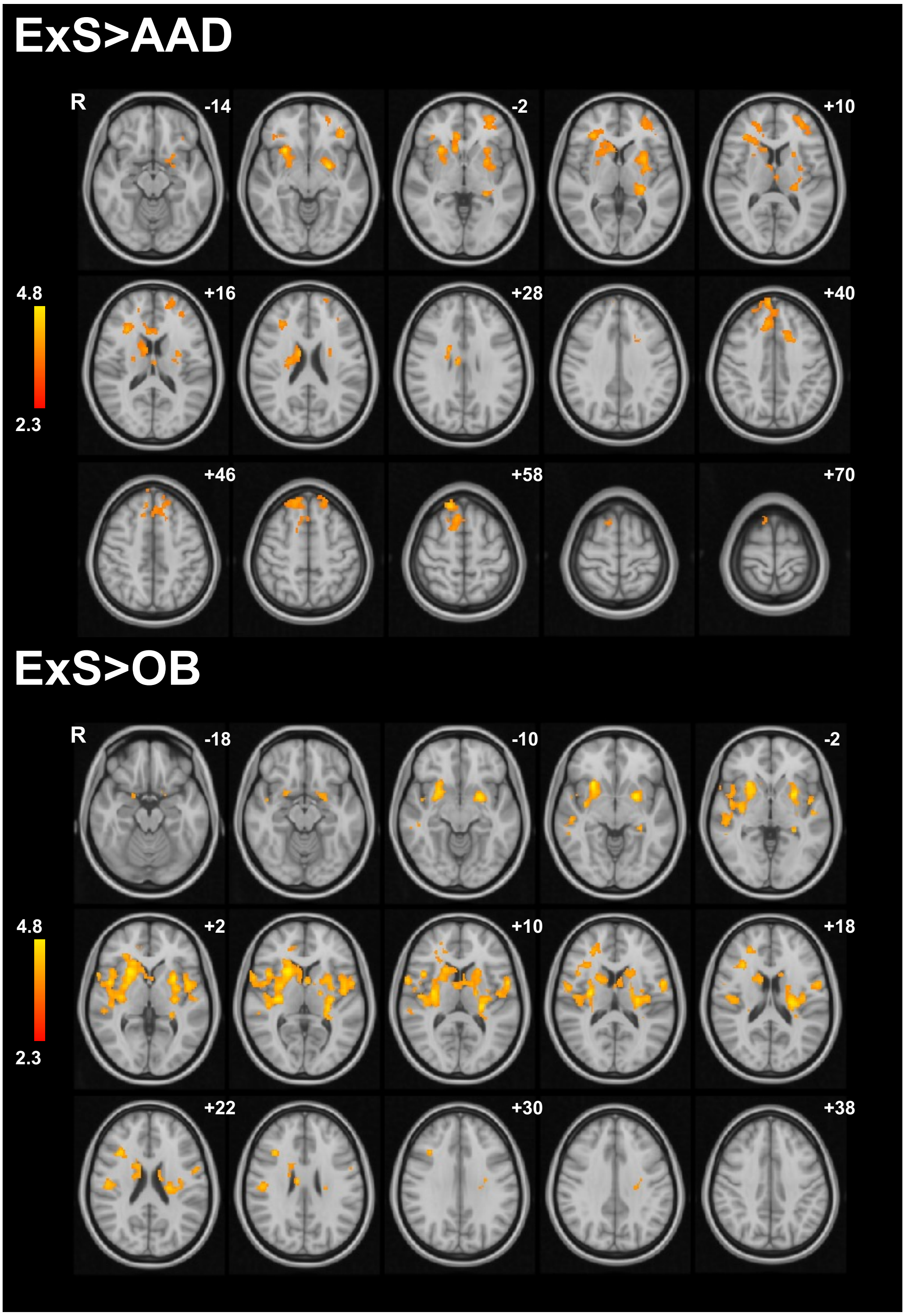

Group activation maps for high-energy food pictures (vs. neutral) contrast for (top) ex-smokers (ExS, n=25) > abstinent alcohol dependence (AAD, n=26), and (bottom) ex-smokers (ExS, n=25) > obesity (OB, n=25) between group comparisons. Cluster-wise threshold Z>2.3, family-wise error (FWE) P<0.05, unpaired t-test. Colour bar indicates Z-scores. z co-ordinates given in Montreal Neurological Institute (MNI) space. Abbreviations: R, right. See Supplementary Table S10 for coordinates.

**Figure S7. Group differences in alcohol cue reactivity from functional region of interest analysis.**

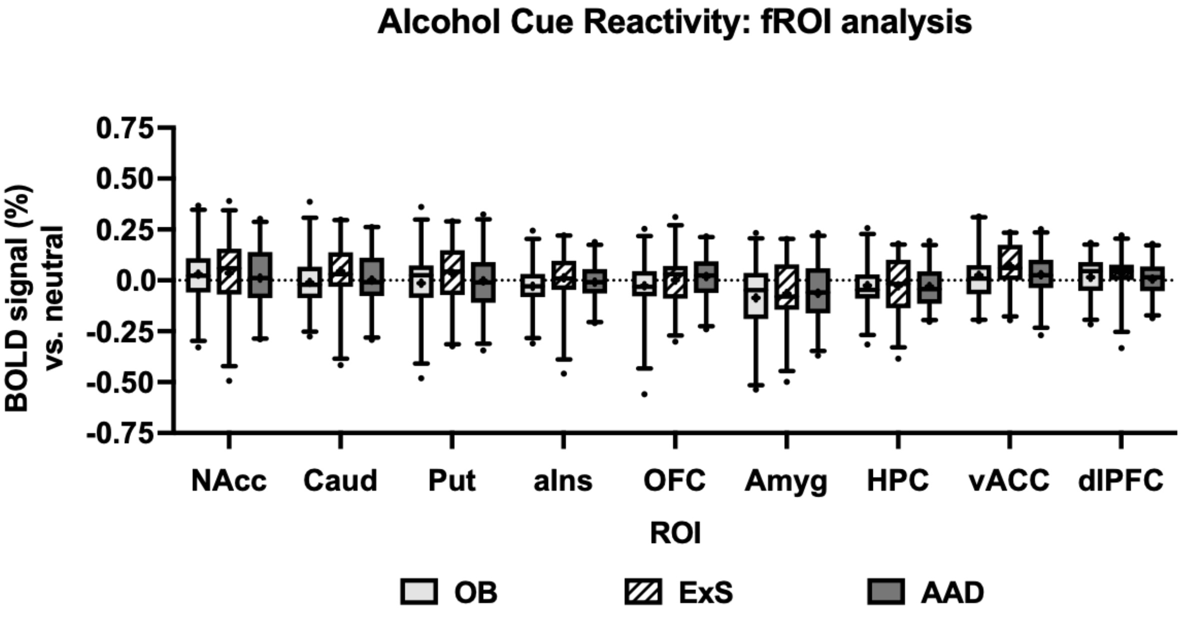

Percentage BOLD signal to preferred alcohol pictures (vs. neutral) during picture evaluation fMRI task in adults with obesity (OB, light grey), ex-smokers (ExS, hatched) and abstinent alcohol dependence (AAD, dark grey), in individual functional regions of interest (fROI): bilateral nucleus accumbens (NAcc), caudate (Caud), putamen (Put), anterior insula (Ins), anterior orbitofrontal cortex (OFC), amygdala (Amyg), hippocampus (HPC), ventral anterior cingulate cortex (vACC), and dorsolateral prefrontal cortex (dlPFC). Data presented as box plot with median (line), mean (+), interquartile range (box) and 5-95$^{th}$ percentiles (error bars), with outliers as a dot. Compared using one-way repeated measures ANOVA: no significant group x ROI interaction or main effects of group (P>0.05), in exploratory analysis no significant between group differences within each fROI using post-hoc LSD or Sidak tests (P>0.05).

**Figure S8. Whole brain analysis of alcohol cue reactivity from picture evaluation fMRI task in obesity.**

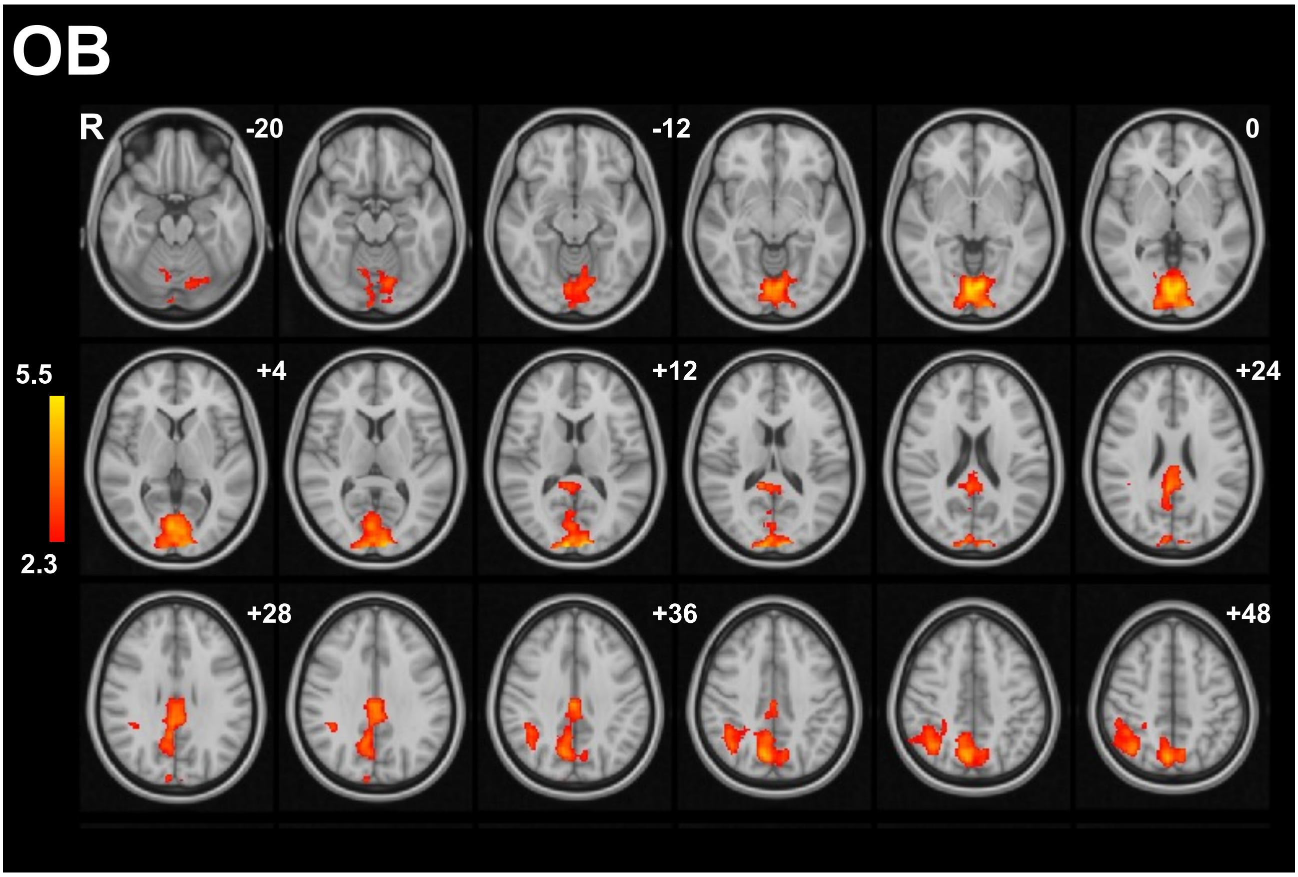

Group activation map for preferred alcohol pictures (vs. neutral) contrast in obesity (OB, n=25), cluster-wise threshold Z>2.3, family-wise error P<0.05. Colour bar indicates Z-score. z co-ordinates given in Montreal Neurological Institute (MNI) space. Abbreviations: R, right. See Supplementary Table S8 for coordinates.

**Figure S9. Whole brain analysis of alcohol cue reactivity from picture evaluation fMRI task in ex-smokers.**

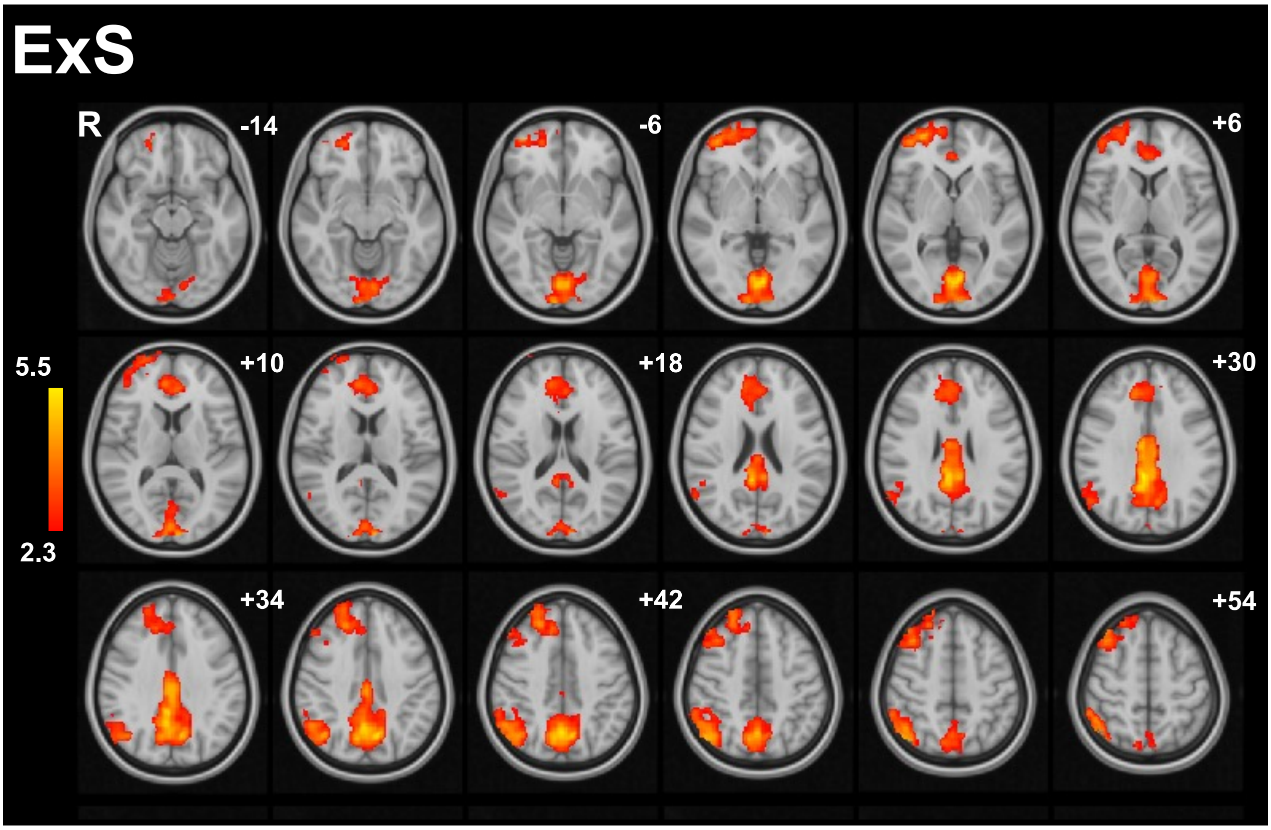

Group activation map for preferred alcohol pictures (vs. neutral) contrast in ex-smokers (exS, n=25), cluster-wise threshold Z>2.3, family-wise error P<0.05. Colour bar indicates Z-score. z co-ordinates given in Montreal Neurological Institute (MNI) space. Abbreviations: R, right. See Supplementary Table S8 for coordinates.

**Figure S10. Whole brain analysis of alcohol cue reactivity from picture evaluation fMRI task in abstinent alcohol dependence.**

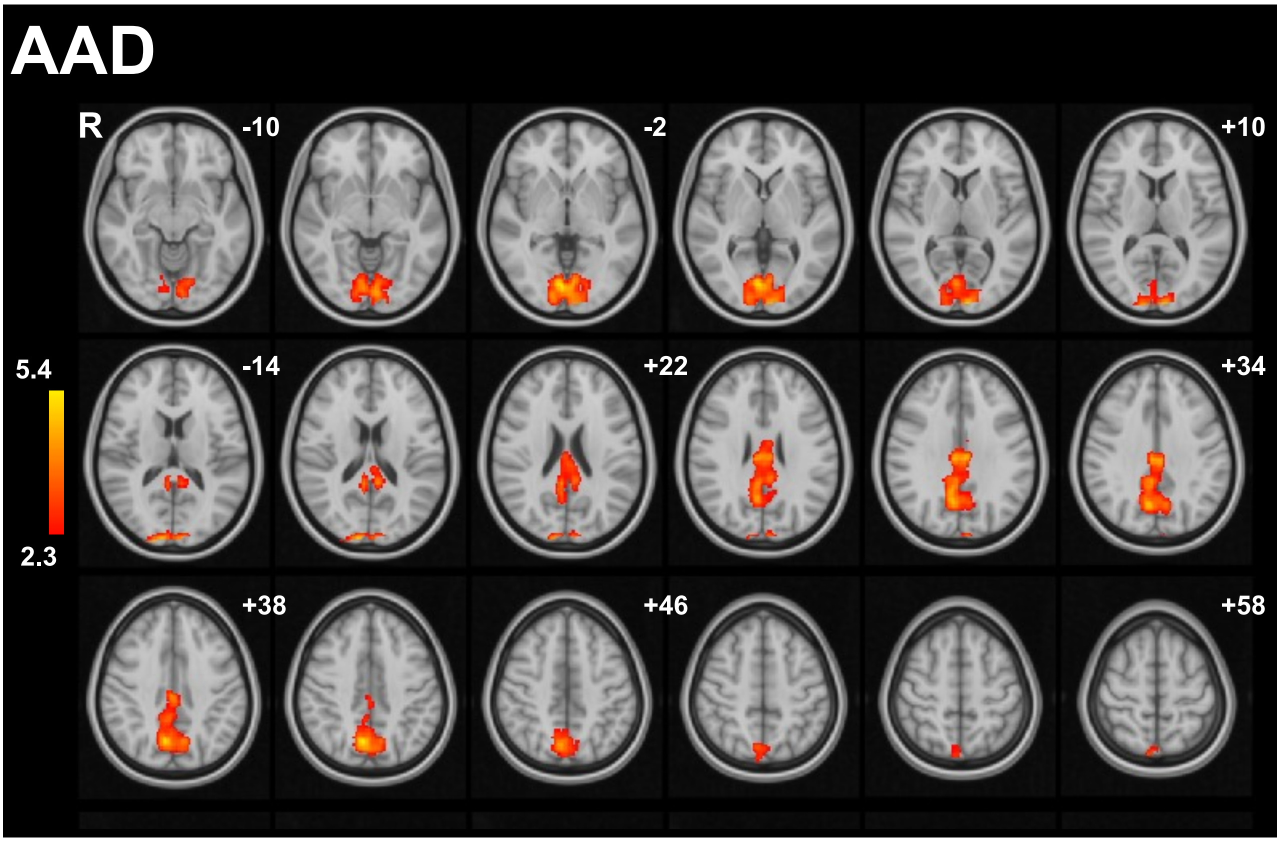

Group activation map for preferred alcohol pictures (vs. neutral) contrast in abstinent alcohol dependence (AAD, n=26), cluster-wise threshold Z>2.3, family-wise error P<0.05. Colour bar indicates Z-score. z co-ordinates given in Montreal Neurological Institute (MNI) space. Abbreviations: R, right. See Supplementary Table S8 for coordinates.

**Figure S11. Group differences in cigarette cue reactivity from functional region of interest analysis.**

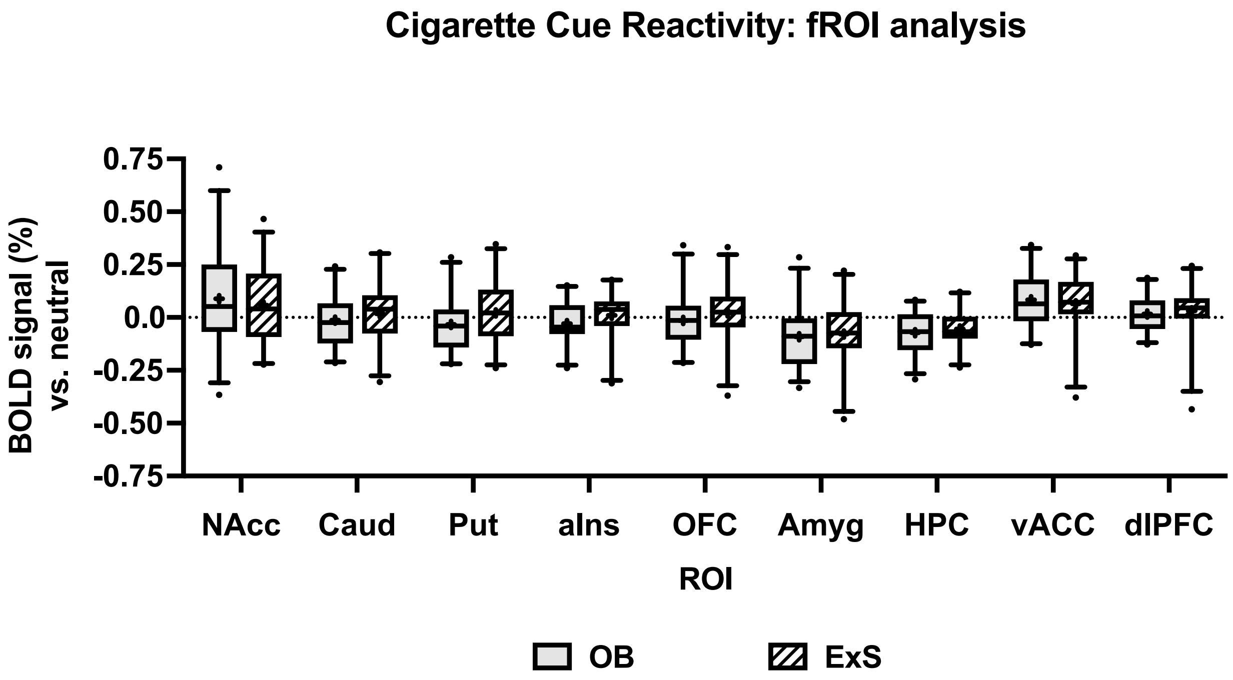

Percentage BOLD signal to cigarette pictures (vs. neutral) during picture evaluation fMRI task in adults with obesity (OB, light grey), and ex-smokers (ExS, hatched), in individual functional regions of interest (fROI): bilateral nucleus accumbens (NAcc), caudate (Caud), putamen (Put), anterior insula (Ins), anterior orbitofrontal cortex (OFC), amygdala (Amyg), hippocampus (HPC), ventral anterior cingulate cortex (vACC), and dorsolateral prefrontal cortex (dlPFC). Data presented as box plot with median (line), mean (+), interquartile range (box) and 5-95$^{th}$ percentiles (error bars), with outliers as a dot. Compared using repeated measures ANOVA: no significant group x ROI interaction or main effects of group (P>0.05), in exploratory analysis no significant between group differences within each fROI using post-hoc LSD or Sidak tests (P>0.05).

**Figure S12. Whole brain analysis of cigarette cue reactivity from picture evaluation fMRI task in obesity.**

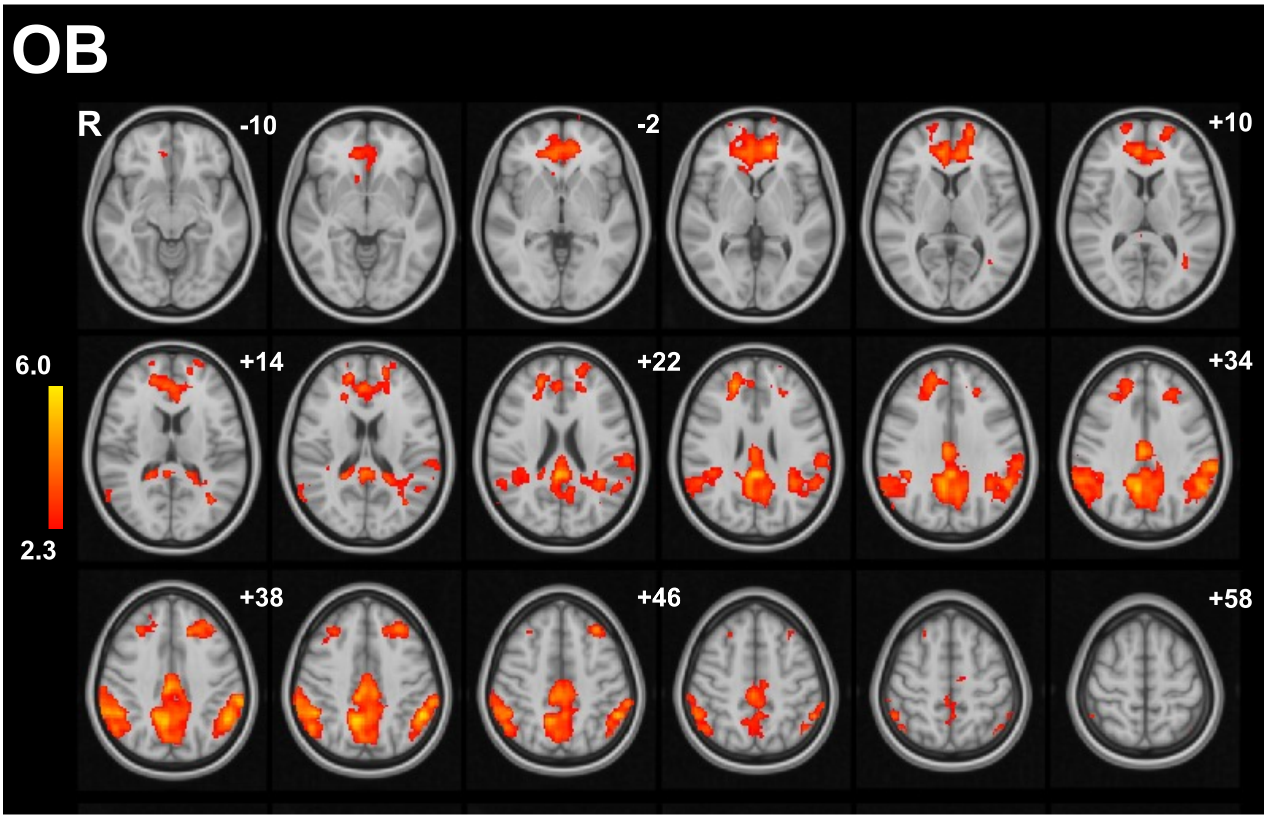

Group activation map for cigarette pictures (vs. neutral) contrast in obesity (OB, n=25), cluster-wise threshold Z>2.3, family-wise error P<0.05. Colour bar indicates Z-score. z co-ordinates given in Montreal Neurological Institute (MNI) space. Abbreviations: R, right. See Supplementary Table S9 for coordinates.

**Figure S13. Whole brain analysis of cigarette cue reactivity from picture evaluation fMRI task in ex-smokers.**

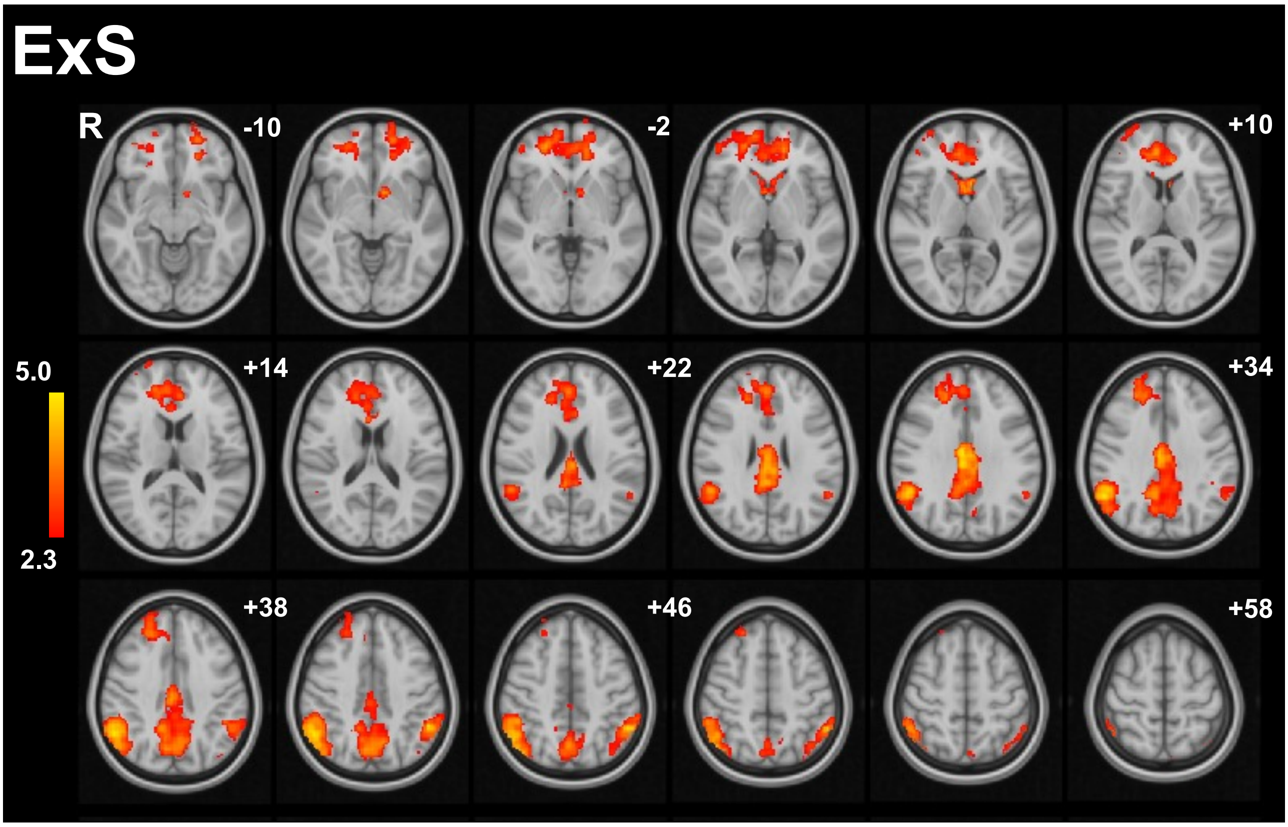

Group activation map for cigarette pictures (vs. neutral) contrast in ex-smokers (ExS, n=25), cluster-wise threshold Z>2.3, family-wise error P<0.05. Colour bar indicates Z-score. z co-ordinates given in Montreal Neurological Institute (MNI) space. Abbreviations: R, right. See Supplementary Table S9 for coordinates.

**Figure S14.** ***Ad libitum*** **lunch taste ratings for creamy taste and ideal creaminess.**

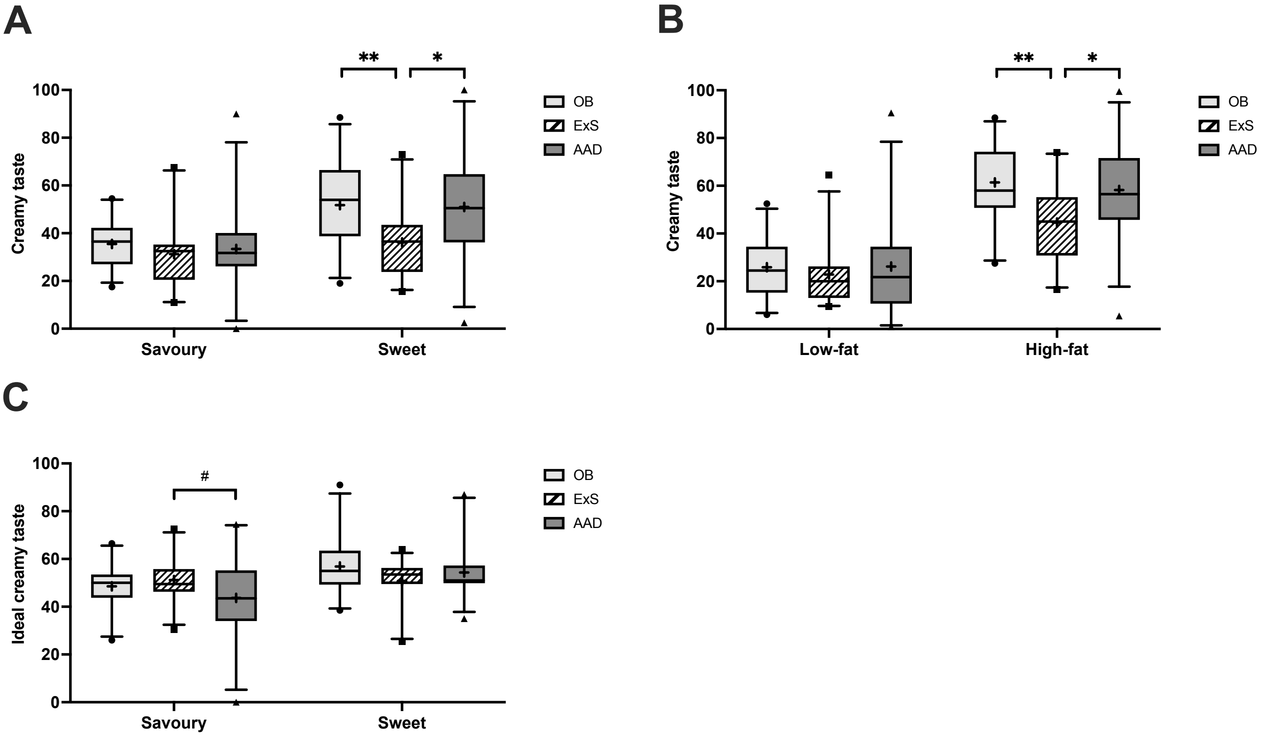

Group comparison of visual analogue scale ratings of (A) creamy taste for savoury and sweet dishes, (B) creamy taste for low-fat and high-fat dishes, and (C) ideal creaminess for savoury and sweet dishes (<50 below ideal, > 50 above ideal), in adults with obesity (OB, light grey, n=25), ex-smokers (ExS, hatched, n=25), and abstinent alcohol dependence (AAD, dark grey, n=26). Data presented as box plot with median (line), mean (+), interquartile range (box) and 5-95th percentiles (error bars), with outliers as a symbol. Groups compared using repeated measures ANOVA with post-hoc Sidak test: * P<0.05, ** P<0.01, or Fisher LSD test: # P<0.05.
